# The genetic case for cardiorespiratory fitness as a clinical vital sign and the routine prescription of physical activity in healthcare

**DOI:** 10.1101/2020.12.08.20243337

**Authors:** Ken B. Hanscombe, Elodie Persyn, Matthew Traylor, Kylie P. Glanville, Mark Hamer, Jonathan R. I. Coleman, Cathryn M. Lewis

**Author notes:** (corresponding author) Ken B. Hanscombe.

## Abstract

**Background:** Cardiorespiratory fitness (CRF) and physical activity (PA) are well-established predictors of morbidity and all-cause mortality. However, CRF is not routinely measured and PA not routinely prescribed as part of standard healthcare. The American Heart Association (AHA) recently presented a scientific case for the inclusion of CRF as a clinical vital sign based on epidemiological and clinical observation. Here, we leverage genetic data in the UK Biobank (UKB) to strengthen the case for CRF as a vital sign, and make a case for the prescription of PA.

**Methods:** We derived two CRF measures from the heart rate data collected during a submaximal cycle ramp test: CRF-vo2max, an estimate of the participants’ maximum volume of oxygen uptake, per kilogram of body weight, per minute; and CRF-slope, an estimate of the rate of increase of heart rate during exercise. Average PA over a 7-day period was derived from a wrist-worn activity tracker. After quality control, 70,783 participants had data on the two derived CRF measures, and 89,683 had PA data. We performed genome-wide association study (GWAS) analyses by sex, and post-GWAS techniques to understand genetic architecture of the traits and prioritize functional genes for follow-up.

**Results:** We found strong evidence that genetic variants associated with CRF and PA influenced genetic expression in a relatively small set of genes in heart, artery, lung, skeletal muscle, and adipose tissue. These functionally relevant genes were enriched among genes known to be associated with coronary artery disease (CAD), type 2 diabetes (T2D), and Alzheimer’s disease (three of the top 10 causes of death in high-income countries) as well as Parkinson’s disease, pulmonary fibrosis, and blood pressure, heart rate, and respiratory phenotypes. Genetic variation associated with lower CRF and PA was also correlated with several disease risk factors (including greater body mass index, body fat and multiple obesity phenotypes); a typical T2D profile (including higher insulin resistance, higher fasting glucose, impaired beta-cell function, hyperglycaemia, hypertriglyceridemia); increased risk for CAD and T2D; and a shorter lifespan.

**Conclusions:** Genetics supports three decades of evidence for the inclusion of CRF as a clinical vital sign. Given the genetic, clinical, and epidemiological evidence linking CRF and PA to increased morbidity and mortality, regular measurement of CRF as a marker of health and routine prescription of PA could be a prudent strategy to support public health.

## Background

Cardiorespiratory fitness (CRF) and physical activity (PA) are well-established predictors of morbidity and mortality (1, 2). CRF is such a strong predictor of cardiovascular health, all-cause mortality, and mortality attributable to various cancers, that in 2016 the American Heart Association (AHA) put forward a scientific case for measuring CRF as a clinical vital sign (3). The AHA recommended that, “(at) a minimum, all adults should have CRF estimated each year (…) during their annual healthcare examination”. Non-exercise testing methods for estimating CRF make this possible in a clinical setting (4). The AHA also noted that measurement of CRF provides “clinicians the opportunity to counsel patients regarding the importance of performing regular physical activity”. That exercise is medicine, preventative and curative, has been known from antiquity – Hippocrates wrote the earliest known prescription for exercise about 2000 years ago (5). Today however, we do not yet routinely prescribe PA, nor do we measure CRF as part of a standard clinical assessment.

Sedentary lifestyles are a significant public health problem. For example, physical inactivity and poor diet were the second leading modifiable, behavioural cause of death in the US in 2000 and are expected to overtake tobacco to become the leading cause (6). Worldwide, physical inactivity was estimated to account for about 10% of premature mortality in 2008, an effect size similar to smoking and obesity (7). In 2013, the economic burden due to direct health-care costs, productivity losses, and disability-adjusted life-years attributable to physical inactivity was estimated at US$53.8 billion (8). This economic burden is reflected in the prevalence of physical inactivity worldwide: 28% in 2016 (23% for men, 32% for women) (9). Most people do not know what the recommended activity guidelines to maintain good health are. A UK British Heart Foundation (BHF) survey estimated about 60% of adults in the UK do not know the recommended minimum level of PA (10). Modifiable physiological characteristics like CRF and behaviours like regular exercise are affected by both environmental and genetic factors. Genetic factors, not addressed in the AHA review, can influence CRF and PA through multiple mechanisms including appetite for leisure activity, physiological response to exercise, and CRF response to PA. Critically, genetics can be leveraged to identify links between CRF, PA, and disease. Here we present the genetic case for the measurement of CRF as a clinical vital sign, and the routine prescription of PA.

CRF is typically measured with incremental exercise (resistance or workload) on a treadmill or cycle ergometer. In a sub-maximal test, CRF is estimated from extrapolating the workload to the age-estimated maximum heart rate. PA has most often been measured by self-report. Previous research has shown that CRF and PA are familial, with family and twin study estimates of their heritability varying widely (CRF: *h*^*2*^=0.25 – 0.65 (11); PA: *h*^2^=0 – 0.78, and 0.48 in the only study that objectively measured PA (12)). There has been some interest in variation in CRF response to exercise. Several candidate gene studies and one genome-wide association study (GWAS) have reported genetic variants associated with baseline CRF, or CRF change in response to exercise, but none was genome-wide significant or reliably replicated (13-15). Two GWASs of PA in the UK Biobank (UKB) reported three loci associated with accelerometer-measured 7-day average PA (SNP heritability, *h*^*2*^_SNP_ =0.14), and none with self-reported PA (16, 17). The genetic contribution to CRF is poorly characterised, and our current understanding of the genetics of PA has not examined differences by sex.

In this study, we performed the largest genome-wide association analysis of CRF, a GWAS of PA, and related their genetic risk profiles to the known genetic risk for anthropomorphic phenotypes, metabolic traits, and chronic diseases. CRF and PA were objectively measured in the UKB (18). Given the multitude of mechanisms by which PA and CRF affect biology (3, 15, 19), the differences in physiognomy (e.g., body fat distribution (20, 21)), behavioural dynamics (e.g., motivators and context preferences (22)), we investigated sex-specific as well as sex-combined CRF and PA phenotypes. Previous UKB analyses of PA have reported genetic correlations between the sexes and included sex as a covariate in combined analyses (17), which by design identifies only genetic factors common to both sexes. Here, we also investigated the potential functional effects of sex-specific associations. Our aim in this study was to characterise the genetic contribution to CRF and PA highlighting genetic links to disease, and thus to strengthen the case for the inclusion of CRF as a clinical vital sign and make more cogent the argument for routine prescription of regular PA in healthcare.

## Methods

### Sample

#### UK Biobank

The UK Biobank (UKB) is a research study of over 500,000 individuals from across the United Kingdom. Participants aged 40 to 69 were invited to attend one of 22 assessment centres between 2006 and 2010. A comprehensive range of health-related data has been collected at baseline and follow up, including disease diagnoses and lifestyle information, with blood samples taken for genome-wide genotyping and a biomarker panel. Further data are available in follow up studies on different subsets of UKB participants, including CRF and PA data. All participants provided electronic signed consent (18, 23).

After selecting the European subsample (the largest cluster from a 4-means clustering of the first 2 UKB-derived principal components), and phenotype and genotype QC described below, we had 70,783 (34,419 male, 36,364 female) participants with CRF data, and 89,683 (39,352 male, 50,331 female) participants with PA data. Additional file 1: **Table S1** shows details of the derivation of the final GWAS sample.

### Phenotypes

#### Cardiorespiratory fitness – submaximal cycle ramp test

CRF was assessed with a submaximal cycle ramp test which used a stationary bicycle and 4-lead electrocardiograph (ECG) to record heartrate data during pre-test, exercise and rest phases (approximately 15 seconds, 6 minutes, and 1 minute respectively). The cycle test procedure is described in detail in the UKB Cardio Assessment document (24). Cycle ramp test data were collected at two instances: from August 2009 at the end of the initial assessment visit 2006–2010 (instance 0), and the first repeat assessment visit 2012–2013 (instance 1). Participants performed an exercise protocol determined by their risk level, *Program Category* (UKB field 6024): ‘minimal’, ‘small’, ‘medium’, or ‘high’. The cycle ramp test included a ‘Pre-test’, ‘Exercise’, and ‘Recovery’ phase. We used data from the ‘Exercise’ phase only, within which, only ‘minimal’ and ‘small’ risk categories included a workload ramp (increase in workload). We selected samples in the ‘minimal’, ‘small’, and ‘medium’ risk categories, with *ECG Bike Method for Fitness Test* (6019) = ‘Bicycle’ (Additional file 1: **Table S2**), modelling heart rate data from the 4-minute workload ramp for the ‘small’ and ‘minimal’ risk groups, and the full 6-minute constant work load for the ‘medium’ risk group. We also included risk category as a covariate in the genetic association analyses.

The raw heart rate data were noisy (Additional file 1: **Figure S1**, panel **A**, shows raw heart rate data for 1,000 random participants). Participants were instructed to cycle at a cadence of 60 revolutions per minute (RPM) and we selected workload and heart rate data where participants cycled in the range 35-125 RPM (Additional file 1: **Figure S1**, panel **B**), within the manufacturer guaranteed range for cycle ergometer workload accuracy. Finally, we selected samples with a minimum of 20 observations of heart rate, workload, and cadence within the ‘Exercise’ phase of the test, and applied a Butterworth filter to the heart rate data (**Figure 1**). Full cycle ramp test QC is described in the supplementary Methods and listed in Additional file 1: **Table S1**.

**Figure 1.**
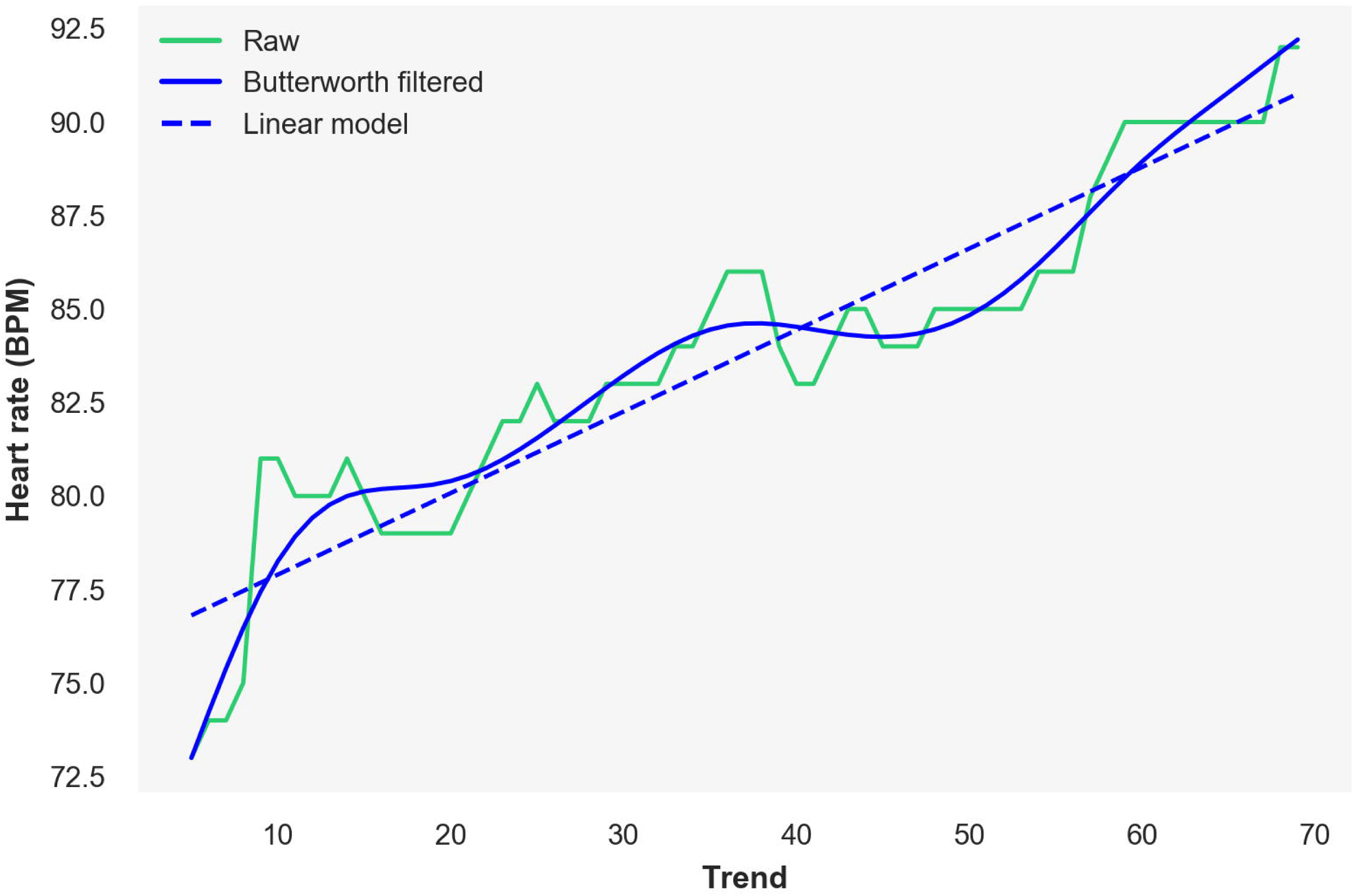
Heart rate signal processing for a single random sample. Trend = repeated measurements within the ‘Exercise’ phase of the submaximal cycle ramp test; Raw = unaltered heart rate data; Butterworth filtered = heart rate data after low-pass Butterworth filter (heart rate at the beginning and end of the exercise period on this filtered signal used to calculate CRF-vo2max); Linear model = fitted heart rate data values from a linear model fitted to the Butterworth filtered signal (CRF-slope is the slope of this linear model).

From this data, we derived two measures: *CRF-vo2max*, an estimate of the maximum volume of oxygen uptake per kilogram of body weight per minute; and *CRF-slope*, the rate of increase of heart rate. Specifically, we fit a linear model to workload and heart rate and extrapolated from the linear model to predict workload at the age-estimated maximum heart rate (HR·age·max = 208 – 0.7 * age, where age is age when attended assessment centre). Maximum workload relative to body weight in kilograms, or relative power (watts/kg), was then used as a proxy for CRF-vo2max:

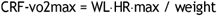

Where WL·HR·max is extrapolated work load at age-estimated maximum heart rate. The coefficients 208 and 0.7 for age-estimated maximum heart rate are commonly used constants and were used in previous studies of CRF in the UKB (25-28).

For CRF-slope, we fitted a linear model to the Butterworth-filtered heart rate signal for each participant and retained the slope, *β*_1_, as rate of increase of heart rate (**Figure 1**):

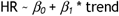

Where HR is heart rate and ‘trend’ refers to the time points at which repeated measurements of heart rate, workload, and cadence were taken within each phase of the submaximal cycle ramp test (**Figure 1**). Samples with negative slopes were excluded.

### Physical activity – wrist-worn accelerometer

PA in the UKB was measured continuously over a period of 7 days with a wrist-worn accelerometer in 103,712 participants, as an add-on measure after the initial assessment between June 2013 and January 2016. We used a wear-time adjusted 7-day average PA and filtered participants on the following test-specific variables: *Data quality, good wear-time* (90015)=‘Yes’, *Data quality, good calibration* (90016)=‘Yes’, and no *Data problem indicator*s (90002). Finally, we excluded samples with PA (accelerometer) > 100 milligravities (m*g*). General PA quality control and calibration of raw accelerometer data and calculation of derived activity level are described in detail elsewhere (29).

### Disease frequency

The top 10 global causes of death in high-income countries in 2016, as reported by the World Health Organization (WHO), are shown in Additional file 1: **Table S3**, along with the relevant International Classification of Diseases revision 10 (ICD-10) codes. Ischaemic heart disease was the number 1 global cause of death among high-income countries; lower respiratory tract infections, ranked 6^th^, was the only communicable cause of death on the list. We used UKB in-patient hospital episode statistics (diagnoses recorded in ICD-10 codes) to retrieve disease frequency for the top 10 global causes of death, among the subsample with CRF or PA data (Additional file 1: **Table S3** includes ICD-10 codes). We ranked participants by level of the phenotypes CRF-vo2max, CRF-slope, and PA after correcting for age and socioeconomic status (SES, Townsend deprivation index), divided the participants into ten approximately equal-sized groups, and calculated the proportion in each group with a main or secondary diagnosis (made during a hospital in-patient admission) for each of the 10 query diseases. ICD-9 diagnoses were not included. Firstly, it is not clear how to map ICD-9 codes onto the ICD-10 codes provided by the WHO. Secondly, ICD-9 codes were observed at very low frequencies: 1 sample with CRF data, and 40 with PA data, were diagnosed with ischaemic heart disease (ICD-9 codes starting with 41[0-9]), the most prevalent of the top 10 global causes of death. Diagnoses were potentially received both before and after the cycle ramp test and the week in which participants wore the physical activity tracker, and we made no distinction between incident and prevalent disease. This analysis was performed separately in males and females, since there are differences in both disease prevalence, and in phenotype distribution by sex.

### Genetic analyses

The full UKB sample were genotyped on one of two custom Applied Biosystems genotyping arrays: 49,950 on the UK Biobank Lung Exome Variant Evaluation (UK BiLEVE) Array, and 438,427 on the UK Biobank Axiom Array. The genetic data underwent standard GWAS QC and were imputed using IMPUTE4 to three reference panels by the UKB: the Haplotype Reference Consortium (HRC) and the merged UK10K and 1000 Genomes phase 3 reference panels. General genotyping considerations, raw genotype data QC, and genetic imputation procedures in the UKB are described in detail elsewhere (18). Details of available genetic data and associated metadata are described in UKB Resource 664 (30).

From UKB-supplied sample QC information, we excluded gender mismatches, samples with putative sex chromosome aneuploidy or excess relatives, and heterozygosity and missingness outliers. We performed linear mixed model association on the residual of each trait regressed on the covariates age, age^2^, sex (for sex-combined analyses), array, centre, and the first 10 principal components calculated on subsamples with data on the trait of interest. Additionally, for CRF phenotypes, we included the covariates risk category (corresponding to CRF test protocol) and number of trend entries (number of observations). Full details of GWAS QC and association parameters, alternative models including BMI, and alcohol and smoking as covariates, and post-GWAS analyses including transcriptome-wide association (TWAS) and colocalization, gene-set enrichment, and genetic correlation, are included in the supplementary methods. We used the standard p=5×10^−8^ as our genome-wide significant threshold, a conservative Bonferroni-corrected p-value for TWAS significance correcting for all genes tested in all tissues (p=0.05/75,486=6.6×10^−7^), and a false-discovery rate (FDR) correction for gene-set enrichment and genetic correlation accepting FDR-adjusted p-values<0.05 as significant.

### Analysis tools

For data analysis and figure creation we used both R (31) and Python 3 (32), in conjunction with the bioinformatic pipeline software Snakemake (33). We used the R package ukbtools (34) to parse all UKB data. Genetic data analyses were performed using FlashPCA (35), BOLT-LMM (36), PLINK 1.9 and 2.0 (37), LDSC (38), FUSION (39), and the web-based tools FUMA (40), LD Hub (41), and LDlink (42).

## Results

### Descriptive statistics and smoking and alcohol drinking status

Descriptive statistics for the CRF and PA phenotypes showed higher values for CRF-vo2max in males, and higher CRF-slope and PA in females (Additional file 1: **Table S4**). Correlations between CRF and PA phenotypes, and age and BMI were all modest and negative (*r*=-0.02 – -0.27), except for CRF-slope and age which had a small positive correlation in females (*r*=0.13, p=1.2×10^−144^) (Additional file 1: **Figure S2**). Combined-sex sample correlations between CRF-vo2max and CRF-slope, and SES were not significant; the PA and SES, and sex-specific correlations for all three traits were significant but small (| *r* |<=0.03). In the combined-sex sample, phenotypic correlations between our three traits were in the directions expected. CRF-vo2max and PA had a modest positive correlation (*r*=0.25, p=1.5×10^−195^, among the subsample who performed both the submaximal cycle ramp test and participated in the 7-day PA monitoring, N=13,561). CRF-slope and PA were not significantly correlated (*r*=-0.01, p=0.11, N=13,561). The two fitness phenotypes, CRF-vo2max and CRF-slope were negatively correlated (*r*=-0.39, p<0.001, N=70,783).

As a proxy for true resting heart rate, heart rate at the beginning of the exercise phase of the cycle ramp test was negatively correlated with CRF-vo2max (*r*=-0.33, p<0.001), and had a near zero correlation with CRF-slope (*r*=-0.03, p=3.8×10^−13^).

Smoking and alcohol drinking status are sometimes considered confounding factors in analyses of CRF and PA. In our data, there were small differences in mean CRF-vo2max, CRF-slope, and PA by smoking and drinking status (Additional file 1: **Table S5**), which contributed little to trait variance in linear regression models of all three phenotypes (ΔR^2^<=0.005 compared to a base model of age and sex, Additional file 1: **Table S6**). We chose not to adjust for the effects of smoking and drinking status in order to obtain unbiased estimates of the genetic factors on CRF and PA. There is evidence that correcting for heritable covariates can lead to biased estimates of the influence of genetic factors (43).

### Disease frequency

For the top 10 global causes of death in high-income countries in 2016, as reported by the World Health Organization (WHO), **Figure 2** shows the frequency at which each disease was observed in the UKB by level of CRF-vo2max, CRF-slope, and PA (after correcting for age and SES). Given the difference in the phenotype distributions of the male and female subsamples, and the inherent difference in disease frequency by sex, disease frequency has to be interpreted separately by sex.

**Figure 2.**
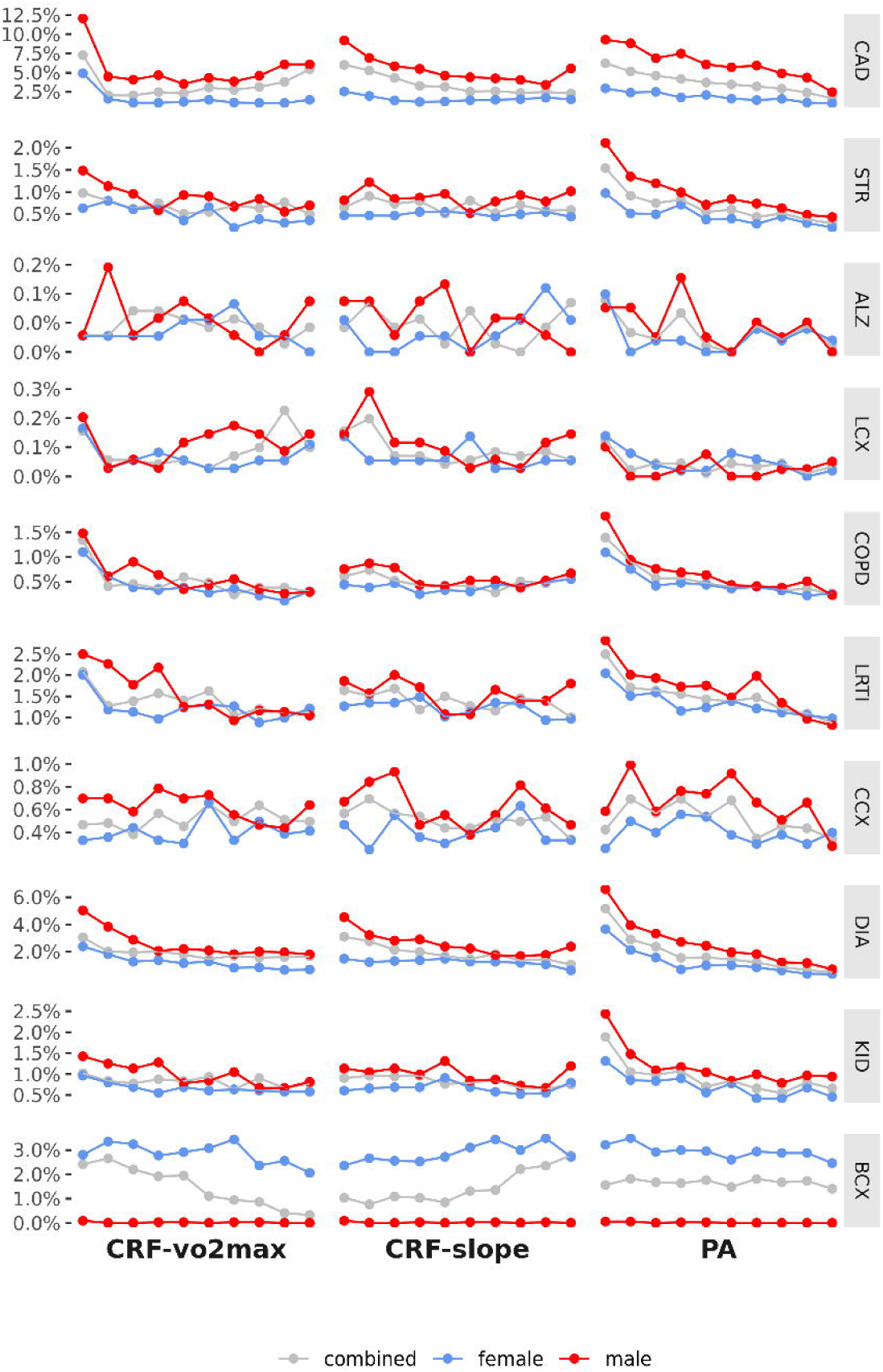
ICD-10 diagnosis frequency by CRF and PA level. Frequency in the UKB for the top 10 global causes of death in high-income countries in 2016, as reported by the World Health Organization (WHO, Additional file 1: **Table S3**). The CRF and PA phenotypes above were controlled for age and socioeconomic status (SES, the Townsend deprivation index) – they are the residuals from a regression on age and SES. For some diseases, e.g., breast cancer, disease trends are only meaningful separated by sex. CAD = ischaemic heart disease; STR = stroke; ALZ = Alzheimer’s disease and other dementias; LCX = trachea, bronchus and lung cancers; COPD = chronic obstructive pulmonary disease; LRTI = lower respiratory tract infections; CCX = colon and rectum cancers; DIA = diabetes mellitus; KID = kidney diseases; BCX = breast cancers

### Disease frequency by CRF

With increasing levels of CRF-vo2max, there were reductions in diagnosis frequency for ischaemic heart disease (CAD) (male: 12.0–6.1%, female: 4.9–1.5%), stroke (male: 1.5–0.7%, female: 0.6–0.4%), chronic obstructive pulmonary (COPD) (male: 1.5–0.3%, female: 1.1–0.3%), lower respiratory tract infection (LRTI) (male: 2.5–1.0%, female: 2.0–1.2%), diabetes mellitus (male: 5.0–1.8%, female: 2.4–0.7%), and kidney diseases (male: 1.4–0.8%, female: 1.0–0.6%). The largest differences were observed for CAD and diabetes, with the starkest reductions for all disease often seen within the lowest 4 deciles of CRF-vo2max.

CRF-slope showed reduction in disease for CAD (male: 9.2–5.6%, female: 2.6–1.5%) and diabetes (male: 4.5– 2.4%, female: 1.5–0.6%). The relationships with other disease diagnoses were more noisy.

### Disease frequency by PA

Disease diagnosis by level of PA showed similar patterns to CRF-vo2max, with a continued decile-to-decile reduction in disease across the range of PA from least to most physically active. With increasing PA we observed reductions in CAD (male: 9.3–2.5% female: 3.0–1.0%), stroke (male: 2.1–0.4%, female: 1.0–0.2%), COPD (male: 1.8–0.2%, female: 1.1–0.3%), LRTI (male: 2.8–0.8%, female: 2.0–1.0%), diabetes mellitus (male: 6.0–0.7%, female: 3.7–0.4%), and kidney diseases (male: 2.4–0.9%, female: 1.3–0.5%). Like CRF-vo2max, the greatest reductions were seen in CAD and diabetes mellitus.

### Genetic associations

Genome-wide association studies were performed for CRF-vo2max, CRF-slope, and PA with results for male, female, and the combined sample (**Figure 3**). An annotated list of the independent genome-wide significant variants, determined by LD clumping of the association results, is included in **Table 1**. Additional file 1: **Figures S3–S37** are the regional plots for each sex-combined GWAS signal peak, and sex-specific GWAS signal peaks where there was evidence of heterogeneity.

**Figure 3.**
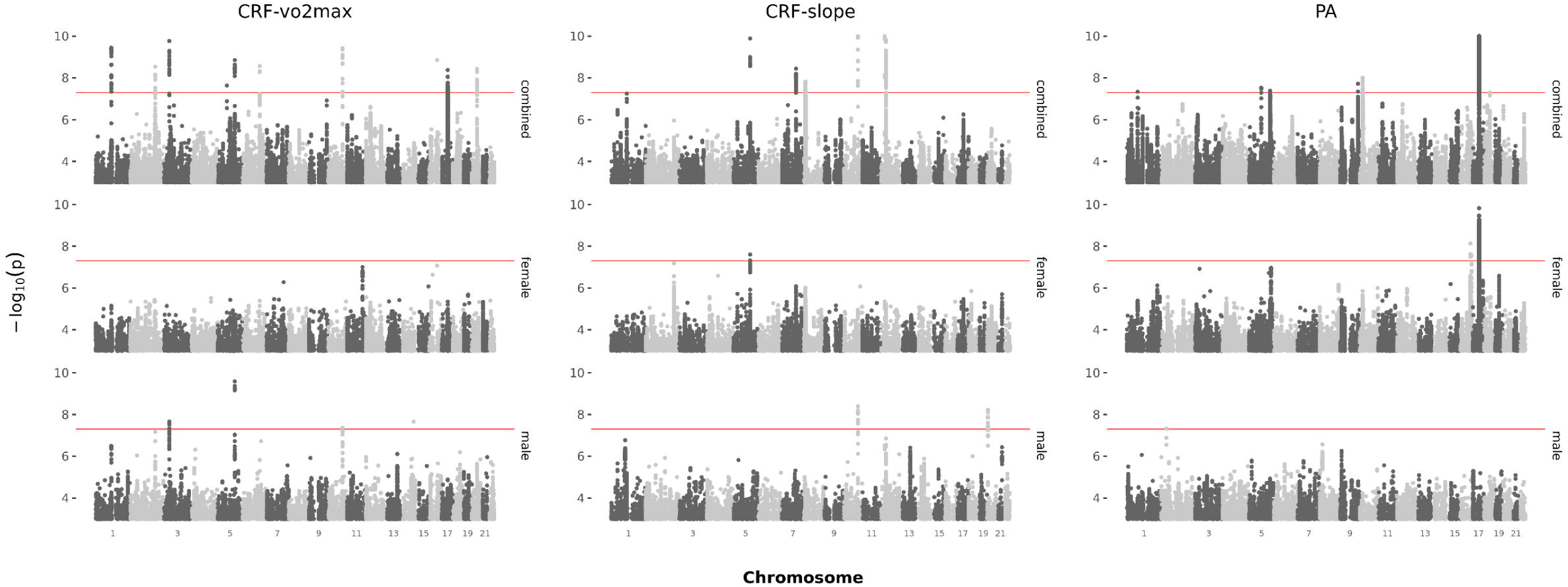
Genome-wide association signal by sex for CRF-vo2max, CRF-slope, and PA. The red line shows genome-wide significance, p=5×10^−8^.

### Sex-combined results

We found twelve significant SNPs for CRF-vo2max and eight independent SNPs for CRF-slope, in the combined sample (**Table 1**). For PA, we found nine significant SNPs. However, three of these were on chromosome 17, two of which are situated within the large 17q21.31 inversion polymorphism (43,624,578 – 44,525,051 MB, human genome build 37 coordinates) and may not reflect independent causal variants (44, 45). We replicated two of the three previously reported PA GWAS significant SNPs (17), rs2696625 (chr17) and rs59499656 (chr18), and tagged the third rs564819152 (chr10) through LD with rs34719019 (chr10), *r*^2^=0.78 in 1000 Genomes EUR (Additional file 1: **Table S7**).

### Sex-specific results

For CRF-vo2max we found four male genome-wide significant SNPs, three of which showed no evidence of heterogeneity and were associated (directly or indirectly through LD) in the combined sample: rs9809798 (*r*^2^=0.79 with rs6801957, chr3), rs111299422 (chr5), rs1006545 (*r*^2^=0.97 with rs11190709, chr10). rs41317306 (chr14) however, showed no evidence for association in females (p=5.1×10^−1^), or the combined sample (p=1.6×10^−4^), and significant effect size heterogeneity (Q=22.13, p<0.001). However, the regional plot for rs41317306 did not show support from correlated SNPs (Additional file 1: **Figure S11**)

For CRF-slope, the one female signal rs111299422 (chr5) was significant in the combined sample, but showed evidence of heterogeneity (Q=8.37, p=3.8×10^−3^). There were two male specific SNPs: rs1006545 (chr10) was indirectly associated in the combined sample (*r*^2^=0.97 with 10:102554618_AT_A) and showed no evidence of heterogeneity. rs1741294 (chr20) showed no evidence of association in females (p=6.0×10^−1^) or the combined sample (p=4.2×10^−4^), and significant effect size heterogeneity (Q=7.96, p=4.8×10^−3^).

For PA, there were four SNPs associated in the female sample, two of which were associated in the combined sample, rs62055696 (chr17) and 17:44262581_A_C (*r*^2^=0.70 with rs62055696, chr17), and showed no evidence of heterogeneity. The two female signals on chromosome 16 signals rs75986475 and rs13329850 were uncorrelated (*r*^2^=0.0005), showed significant heterogeneity (Q=8.51, p=3.5×10^−3^; Q=16.98, p<0.001 respectively), and no evidence of association in males (p=3.3×10^−1^, p=4.5×10^−1^ respectively) or the combined sample (p=1.3×10^−6^, p=7.6×10^−4^ respectively). The male-specific SNP identified in PA, rs78661713 (chr2), showed no evidence for association in females (p=6.1×10^−1^) or the combined sample (p=5.9×10^−4^), and significant heterogeneity (Q=20.63, p<0.001).

### Genes to prioritize for experimental follow-up

For each trait, we report genes identified as both *functional* candidates by TWAS and colocalization analysis, and *positional* candidates (**Table 1**) or gene-based association candidates (Additional file 1: **Table S8**). We then report any additional noteworthy TWAS-significant functional candidates. The functional candidates reported below all passed the TWAS multiple testing threshold (16 for CRF-vo2max, 10 for CRF-slope, and 24 for PA), and are shown in **Figure 4**. Where functional candidates were also implicated in the gene-based association test these are indicated in **Figure 4**. The full list of significant genes from the gene-based test of association are shown in Additional file 1: **Table S8**.

**Figure 4.**
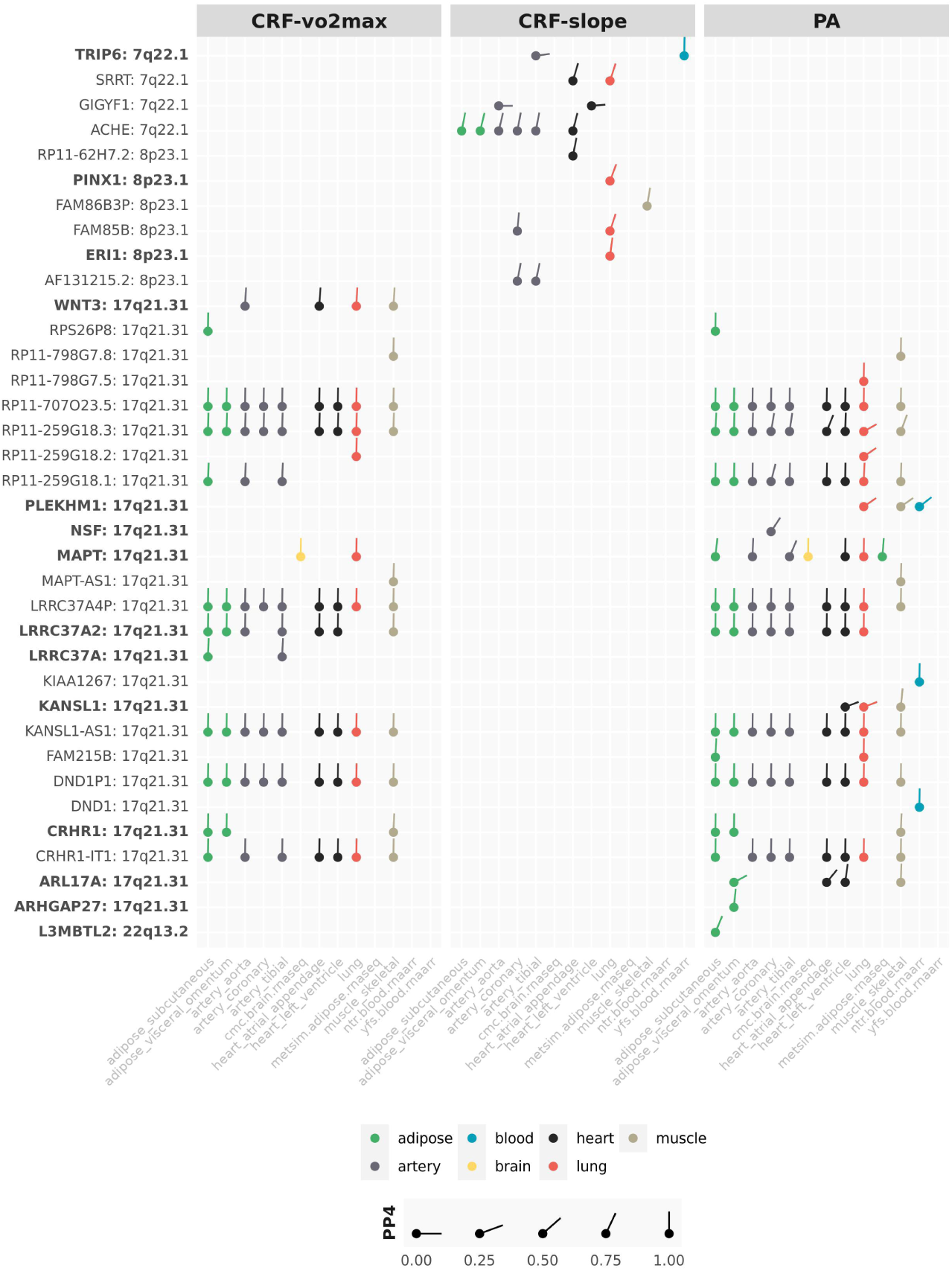
TWAS and colocalization identified genes. Genes listed on the y-axes are significant at the conservative Bonferroni corrected TWAS p-value. On the x-axis is tissue type; category of tissue is represented by colour, colocalization posterior probability 4 (COLOC PP4 – the probability of a shared causal SNP driving the association signal and expression in a reference tissue) represented by line angle (vertical line is a probability of 1.0 that the GWAS-identified SNP is an eQTL for the gene). The complementary TWAS and colocalization approaches provide a priority list of genes for further exploration. Genes in bold were significant in the gene-based test of association.

### CRF-vo2max

All functional candidates were located within cytochrome band 17q21.31, within or near the chromosome 17 inversion. In particular, five functional candidates were identified in the gene-based association and had a high probability of a shared SNP driving both the CRF-vo2max association and expression in the listed tissues (PP4 range=0.94 – 0.99, **Figure 4**): *WNT3* (Wnt family member 3) in aorta, heart (atrial appendage), lung and skeletal muscle; *MAPT* (microtubule associated protein tau) in brain and lung; *LRRC37A2* (leucine rich repeat containing 37A2) in and *LRRC37A* (leucine rich repeat containing 37A) in adipose (subcutaneous and visceral omentum), artery (aorta and tibial), heart (atrial appendage and left ventricle), and skeletal muscle; and *CRHR1* (corticotropin releasing hormone receptor 1) in adipose (subcutaneous and visceral omentum) and skeletal muscle. Notable GWAS catalog associations for this set of genes include disease (atrial fibrillation (46), diabetes mellitus, and coronary artery disease (47), Parkinson’s (48), and Alzheimer’s disease (49)), lung function (FEV1, FVC (50), peak expiratory flow (51)), heart and electrocardiography measures (QRS measures (52), cardiac arrhythmia (53)), and blood phenotypes (systolic blood pressure, eosinophil count (54), haemoglobin level (55), haematocrit (56), red blood cell density (57)).

### CRF-slope

Positional candidate *ACHE* (acetylcholinesterase), implicated by rs4582488 (52.87kb upstream), was significantly associated with expression in adipose (subcutaneous, visceral omentum), artery (aorta, coronary, and tibial), and heart (atrial appendage) tissue. Each of these tissue associations had high probability of a shared SNP driving both the CRF-slope association and expression (PP4 range=0.79 – 0.84). *ACHE* has notable reported associations with resting heart rate (58), heart rate response to exercise and recovery (59), electrocardiography (60), diastolic blood pressure (61), and diabetes mellitus (62).

Three functional candidates were also significant in the gene-based association: *TRIP6* (thyroid hormone receptor interactor 6) with high probability of a shared SNP driving both CRF-slope association and expression in whole blood, and *PINX1* (PIN2 (TERF1) interacting telomerase inhibitor 1) and *ERI1* (exoribonuclease 1) had significant expression in lung, with strong evidence the same variant drives expression and the CRF-slope association (PP4 range=0.71 – 0.98). *TRIP6, PINX1*, and *ERI1* have notably been associated with blood pressure (63), triglyceride levels (64), HDL cholesterol (65), diabetes mellitus (66), and atherosclerosis (67).

Additional TWAS significant functional candidates with high probability of the GWAS variant driving expression included *SRRT* (serrate, RNA receptor molecule) in lung and heart (atrial appendage) tissue, *RP11-62H7*.*2* in heart (atrial appendage), *FAM86B3P* (family with sequence similarity 86 member B3) in skeletal muscle, *FAM85B* (family with sequence similarity 85 member B) in lung and coronary artery, and *AF131215*.*2* in artery (coronary and tibial) (PP4 range=0.74 – 0.92). *SRRT* and *FAM86B3P* are notably associated with heart rate response to exercise and recovery (59), electrocardiography (RR interval) (68), diabetes mellitus (66), and obesity (69).

### PA

All PA functional candidates were on chromosome 17, except *L3MBTL2* on chromosome 22. The number of loci and the best candidates should be interpreted with caution as this is a gene dense region with many copy number variants, and all the chromosome 17 candidates were within or near the large 17q21.3 inversion polymorphism (44, 45). Nonetheless, the positional candidates were supported by the functional analyses.

Ten significant genes from the gene-based association analysis were identified as functional candidates: *PLEKHM1* (plekstrin homology domain containing M1) in lung, skeletal muscle, and peripheral blood with low probability of the GWAS variant driving expression (PP4 range=0.30 – 0.33); *NSF* (N-Ethylmaleimide-Sensitive Factor) in coronary artery (PP4=0.54); *MAPT* in subcutaneous adipose (PP4=0.91), artery (aorta PP4=0.96, tibial PP4=0.67), and brain, left ventricle, lung, adipose (PP4 range=0.93 – 0.99); *LRRC37A2* in adipose (visceral omentum and subcutaneous), artery (aorta, coronary), heart (atrial appendage and left ventricle), lung (all PP4=0.99); *KANSL1* (KAT8 Regulatory NSL Complex Subunit 1) in skeletal muscle (PP4=0.92); *CRHR1* in adipose (visceral omentum and subcutaneous) and skeletal muscle (PP4>=0.98); *ARL17A* (ADP ribosylation factor like GTPase 17A) with high probability of a shared variant driving PA association and gene expression in left ventricle (PP4=0.88) and skeletal muscle (PP4=0.97); *ARHGAP27* (rho GTPase activating protein 27) in visceral omentum (PP4=0.92); and *L3MBTL2* (L3MBTL histone methyl-lysine binding protein 2) in subcutaneous adipose (PP4=0.67).

Previous notable associations with these ten functional candidates include disease (Parkinson’s disease (70), hypertrophic cardio myopathy (71), ischaemic stroke, diabetes mellitus and coronary artery disease (72)) anthropometrics (BMI-adjusted waist hip to ratio, BMI (73)), blood phenotypes (haemoglobin measurement (55), red blood cell density, hematocrit (57), eosinophil count and systolic blood pressure (54)), lung function (FVC (54), FEV1 (51)), and heart and electrocardiography measures (QRS measures (52), cardiac arrhythmia (53)).

**Figure 4** includes the full list of TWAS significant candidates, including those with lower posterior probabilities of a shared variant driving both GWAS trait association and expression.

### Gene set enrichment

We also performed gene set enrichment analyses on the TWAS significant genes for sex-combined CRF-vo2max (16 genes), CRF-slope (10 genes) and PA (24 genes, 23 with recognized Ensembl ID), as well as female PA (22 genes: 21 with recognized Ensembl ID). TWAS analyses of the other sex-specific traits did not yield sufficient gene associations to perform enrichment analyses. Enrichment of TWAS-identified functional candidates in gene sets defined by genes identified in previous GWASs, reported in the GWAS Catalog, are shown in Additional file 1: **Figures S38**–**S40**.

Functional candidates for CRF-vo2max were enriched in GWAS gene sets for multiple diseases including multiple system atrophy, Alzheimer’s disease, Parkinson’s disease, idiopathic pulmonary fibrosis, and primary biliary cirrhosis (FDR-adjusted p range=6.9×10^−3^ – 2.0×10^−5^), as well as lung function (FEV1, FDR-adjusted p=) and blood phenotypes (haematocrit FDR-adjusted p values=2.8×10^−2^, haemoglobin concentration FDR-adjusted p values= 4.0×10^−2^). Enrichment for genes implicated in Alzheimer’s and Parkinson’s disease are noteworthy given there is some evidence of an underlying insulin resistance pathology.

CRF-slope functional genes were enriched in GWAS gene sets for the thrombosis and atherosclerosis risk factor *PAI-1* (plasminogen activator inhibitor-1: FDR-adjusted p=5.0×10^−6^), heart rate response to exercise and recovery (FDR-adjusted p=1.2×10^−5^), and resting heart rate (FDR-adjusted p=1.4×10^−3^)

GWASs genes sets enriched for PA functional candidates included Parkinson’s disease, idiopathic pulmonary fibrosis, Alzheimer’s disease, multiple system atrophy and primary biliary cirrhosis (FDR-adjusted p range=2.7×10^−2^ – 2.0×10^−8^), and mental health traits (e.g., neuroticism, mood instability, and alcohol use disorder). Also noteworthy were enrichment of female PA-associated functional candidates in FEV1 (FDR-adjusted p=4.1×10^−4^) and FVC (FDR-adjusted p=3.1×10^−3^) given their use in the diagnosis of obstructive and restrictive pulmonary diseases.

### Genetic correlations within CRF-vo2max, CRF-slope, and PA

CRF-vo2max, CRF-slope and PA were all polygenic with modest SNP heritabilities of between 0.08 and 0.14 (**Table 2**). There was no observed difference between sexes.

**Table 2.** Heritability of CRF and PA.

|  | CRF-vo2max | CRF-slope | PA |
| --- | --- | --- | --- |
| Combined | 0.10 (0.01) | 0.08 (0.01) | 0.14 (0.01) |
| Male | 0.10 (0.01) | 0.09 (0.02) | 0.13 (0.01) |
| Female | 0.11 (0.02) | 0.10 (0.02) | 0.15 (0.01) |

Note. Values shown are SNP heritabilities and standard errors on the observed scale. CRF-vo2max = maximum volume of oxygen uptake per kg of bodyweight per minute, estimated from workload and heart rate during the cardiorespiratory fitness test, as well as age and weight; CRF-slope= the rate of increase of heart rate during the exercise period of the cycle ramp test; PA = 7-day average physical activity

For the sex-combined between-phenotype genetic correlations, CRF-vo2max and CRF-slope were negatively correlated (*r*_*g*_=-0.66, se=0.04, p=5.4×10^−53^), suggesting genetic variants that contribute to greater fitness are also associated with a lower rate of increase of heart rate during exercise. PA had a moderate positive correlation with CRF-vo2max (*r*_*g*_=0.37, se=0.04, p=1.9×10^−16^), and a modest negative correlation with CRF-slope (*r*_*g*_=-0.14, se=0.06, p=1.0×10^−2^). These two genetic correlations suggest that higher levels of activity have a shared genetic contribution with greater fitness, and with a lower rate of increase of heart rate during exercise. The within-sex between-phenotype genetic correlations were similar to those for the sex-combined results (Additional file 1: **Figure S41**). Within-phenotype between-sex genetic correlations were all near 1.0, reflecting that the overall genomic signal is highly correlated despite a few different loci between sexes (Additional file 1: **Figure S41**).

### Genetic correlations with multiple traits and diseases

A comprehensive analysis of genetic correlations of CRF-vo2max, CRF-slope and PA with a wide array of traits was performed using LD-HUB (**Figure 5** shows the combined-sex result, Additional file 1: **Figures S42**– **S44** show by-sex results). All genetic correlations described below were FDR-significant in the sex-combined sample. Additional file 1: **Table S9** includes the sex-combined, male, and female FDR-significant genetic correlations, as well as heritabilities, associated standard errors, and GWAS study PubMed IDs for all FDR-significant traits and diseases.

**Figure 5.**
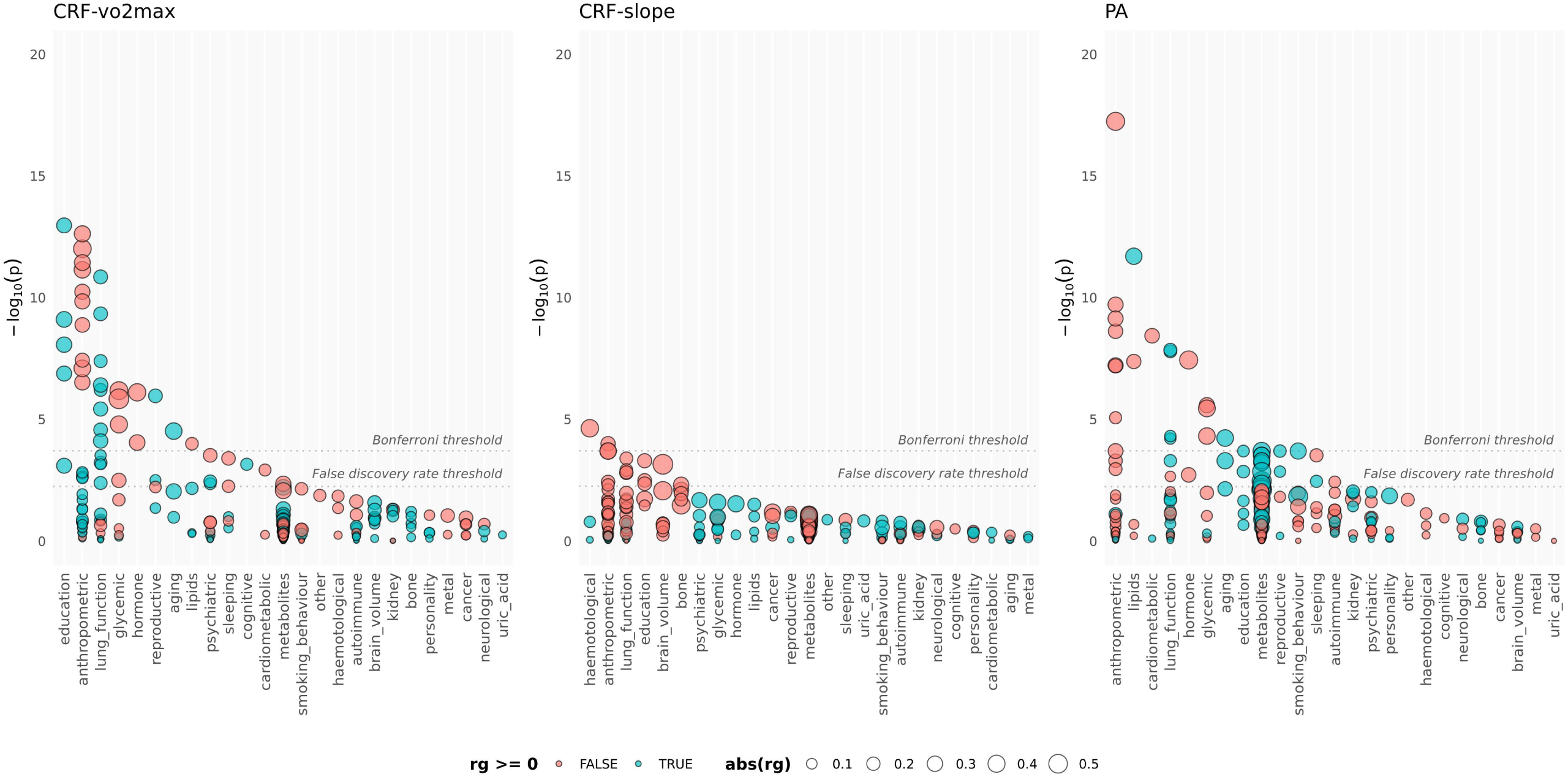
Genetic correlations for combined males and females. red = negative correlations; blue = positive correlations; size = relative size of the genetic correlation; Bonferroni threshold shows p=0.05/total number of correlations per trait; False discovery rate threshold indicates the largest FDR-adjusted p-value < 0.05

### Shared genetic risk: lipids, glycaemic and cardiometabolic traits

Genetic variation associated with lower CRF-vo2max was correlated with features of the typical type-2 diabetes (T2D) profile: higher insulin resistance (HOMA-IR: *r*_*g*_=-0.58, se=0.12) and higher fasting insulin (*r*_*g*_=-0.45, se=0.09), impaired beta-cell function (HOMA-B: *r*_*g*_=-0.40, se=0.09), hyperglycaemia (HbA1c: *r*_*g*_=-0.25, se=0.08) and hypertriglyceridemia (*r*_*g*_=-0.17, se=0.04). In the PA combined sample, as well as genetic correlations with fasting insulin (*r*_*g*_=-0.31, se=0.07), insulin resistance (*r*_*g*_=-0.29, se=0.07), and triglycerides (*r*_*g*_=-0.20, se=0.04), we observed a positive correlation with HDL cholesterol (*r*_*g*_=0.28, se=0.04) and a direct negative correlation with T2D (*r*_*g*_=-0.23, se=0.05). Genetic variation associated with lower levels of physical activity was associated with increased risk of T2D.

Both CRF-vo2max and PA were negatively correlated with coronary artery disease (CRF-vo2max *r*_*g*_=-0.15, se=0.05; PA *r*_*g*_=-0.21, se=0.04). Like T2D, genetic variation associated with lower CRF-vo2max and lower PA was associated with increased risk for coronary artery disease. Additional file 1: **Figure S45** highlights the FDR-significant glycaemic, cardiometabolic, and lipid traits in the combined sample, as well as in males and females separately.

### Shared genetic risk: metabolites, hormones

CRF-vo2max was significantly genetically correlated with glycoprotein acetyls, GlycA (*r*_*g*_=-029, se=0.10), such that genetic variation associated with greater CRF-vo2max was associated with lower levels of the GlycA biomarker. Elevated blood concentrations of GlycA are a strong predictor for long-term risk of morbidity and mortality from diverse diseases (74).

For PA we observed positive FDR-significant genetic correlations with several related HDL metabolites. The largest HDL metabolite correlation with PA was total cholesterol in medium HDL (*r*_*g*_=0.42, se=0.15). Genetic variation associated with greater CRF-vo2max and PA were associated with higher levels of HDL. PA was also significantly correlated with acetate (*r*_*g*_=0.36, se=0.13) and apolipoprotein A-I (*r*_*g*_=0.35, se=0.12) – the major protein component of HDL which enables fat efflux from within cells for transport to the liver for excretion. Additional file 1: **Figure S50** shows the genetic correlations with blood metabolites for the combined sample, and males and females separately.

Circulating levels of the satiety hormone leptin were negatively correlated with CRF-vo2max and PA (CRF-vo2max *r*_*g*_=-0.41, se=0.08; PA *r*_*g*_=-0.37, se=0.07), and for BMI-adjusted leptin (CRF-vo2max *r*_*g*_=-0.32, se=0.08; PA *r*_*g*_=-0.21, se=0.07) (Additional file 1: **Figure S45**). The genetic profile associated with greater levels of fitness and activity is correlated with greater circulating levels of leptin, which inhibits hunger and reduces fat storage in adipocytes.

### Shared genetic risk: body measures

CRF-vo2max was positively genetically correlated with height (*r*_*g*_=0.13, se=0.04), birth weight (*r*_*g*_=0.14, se=0.05) and own birth weight (*r*_*g*_=0.13, se=0.04). All other FDR-significant anthropometric correlations were negative (*r*_*g*_ range=-0.23 – -0.44, se range=0.04 – 0.07): body fat, waist circumference, measures of obesity, BMI, waist-to-hip ratio and hip circumference. CRF-slope had FDR-significant negative genetic correlations with childhood obesity (*r*_*g*_=-0.24, se=0.06) and childhood height (*r*_*g*_=-0.26, se=0.07), BMI (*r*_*g*_=-0.18, se=0.05), birth weight (*r*_*g*_=-0.15, se=0.05) and own birth weight (*r*_*g*_=-0.12, se=0.04). For PA, all FDR-significant body measures including height were negative (*r*_*g*_=-0.12 – -0.37, se=0.03 – 0.06): body fat, waist circumference, measures of obesity, BMI, waist-to-hip ratio and hip circumference, and height. Genetic variation associated with greater CRF-vo2max and higher levels of PA was associated with lower body fat, BMI, obesity, and waist and hip circumferences. Additional file 1: **Figures S46**–**S48** show FDR-significant genetic correlations with anthropometric measures for the combined sample and males and females separately.

### Shared genetic risk: lung function and heart rate

CRF-vo2max was positively associated with lung function: forced vital capacity (FVC: *r*_*g*_=0.22 – 0.26, se=0.03 – 0.06), forced expiratory volume in 1 second (FEV1: *r*_*g*_=0.18 – 0.24, se=0.03 – 0.07), and peak expiratory flow (*r*_*g*_=0.12, se=0.03). CRF-slope was negatively correlated with FVC (*r*_*g*_=-0.13, se=0.04) and peak expiratory flow (*r*_*g*_=-0.14, se=0.04). For PA, we found modest positive genetic correlations with FVC (*r*_*g*_=0.13 – 0.15, se=0.03 – 0.04) and FEV1 (*r*_*g*_=0.10, se=0.02). Genetic variation associated with both fitness and activity is associated with improved lung function.

Only CRF-slope was genetically correlated with resting heart rate (*r*_*g*_=-0.31, se=0.07). Genetic correlation associated with greater acceleration of heart rate during exercise was associated a lower resting heart rate. Additional file 1: **Figure S49** shows genetic correlations with lung function and heart rate, for the combined sample and males and females separately.

### Shared genetic risk: smoking behaviour and longevity

We observed a positive genetic correlation between PA and former vs current smoking status (*r*_*g*_=0.27, se=0.07). A genotype associated with greater levels of activity associated with smoking cessation.

Finally, we found that genetic variation associated with greater CRF-vo2max and PA was also significantly associated with longevity, as measured by father’s age at death (CRF-vo2max *r*_*g*_=0.37, se=0.09; PA *r*_*g*_=0.30, se=0.07) and parents’ ages at death (PA *r*_*g*_=0.28, se=0.08). FDR-significant genetic correlations with smoking behaviour, cancer and longevity reported above are highlighted in Additional file 1: **Figure S51**.

### Shared genetic risk: demographics

CRF-vo2max and PA were positively correlated with years of schooling (CRF-vo2max *r*_*g*_=0.28 – 0.32, se=0.04 – 0.05; PA *r*_*g*_=0.11 – 0.14, se=0.03 – 0.04) and age at birth of first child (CRF-vo2max *r*_*g*_=0.22, se=0.05; PA *r*_*g*_=0.13, se=0.04). CRF-slope had a corresponding negative genetic correlation with years of schooling (*r*_*g*_=-0.15 – -0.17, se=0.05). FDR-significant genetic correlations with education and reproductive phenotypes are highlighted in Additional file 1: **Figure S52**.

## Discussion

Genetic variation explains a modest but significant proportion of the variation in vo2max, rate of increase of heart rate during exercise, and average weekly activity level, for both sexes. Genetic association signals were highly correlated between sexes, with only six SNPs that showed evidence for a sex-specific association (CRF-vo2max: rs1741294 male; CRF-slope: rs111299422 female, rs1741294 male; PA rs75986475, rs13329850 female, rs78661713 male). The two derived fitness traits, CRF-vo2max (vo2max) and CRF-slope (rate of increase of heart rate during exercise), were negatively correlated, phenotypically and genetically. Greater levels of fitness corresponded to a slower rate of increase of heart rate during exercise. CRF-vo2max was positively correlated with PA; greater fitness was associated with higher levels of physical activity.

### Lower CAD and T2D among most fit and active

We found that higher levels of CRF-vo2max and PA were associated with better health outcomes in a dose-dependent way. The frequency of CAD and diabetes mellitus diagnosis was substantially lower among individuals with higher levels of CRF-vo2max and PA, even after controlling for BMI and SES. In this large UK population-based sample, we find the same associations between PA and CRF-vo2max observed in epidemiological and clinical studies of coronary heart disease and cardiovascular events (75) and T2D (76). As has been previously observed (3), there was a noteworthy steeper drop-off in disease occurrence with increased PA and CRF-vo2max among the least fit and active third of the sample. The fitness and activity associations with disease observed in the combined sample were most stark among males. For CRF-vo2max and CRF-slope, disease diagnosis frequency was generally lower as we only included individuals with no contraindications for the standard cycle ramp test protocol. Disease diagnosis association with CRF-slope was noisy, with no clear pattern of association except for a small reduction in CAD and diabetes mellitus. It is not clear why there would be a reduction in disease diagnoses with increasing CRF-slope, given the negative correlation with CRF-vo2max. One possible factor is that among the participants with high CRF-slope levels there is a mix of the least fit samples whose heart rates increased by a larger amount during exercise, participants with lower resting heart rates due to medical conditions and medication (expected to be low given risk category exclusions), and the most fit samples who would have lower resting heart rates.

### Disease associated genes implicated in CRF and PA

Our functional genetic analyses identified genes associated with CRF and PA, with known associations in disease and clinical vital sign related phenotypes. For CRF-vo2max, we found functional candidates associated with expression in heart, artery, lung, brain, adipose, and skeletal muscle. All functional candidates were within 17q21.31, with five genes also identified in the gene-based association analysis (*WNT3, MAPT, LRRC37A2, LRRC37A, CRHR1*). These five genes have previously been associated with GWASs for atrial fibrillation (46), diabetes mellitus, and coronary artery disease (47), Parkinson’s (48), and Alzheimer’s disease (49), lung function (FEV1, FVC (50), peak expiratory flow (51)), heart and electrocardiography measures (52), cardiac arrhythmia (53), and blood phenotypes (systolic blood pressure, eosinophil count (54), haemoglobin level (55), haematocrit (56), red blood cell density (57)). Subsets of CRF-vo2max functional candidates were enriched in the GWAS catalog for multiple system atrophy, Alzheimer’s disease, Parkinson’s disease, idiopathic pulmonary fibrosis, primary biliary cirrhosis, FEV1, haematocrit, and haemoglobin concentration.

For CRF-slope, functional candidates fell within one of two cytogenic regions, 7q21.1 and 8p23.1. *ACHE* within 7q21.1, also identified as a positional candidate 52.87kb upstream of rs4582488, was significantly associated with expression in adipose, artery and heart tissue, and has previously been associated with resting heart rate (58), heart rate response to exercise and recovery (59), electrocardiography (60), diastolic blood pressure (61), and diabetes mellitus (62). The gene-based association identified genes (*TRIP6, PINX1*, and *ERI1*) were significantly associated with expression change in whole blood and lung, and have previously been associated with blood pressure (63), triglyceride levels (64), HDL cholesterol (65), diabetes mellitus (66), and atherosclerosis (67). Other promising functional candidates (*SRRT, RP11-62H7*.*2, FAM86B3P, FAM85B*, and *AF131215*.*2*) were associated with expression in lung, heart, skeletal muscle, and coronary and tibial artery, and have been associated in previous GWASs with heart rate response to exercise and recovery (59), electrocardiography (68), diabetes mellitus (66), and obesity (69). We found subsets of the CRF-slope functional candidates were enriched in GWAS gene sets for the thrombosis and atherosclerosis risk factor *PAI-1*, heart rate response to exercise and recovery, and resting heart rate.

For PA, the functional candidates on 17q21.31 (*MAPT, LRRC37A2, LRRC37A, KANSL1, CRHR1, ARL17A*, and *ARHGAP27*) and 22q13.2 (*L3MBTL2*) with a high probability of the same variant driving PA association and expression in adipose, brain, heart, lung and muscle, have previously been associated with disease (Parkinson’s disease (70), hypertrophic cardio myopathy (71), ischaemic stroke, diabetes mellitus and coronary artery disease (72)) anthropometrics (BMI-adjusted waist hip to ratio, BMI (73)), blood phenotypes (haemoglobin measurement (55), red blood cell density, hematocrit (57), eosinophil count and systolic blood pressure (54)), lung function (FVC (54), FEV1 (51)), and heart and electrocardiography measures (QRS measures (52), cardiac arrhythmia (53)). We found PA functional candidates enriched in Parkinson’s and Alzheimer’s disease, noteworthy given their underlying insulin resistance pathology (although the insulin resistance link is controversial for Alzheimer’s disease), idiopathic pulmonary fibrosis, multiple system atrophy and primary biliary cirrhosis. Functional candidates from the female PA GWAS were also enriched in the lung function phenotypes FEV1 and FVC.

### Genetic correlations link CRF and PA with disease

Genetic correlations complement epidemiological and clinical observations. Because inherited genetic risk cannot be due to confounding, genetic correlations limit the number of potential confounders linked through genotype. The shared genetic architecture of both CRF-vo2max and PA clearly implicated the lifestyle-related chronic diseases obesity, T2D, and CAD, and their body composition and metabolic risk factors. CRF-vo2max and PA were negatively genetically correlated with obesity, body fat, body mass index, and waist-to-hip circumference, triglycerides, the satiety hormone leptin, fasting insulin, insulin resistance, and T2D (significant only for PA), and CAD. These negative genetic correlations corroborate the epidemiological associations, previous research findings, and clinical observations of CRF and PA and body composition (77, 78), lipid profile (79, 80), adipocytokines (81), fasting insulin and insulin resistance (82-86) and leptin (76), T2D (87-89), and CAD (90). Likewise, the positive genetic correlations are consistent with previous observations of positive relationships between CRF and PA, and HDL cholesterol (80, 91) and longevity (92-94).

CRF-vo2max was negatively genetically correlated with blood levels of glycoprotein acetyls (GlycA). GlycA is associated with both acute and chronic inflammation and is a strong predictor for long-term risk of morbidity and mortality from a wide range of diseases (74).

Additionally, CRF-vo2max and PA had positive genetic correlations with age at birth of first child and level of education, respectively a correlate and component of SES. CRF-slope showed a corresponding negative correlation with years of education. This is consistent with population-based observations of a socioeconomic gradient in PA and CRF: people living in higher socioeconomic status areas tend to be more active and more fit, and from a public health perspective particularly important as “reduced PA and fitness levels may contribute to social inequalities in health” (95).

We note several limitations. The gold standard for CRF measurement involves exercise to volitional exhaustion with measurement of gas exchange (uptake of oxygen, elimination of carbon dioxide). A submaximal CRF test with extrapolation of VO2max scores is however a much more practical solution, especially for the collection of a large sample in the age ranges studied. The objective PA monitoring only captured a 7-day window, effectively a snapshot of participant behaviour, and we do not know if this reflects their habitual behaviour. We also used raw acceleration and did not attempt to tease apart PA intensity, recognized as important for health outcomes. All genetic associations, especially the sex-specific SNPs identified, will require replication in an independent sample. In our genetic correlation analyses, some of the largest correlations were observed with blood metabolites but these were based on relatively small GWAS reference samples (metabolites N=24,925 compared to body fat N=100,716) and many metabolite correlations failed to pass the multiple testing threshold. As well as this sample size (or power of each discovery GWAS) limitation, genetic correlations cannot resolve reverse causation. Another limitation is correlated lifestyle risk factors not considered, in particular eating behaviours and nutrition. Eating habits, diet, and exercise behaviours tend to go hand in hand, in large part influenced by the environment, e.g., sedentary time spent watching television and eating high-fat foods (96, 97). A holistic approach to health – addressing eating habits and including an effective dietary intervention (98), alongside prescription PA and monitoring of CRF – may be the most effective practical approach, especially in already overweight individuals given the possibility that there exists a bi-directional relationship between adiposity and PA (99). Despite the complex interplay of eating habits, nutrition, environment, and PA, CRF as a regularly measured vital sign is an objective and easily measured sentinel for health.

## Conclusions

In summary, we found strong evidence that genetic variants associated with CRF and PA also influenced genetic expression in a small set of genes in heart, artery, brain, lung, muscle, and adipose tissue. These functionally relevant genes were enriched among genes known to be associated with CAD, T2D and Alzheimer’s disease – three of the top 10 causes of death in high-income countries – as well as Parkinson’s, pulmonary fibrosis, and blood pressure, heart rate, and respiratory phenotypes. Finally, genetic correlation related lower fitness and activity with several disease risk factors including greater waist-to-hip ratio, BMI, and obesity; with a typical T2D profile including higher insulin resistance, higher fasting glucose, impaired beta-cell function, hyperglycaemia, hypertriglyceridemia; increased risk for CAD and T2D; and shorter lifespan. Genetics supports three decades of evidence for the inclusion of CRF as a clinical vital sign (3). CRF is a window onto general health and wellbeing, and physical activity is the primary means of improving CRF. In fact, to meet the WHO policy of reduction in sedentary lifestyles that carry a heavy cost to personal and public health, “policies to increase population levels of physical activity need to be prioritised and scaled up urgently” (9). PA too is “vital”; we think both measures provide important information and ideally should be collected in tandem if feasible. Given the global health burden of non-communicable disease related to lifestyle choices, as well as dietary advice, regular measurement of CRF as a marker of health and routine prescription of PA is imperative.

## Data Availability

All data are available through an application to the UK Biobank. Scripts for derived variables available upon request.

## List of abbreviations

CRF: cardiorespiratory fitness
PA: physical activity
AHA: American Heart Association
UKB: UK Biobank
CRF-vo2max: an estimate of the participants’ maximum volume of oxygen uptake, per kilogram of body weight, per minute
CRF-slope: an estimate of the rate of increase of heart rate during exercise
GWAS: genome-wide association study
CAD: coronary artery disease/ ischaemic heart disease
T2D: type 2 diabetes
*h*^*2*^: heritability
*h*^*2*^_SNP_: SNP heritability
ECG: electrocardiograph
BMI: body mass index
HR: heart rate
WL: workload
WHO: world health organization
ICD-10: International Classification of Diseases revision 10
SES: socioeconomic status
TWAS: transcriptome-wide association
FDR: false-discovery rate
eQTL: expression quantitative trait locus
PP4: posterior probability 4

## Declarations

### Ethics approval and consent to participate

Ethics approval for the UK Biobank study was obtained from the North West Centre for Research Ethics Committee (11/NW/0382); participants provided electronic signed consent at recruitment. This research has been conducted under application number 13427.

### Consent for publication

Not applicable.

### Availability of data and materials

The data that support the findings of this study are available upon application to the UK Biobank (UKB). The UKB database is globally accessible to approved researchers.

### Competing interests

The authors declare that they have no competing interests.

### Funding

This research was funded by the National Institute for Health Research (NIHR) Biomedical Research Centre (BRC) South London and Maudsley, and Guy’s and St Thomas’, NHS Foundation Trusts, and King’s College London. The views expressed are those of the authors and not necessarily those of the NHS, the NIHR or the Department of Health and Social Care. CML receives funding from a UK Medical Research Council (MRC) grant (MR/N015746/1).

### Authors’ contributions

KBH, MH, CML conceived and designed the study. KBH analysed the data. KBH, EP, MT, KPG, MH, JRIC, CML interpreted the results and wrote the manuscript. All authors read and approved the final manuscript.

## Acknowledgements

We would like to acknowledge the UKB participants for their contributions.

## Additional files

### Additional file 1

Additional file 1.docx includes supplementary Methods (genetic data quality control, GWAS considerations, alternate genetic association models, and post GWAS details), Results (including alternate association models, GWAS post-processing), Tables S1 – S9, and Figures S1 – S52.

## Notes

### Competing Interest Statement

The authors have declared no competing interest.

### Author Declarations

Ethics approval for the UK Biobank study was obtained from the North West Centre for Research Ethics Committee (11/NW/0382); participants provided electronic signed consent at recruitment.

### Summary of Updates

We have increased the number of participants by changing our exclusion criteria. We included samples where PA was not calibrated on their own data, excluded samples with overall PA >100mg (not 3SD outliers), and did not exclude BMI outliers. This increased the PA sample size from 75,799 to 89,683 (an 18% increase). For CRF, by including "minimal", "small" and "medium" CRF risk categories as done by Kim et al. (2018), widening the range of allowable exercise test cadence (35-125 RPM), and not excluding BMI outliers, we increased the CRF sample size from 38,160 to 70,783 (an 85% increase). We also derived CRF-vo2max differently here (the formula used initially is usually used in a steady-state fitness test): we fitted a linear model to the ramp stages in the "minimal" and "small" categories, a linear model to the constant load in the "medium" category (which did not include a ramped work rate), and for all three categories predicted relative workload (or power, in watts/ kg) at the age estimated maximum heart rate as a proxy for vo2max. This is similar to the approach taken by Kim et al. (2018). We then performed a mixed model GWAS of each phenotype using BOLT-LMM (replacing the previous logistic regression models).

