## Supplementary material for "The genetic case for cardiorespiratory fitness as a clinical vital sign and the routine prescription of physical activity in healthcare": Table 1

**Table 1. GWAS significant (p < 5x10^-8^) SNP associations**

| **SEX** | **CHR** | **BP** | **SNP** | **LOCUS** | **A1** | **A0** | **A1 FREQ** | **BETA** | **SE** | **P** | **P (other sex)** |
| --- | --- | --- | --- | --- | --- | --- | --- | --- | --- | --- | --- |
| ***CRF-vo2max*** |  |  |  |  |  |  |  |  |  |  |  |
| combined | 1 | 112592672 | rs269071 | intergenic^1^ | A | G | 0.63 | 0.031 | 0.005 | 3.6x10^-10^ |  |
| combined | 2 | 179747068 | rs142556838 | *CCDC141* | C | T | 0.91 | 0.049 | 0.008 | 2.9x10^-9^ |  |
| combined | 2 | 179839888 | rs10497529 | *CCDC141* | G | A | 0.96 | 0.085 | 0.013 | 1.0x10^-11^ |  |
| combined | 3 | 38767315 | rs6801957 | *SCN10A* | T | C | 0.41 | 0.030 | 0.005 | 1.7x10^-10^ |  |
| combined | 5 | 65264090 | rs251295 | *ERBB2IP* | A | G | 0.59 | -0.027 | 0.005 | 2.3x10^-8^ |  |
| combined | 5 | 121868475 | rs111299422 | intergenic^2^ | T | TA | 0.69 | -0.038 | 0.005 | 6.5x10^-14^ |  |
| combined | 6 | 122089704 | rs58730006 | intergenic^3^ | A | AT | 0.90 | 0.047 | 0.008 | 2.7x10^-9^ |  |
| combined | 10 | 102552663 | rs11190709 | *PAX2* | G | A | 0.11 | -0.046 | 0.007 | 3.8x10^-10^ |  |
| combined | 16 | 56803199 | rs78291913 | *NUP93* | C | T | 0.99 | -0.143 | 0.024 | 1.4x10^-9^ |  |
| combined | 17 | 43668512 | rs527325496 | intergenic^4^ | C | CAAA | 0.81 | -0.036 | 0.006 | 4.2x10^-9^ |  |
| combined | 17 | 44335579 | rs139077859 | *LOC644172* | G | A | 0.79 | -0.033 | 0.006 | 1.7x10^-8^ |  |
| combined | 20 | 36849088 | rs4811602 | *KIAA1755* | G | A | 0.53 | -0.028 | 0.005 | 3.7x10^-9^ |  |
| male^a^ | 3 | 38773805 | rs9809798 | *SCN10A* | A | C | 0.47 | 0.040 | 0.007 | 2.2x10^-8^ | 6.3x10^-3^ |
| male^b^ | 5 | 121868475 | rs111299422 | intergenic^2^ | T | TA | 0.69 | -0.050 | 0.008 | 2.6x10^-10^ | 3.3x10^-5^ |
| male^c^ | 10 | 102553647 | rs1006545 | *PAX2* | G | T | 0.11 | -0.062 | 0.011 | 4.3x10^-8^ | 2.7x10^-3^ |
| male^d^ | 14 | 96864374 | rs41317306 | *AK7* | T | G | 0.98 | -0.137 | 0.024 | 2.2x10^-8^ | 5.1x10^-1^ |
| ***CRF-slope*** |  |  |  |  |  |  |  |  |  |  |  |
| combined | 5 | 121868475 | rs111299422 | intergenic^2^ | T | TA | 0.69 | 0.007 | 0.001 | 1.3x10^-10^ |  |
| combined | 7 | 100546458 | rs4582488 | integenic^5^ | G | T | 0.74 | 0.007 | 0.001 | 3.6x10^-9^ |  |
| combined | 8 | 8317817 | rs2921060 | intergenic^6^ | A | C | 0.55 | 0.006 | 0.001 | 1.6x10^-8^ |  |
| combined | 8 | 10822431 | rs35792458 | *XKR6* | G | C | 0.56 | -0.006 | 0.001 | 1.5x10^-8^ |  |
| combined | 8 | 11423072 | rs12541800 | intergenic^7^ | A | G | 0.52 | 0.006 | 0.001 | 4.7x10^-8^ |  |
| combined | 10 | 102554618 | 10:102554618_AT_A | *PAX2* | AT | A | 0.11 | 0.012 | 0.002 | 1.5x10^-13^ |  |
| combined | 12 | 24758480 | rs4963772 | intergenic^8^ | G | A | 0.85 | -0.009 | 0.001 | 5.8x10^-11^ |  |
| combined | 12 | 33633599 | rs7303356 | intergenic^9^ | G | C | 0.49 | -0.006 | 0.001 | 1.5x10^-10^ |  |
| female^e^ | 5 | 121868475 | rs111299422 | intergenic^2^ | T | TA | 0.69 | 0.010 | 0.002 | 2.5x10^-8^ | 1.1x10^-3^ |
| male^f^ | 10 | 102553647 | rs1006545 | *PAX2* | G | T | 0.11 | 0.012 | 0.002 | 6.0x10^-12^ | 2.0x10^-5^ |
| male^g^ | 20 | 4131944 | rs1741294 | *SMOX* | C | G | 0.96 | 0.016 | 0.003 | 6.0x10^-9^ | 6.0x10^-1^ |
| ***PA*** |  |  |  |  |  |  |  |  |  |  |  |
| combined | 1 | 78450517 | rs34517439 | intergenic^10^ | C | A | 0.88 | 0.316 | 0.058 | 4.6x10^-8^ |  |
| combined | 5 | 87942506 | rs10067451 | *LINC00461* | G | A | 0.89 | 0.333 | 0.060 | 3.0x10^-8^ |  |
| combined | 5 | 152238114 | 5:152238114* | *LOC101927134* | TTTTTTTTTTTTC | T | 0.71 | 0.230 | 0.042 | 4.1x10^-8^ |  |
| combined | 9 | 128195657 | rs1268539 | intergenic^11^ | C | A | 0.58 | -0.214 | 0.038 | 1.9x10^-8^ |  |
| combined | 10 | 21885577 | rs34719019 | *MLLT10* | A | T | 0.73 | 0.242 | 0.042 | 9.7x10^-9^ |  |
| combined | 17 | 43758125 | rs62055696 | *CRHR1* | A | G | 0.78 | -0.309 | 0.046 | 1.3x10^-11^ |  |
| combined | 17 | 44326864 | rs2696625 | intergenic^12^ | A | G | 0.77 | -0.310 | 0.045 | 4.8x10^-12^ |  |
| combined | 17 | 44828931 | rs199533 | *NSF* | G | A | 0.79 | -0.250 | 0.046 | 4.0x10^-8^ |  |
| combined | 18 | 40768309 | rs59499656 | intergenic^13^ | A | T | 0.66 | -0.215 | 0.039 | 5.0x10^-8^ |  |
| female^h^ | 16 | 71464058 | rs75986475 | intergenic^14^ | C | G | 0.88 | -0.436 | 0.076 | 7.4x10^-9^ | 3.3x10^-1^ |
| female^i^ | 16 | 80784797 | rs13329850 | *CDYL2* | C | G | 0.74 | -0.308 | 0.055 | 2.8x10^-8^ | 4.5x10^-1^ |
| female^j^ | 17 | 43758125 | rs62055696 | *CRHR1* | A | G | 0.78 | -0.358 | 0.059 | 1.3x10^-9^ | 2.4x10^-4^ |
| female^k^ | 17 | 44262581 | 17:44262581_A_C | *KANSL1* | A | C | 0.83 | -0.450 | 0.070 | 1.5x10^-10^ | 8.2x10^-3^ |
| male^l^ | 2 | 36592600 | rs78661713 | *CRIM1* | G | A | 0.94 | -0.700 | 0.128 | 4.8x10^-8^ | 6.1x10^-1^ |

Note. These are independent SNP associations determined by p-value informed LD clumping (SNPs correlated 0.2 or greater in a 500 kb). 2 CRF-slope associated SNPs (rs587631263, chr7; rs10623635, chr10) and 16 PA associated SNPs (rs10828247, chr10; 15 chr17 SNPs listed in supplementary material) not available in LD reference data. LOCUS = nearest gene; A1 = effect allele; A0 = reference allele; A1 FREQ = effect allele frequency; BETA = effect size (from BOLT-LMM approximation to infinitesimal mixed model); SE = standard error of the effect size

* 5:152238114_TTTTTTTTTTTTC_T

^1^ *LOC643355* (+51.2kb), *CTTNBP2NL* (-346.1kb); ^2^ *MGC32805* (+53.7kb), *LOC101927379* (-96.2kb); ^3^ *GJA1* (+318.8kb), *HSF2* (-631kb); ^4^ *LOC644172* (-9.0kb), *LRRC37A4P* (+70.62kb); ^5^ *MUC3A* (-0.593kb), *ACHE* (+52.87kb); ^6^ *SGK223* (+78.47kb), *CLDN23* (-241.8kb); ^7^ *BLK* (+0.964kb), *LINC00208* (-10.97kb); ^8^ *LINC00477* (+21.38kb), *BCAT1* (-204.5kb); ^9^ *SYT10* (+40.84kb), *ALG10* (-541.6kb); ^10^ *FUBP1* (+5.74kb), DNAJB4 (-20.12kb); ^11^ *MAPKAP1* (-4.015kb), *GAPVD1* (+68.37kb); ^12^ *LOC644172* (+3.71kb), *LRRC37A* (-45.63kb); ^13^ *RIT2* (+72.65kb), *SYT4* (-79.55kb); ^14^ *ZNF23* (-17.44kb), *CALB2* (+39.72kb)

^a^ female BETA = 0.0163, SE = 0.0060: Q = 6.58, p = 1.0x10^-2^; *I*^2^ = 84.8%, 95% CI = 38.0% – 96.3% (ns_CRF_ = not significant at 0.05/7, for 7 CRF SNPs tested)

^b^ female BETA = -0.0271, SE = 0.0065: Q = 4.86, p = 2.8x10^-2^; *I*^2^ = 79.4%, 95% CI = 11.1% – 95.2% (ns_CRF_)

^c^ female BETA = -0.0281, SE = 0.0094: Q = 5.34, p = 2.1x10^-2^; *I*^2^ = 81.3%, 95% CI = 20.3% – 95.6% (ns_CRF_)

^d^ female BETA = 0.0136, SE = 0.0206: Q = 22.13, p < 0.001; *I*^2^ = 95.5%, 95% CI = 86.8% – 98.5%

^e^ male BETA = 0.0038, SE = 0.0012: Q = 8.37, p = 3.8x10^-3^; *I*^2^ = 88.1%, 95% CI = 54.1% – 96.9%

^f^ female BETA = 0.0111, SE = 0.0026: Q = 0.04, p = 8.3x10^-1^; *I*^2^ = 0.0% (ns_CRF_)

^g^ female BETA = 0.0021, SE = 0.0040: Q = 7.96, p = 4.8x10^-3^; *I*^2^ = 87.4%, 95% CI = 51.1% – 96.8%

^h^ male BETA = -0.0893, SE = 0.0916: Q = 8.51, p = 3.5x10^-3^; *I*^2^ = 88.3%, 95% CI = 55.0% – 96.9%

^i^ male BETA = 0.0503, SE = 0.0670: Q = 16.98, p < 0.001; *I*^2^ = 94.1%, 95% CI = 81.4% – 98.1%

^j^ male BETA = -0.2636, SE = 0.0718: Q = 1.03, p = 3.1x10^-1^; *I*^2^ = 3.0% (ns_PA_ = not significant at 0.05/5, for 5 PA SNPs tested)

^k^ male BETA = -0.2258, SE = 0.0855: Q = 4.09, p = 4.3x10^-2^; *I*^2^ = 75.6%, 95% CI = 0.0% – 94.5% (ns_PA_)

^l^ female BETA = 0.0542, SE = 0.1057: Q = 20.63, p < 0.001; *I*^2^ = 95.2%, 95% CI = 85.5% – 98.4%
