## Supplementary material for "The genetic case for cardiorespiratory fitness as a clinical vital sign and the routine prescription of physical activity in healthcare"

**Table of supplementary tables**

**Table S1**. Derivation of final GWAS sample

**Table S2**. CRF cycle ramp test risk categories

**Table S3**. Top 10 global cause of death in high-income countries, 2016

**Table S4**. Distribution of CRF-vo2max, CRF-slope, and PA

**Table S5**. Mean trait value by smoking and alcohol status

**Table S6**. Linear models of trait ~ age + sex + smoking status + alcohol status

**Table S7**. PA association statistics for rs564819152, chromosome 10

**Table S8**. Gene-based association test significant genes

**Table S9**. Genetic correlation (FDR-significant)

**Table of supplementary figures**

**Figure S1**. Raw heart rate and cadence

**Figure S2**. Phenotypic correlations between CRF and PA, and age, BMI, and SES

**Figure S3–S37**. Regional association plots

**Figure S38–S40**. Functional candidates enriched in GWAS catalog traits

**Figure S41**. Genetic correlations between CRF and PA by sex

**Figure S42**. Genetic correlations between CRF-vo2max and LD Hub traits by sex

**Figure S43**. Genetic correlations between CRF-slope and LD Hub traits by sex

**Figure S44**. Genetic correlations between PA and LD Hub traits by sex

**Figure S45**. Genetic correlations with glycaemic, cardiometabolic, and lipid traits

**Figure S46**. Genetic correlations with anthropometric measures (CRF-vo2max).

**Figure S47**. Genetic correlations with anthropometric measures (CRF-slope).

**Figure S48**. Genetic correlations with anthropometric measures (PA).

**Figure S49**. Genetic correlations with lung function and heart rate traits

**Figure S50**. Genetic correlation with blood metabolites

**Figure S51**. Genetic correlations with smoking behaviour, cancer and longevity

**Figure S52**. Genetic correlations with education and reproductive measures

**Supplementary Methods**

**Genome-wide association**

**QC.** Using the supplied genotype metadata described in UKB Resource 531 (https://biobank.ctsu.ox.ac.uk/crystal/refer.cgi?id=531), we excluded individuals with mismatching reported and genotype-inferred gender, with excess relatives, with putative sex chromosome aneuploidy, samples identified as heterozygosity or missingness outliers.

From the full imputed set of SNPs, we excluded SNPs with imputation info score < 0.7 (--bgenMinINFO=0.7), minor allele frequency < 0.01 (--bgenMinMAF=0.01), and filtered individuals and SNPs with missing rates exceeding 0.1 (i.e., missing greater than 10%). Hardy-Weinberg equilibrium (HWE) filtering (p<1x10^-50^) was performed as an association post-processing step (BOLT-LMM does not include HWE filtering).

We calculated the first 10 principal components for the subset of participants with CRF and PA traits separately, to control for population structure in the genetic association analyses. For the principal components analyses, we further pruned the genetic variants as follows: genotyped SNPs only, pruned for long- and short-range LD. Long-range LD-pruning removed the long-range high-LD regions reported by Price et al. ^1^ (lifted over to hg19 with PyLiftover, https://www.encodeproject.org/software/pyliftover/), as well as the chromosome 17 inversion (42 – 46 Mb). Short-range LD-pruning was done with PLINK’s --indep-pairwise flag, window size 10,000 kb, step size 50, *r*^2^ threshold 0.05.­­­­­

**Association.** We performed sex-specific and sex-combined genome-wide linear mixed model association analyses of our three phenotypes (CRF-vo2max, CRF-slope, PA), with imputed genotype data in PLINK 2.0. Association analyses were performed on the residuals of the trait of interest regressed on the covariates, for four different models:

m1:	age + sex (for sex combined analyses) + array + centre + trend (CRF only) + category (CRF only)

m2:	m1 + age^2^

m3:	m2 + BMI

m4:	m3 + smoking status + alcohol status

We report the results of model m2 in the main paper and include manhattan plots for the alternate models m1, m3, and m4 in this supplementary material. We used the standard p = 5 x 10^-08^ as our genome-wide significant threshold. We used a p-value informed LD-clumping post-processing strategy (PLINK 1.9 --clump with parameters --clump-p1 5e-8 --clump-kb 500 --clump-r2 0.2) to derive a list of independent associations.

For all SNPs identified in the male- and female-specific association analyses, we calculated variation in effect size attributable to heterogeneity beyond chance, *I^2^*, and performed a formal test of effect size difference with Cochran’s Q-statistic which distributes as χ^2^ (with 1 df, for the male–female test).

**Post GWAS.** From the summary statistics of our GWASs for CRF-vo2max, CRF-slope and PA, we (1) performed transcriptome-wide association studies (TWASs) and colocalization with expression in selected tissues: brain, adipose, blood, skeletal muscle, lung, artery, and heart analysis using FUSION which includes an interface to COLOC ^2,3^; (2) tested gene-set enrichment in the GWAS Catalog (FUMA) ^4,5^; (3) estimated SNP heritabilities for our three phenotypes (LDSC) and derived genetic correlations with a broad cross-section of traits (LD Hub) ^6-8^.

TWAS is a test of significant association between tissue-specific gene expression and a GWAS trait. Colocalization is a complementary approach that attempts to discover genes whose expression in a particular tissue is regulated by the same variants driving GWAS trait association signals. For TWAS in FUSION we used GWAS summary statistics for our CRF and PA phenotypes and pre-computed functional weights for each gene based on expression in reference data (http://gusevlab.org/projects/fusion/): Netherlands Twin Register, NTR (peripheral blood), Young Finns Study, YFS (whole blood), ComonMind Consortium, CMC (brain), Metabolic Syndrome in Men, METSIM (adipose), Genotype Tissue Expression, GTEx (adipose subcutaneous, adipose visceral omentum, muscle skeletal, lung, artery aorta, artery coronary, artery tibial, heart atrial appendage, heart left ventricle). We selected TWAS-significant genes and followed up those with a high COLOC posterior probability of a shared causal SNP driving the association signal and expression in a particular reference tissue (PP4). We used a conservative Bonferroni-corrected p-value for TWAS significance, correcting for all genes tested in all tissues, p = 0.05 / 75,486 = 6.6 x 10^-07^.

We performed gene-set enrichment analyses of the TWAS significant genes with FUMA’s hypergeometric test, among sets defined by genes identified in previous GWASs reported in the GWAS Catalog, against a background of 20,260 protein coding genes. We also calculated gene-based association statistics which FUMA derives by combining GWAS summary statistics for SNPs within a gene and incorporating LD from a reference population ^4^.

We computed SNP heritabilities from CRF-vo2max, CRF-slope, and PA summary association statistics in HapMap3 European reference data using a subset of SNPs considered well-imputed in most studies, and genetic correlations between our three phenotypes and 702 traits and diseases from LD Hub.

**Supplementary Results**

**Genetic associations**

*Alternate model fit*

Reported in the main paper are the genetic association model m2 association results and summary statistics used for post GWAS analyses. Alternative model m3 including BMI did explain additional variance; m4 including smoking and alcohol did not. However, neither model changed the pattern of association (cf. **Figure 2**).

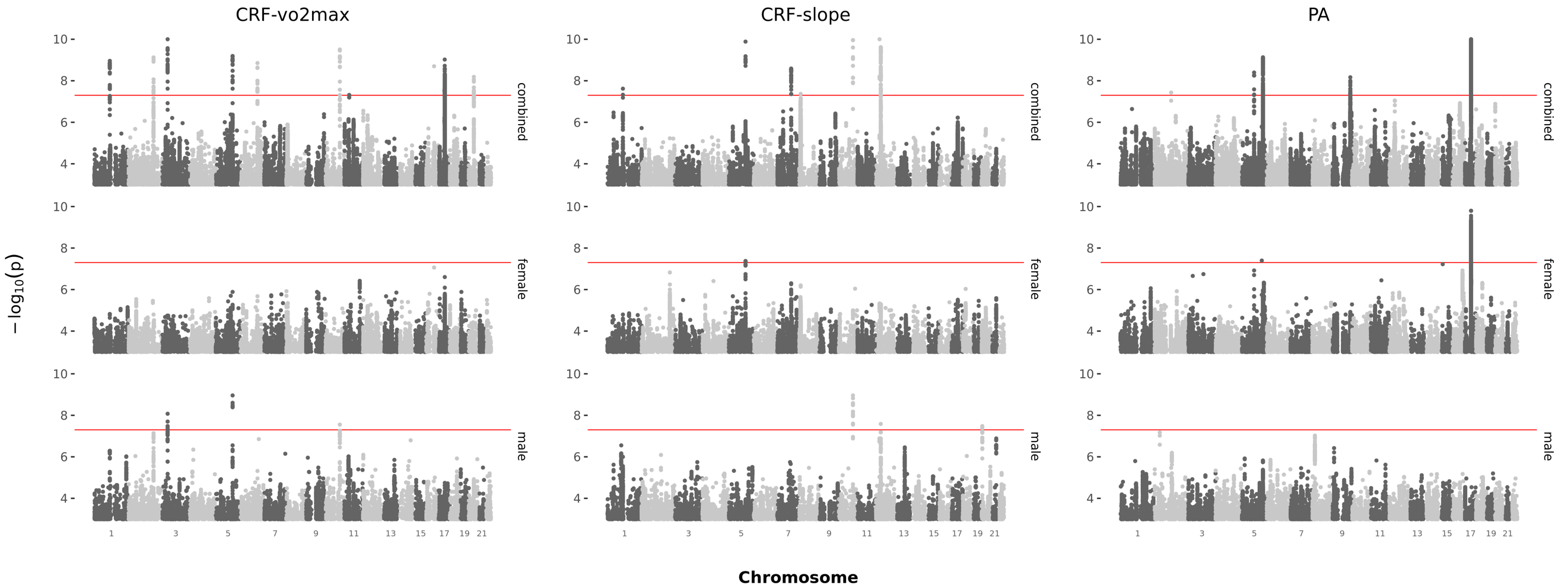

**m3:** m3 – m2 delta R2 range across all traits and sexes = 0.00303987 – 0.00797246

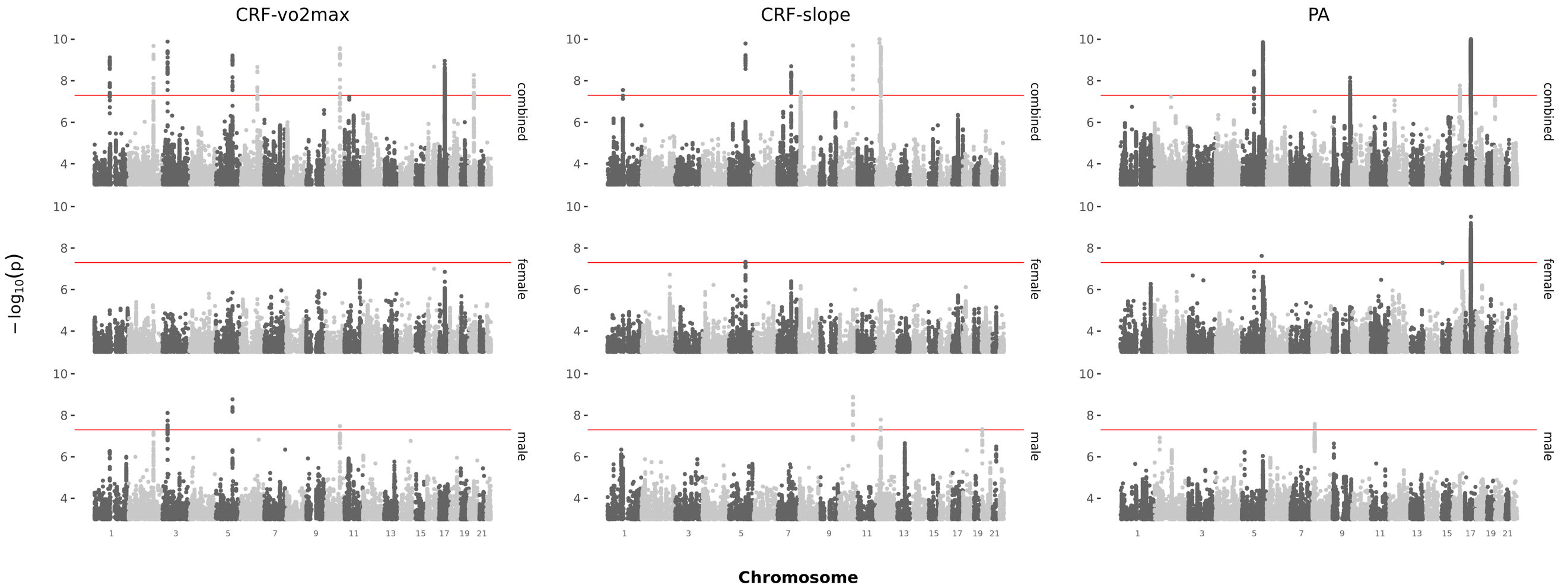

**m4**: m4 – m3 delta R2 range across all traits and sexes = 0.00025429 – 0.00483781

*GWAS postprocessing*

Additional PA-associated chromosome 17 SNPs not in LD reference data and not included in PLINK –clump: rs111443054, rs559943616, rs872972, rs35331519, rs112904481, rs56303031, 17:43897026_TGGAG_T, rs10491140, rs9901937, rs62063678, rs111990897, 17:44163547_ACG_A, rs776509440, rs2668692, rs2696531.

**Table S1. Derivation of final GWAS sample**

461,991 after genetic sample QC: include European (largest group from 4-means clustering), and exclude gender mismatch, excess relatives, putative sex chromosome aneuploidy, heterozygosity and missingness outliers)

**Cardiorespiratory fitness (CRF) | 70,784 final GWAS sample**

**61,227** in Category 1, 2, or 3^1^ (initial assessment)

**17,790** in Category 1, 2, or 3 (first repeat assessment)

**61,227** with age at first assessment (initial assessment)

**17,523** with age at first assessment (first repeat assessment)

**60,395** with 'Bicycle' method of assessment (initial assessment)

**17,458** with 'Bicycle' method of assessment (first repeat assessment)

**57,604** after cycle ramp QC (cadence within 35-125 BPM, F > 30 watts, M > 40 watts, >20 'Exercise' phase heart rate observations^2^) (initial assessment)

**16,655** after cycle ramp QC (first repeat assessment)

**70,783** after phenotype derivation, and initial and first repeat assessments merge^3^

**Physical activity (PA) | 89,683 final GWAS sample**

**89,683** after PA QC (data quality good wear time and good calibration, no data problem indicators, exclude >100 m*g* no wear-time bias adjusted PA)

^1^ Category 1, 2, and 3 included about 90% of samples recruited to the cycle ramp test

^2^ Butterworth filter failed with low number of heart rate observations (or "entries")

^3^ Where a participant had data at both assessments, their initial assessment data was used

**Table S2. CRF cycle ramp test risk categories**

| **Category** | **Protocol** | **Initial assessment*** | **First repeat assessment*** |
| --- | --- | --- | --- |
| 1 | cycle (load) rising to 50% level | 51,464 | 14,796 |
| 2 | cycle (load) rising to 35% level | 7,836 | 2,451 |
| 3 | cycle at constant level | 1,927 | 543 |
| 4 | at-rest measurement | 6,989 | 1,754 |
| No category | ECG not to be done | 1,102 | 222 |

* after genetic meta-data QC (include European, exclude gender mismatch, excess relatives, putative sex chromosome aneuploidy, heterozygosity and missingness outliers), before phenotype-specific QC

**Table S3. Top 10 global cause of death in high-income countries, 2016**

| **Rank** | **Cause of death** | **ICD-10 codes** |
| --- | --- | --- |
| 1 | Ischaemic heart disease | I20 – I25 |
| 2 | Stroke | I60 – I69 |
| 3 | Alzheimer’s disease and other dementias | F01 – F03, G30 – G31 |
| 4 | Trachea, bronchus and lung cancers | C33 – C34 |
| 5 | Chronic obstructive pulmonary disease | J40 – J44 |
| 6 | Lower respiratory tract infections (communicable) | J09 – J22, P23, U04 |
| 7 | Colon and rectum cancers | C18 – C21 |
| 8 | Diabetes mellitus | E10 – E14 (minus E10.2 – E10.29, E11.2 – E11.29, E12.2, E13.2 – E13.29, E14.2) |
| 9 | Kidney diseases | N00 – N19, E10.2 – E10.29, E11.2 – E11.29, E12.2, E13.2 – E13.29, E14.2 |
| 10 | Breast cancer | C50 |

Source: Global Health Estimates 2016: Deaths by Cause, Age, Sex, by Country and by Region, 2000 – 2016. Geneva, World Health Organization; 2018.

World Bank List of economies (June 2017). Washington, DC: The World Bank Group; 2017.

<https://www.who.int/news-room/fact-sheets/detail/the-top-10-causes-of-death>

High-income (including United Kingdom and the United States) definition from World bank

<https://datahelpdesk.worldbank.org/knowledgebase/articles/906519-world-bank-country-and-lending-groups>

<https://www.who.int/healthinfo/global_burden_disease/definition_regions/en/>

Mapping to ICD-10 codes in WHO global burden of disease methods (Annex Table A GHE cause categories and ICD‐10codes) <https://www.who.int/healthinfo/global_burden_disease/GlobalCOD_method_2000_2015.pdf>

**Table S4. Distribution of CRF-vo2max, CRF-slope, and PA**

|  | **N** | **MIN** | **MAX** | **MEAN** | **SE** |
| --- | --- | --- | --- | --- | --- |
| CRF-vo2max | 70783 | 0.06 | 31.0 | 2.70 | 0.004 |
| male | 34419 | 0.27 | 22.0 | 3.12 | 0.006 |
| female | 36364 | 0.06 | 31.0 | 2.30 | 0.005 |
| CRF-slope | 70783 | 0.00 | 3.53 | 0.47 | 0.001 |
| male | 34419 | 0.00 | 3.37 | 0.39 | 0.001 |
| female | 36364 | 0.00 | 3.53 | 0.53 | 0.001 |
| PA | 89683 | 2.57 | 97.10 | 28.10 | 0.028 |
| male | 39352 | 3.55 | 97.10 | 27.60 | 0.044 |
| female | 50331 | 2.57 | 95.20 | 28.50 | 0.036 |

**Table S5. Mean trait value by smoking and alcohol status**

|  | **smoking status** | | |  | **alcohol status** | |  |
| --- | --- | --- | --- | --- | --- | --- | --- |
|  | *CRF-vo2max* | *CRF-slope* | *PA* |  | *CRF-vo2max* | *CRF-slope* | *PA* |
| Never | 2.69 (0.01) 55% | 0.48 (0.00) 55% | 28.5 (0.04) 57% |  | 2.31 (0.02) 3% | 0.51 (0.01) 3% | 27.4 (0.18) 3% |
| Previous | 2.67 (0.01) 36% | 0.45 (0.00) 36% | 27.8 (0.05) 36% |  | 2.58 (0.03) 3% | 0.47 (0.01) 3% | 26.7 (0.18) 3% |
| Current | 2.87 (0.02) 8% | 0.44 (0.00) 8% | 26.7 (0.11) 7% |  | 2.71 (0.00) 94% | 0.46 (0.00) 94% | 28.2 (0.03) 95% |
| Prefer not to answer | 2.56 (0.06) | 0.46 (0.01) | 27.20 (0.53) |  | 2.62 (0.10) | 0.46 (0.02) | 28.10 (1.12) |

Note. Each cell shows mean (standard error) and percent of sample with named status. <1% of the sample indicated 'Prefer not to answer' to smoking and alcohol status for all three traits.

**Table S6. Linear models**

| **trait** | **model** | **R^2^** | **df** | **logL** | **AIC** | **BIC** | **nobs** |
| --- | --- | --- | --- | --- | --- | --- | --- |
| vo2max | m1 | 0.3745 | 21 | -91499.52 | 183045.04 | 183255.89 | 70783 |
|  | m2 | 0.3755 | 22 | -91441.90 | 182931.80 | 183151.82 | 70783 |
|  | m3 | 0.4184 | 23 | -88893.90 | 177837.79 | 178066.97 | 70772 |
|  | m4 | 0.4187 | 27 | -88488.88 | 177035.75 | 177301.47 | 70455 |
| slope | m1 | 0.3013 | 21 | 16915.03 | -33784.06 | -33573.21 | 70783 |
|  | m2 | 0.3051 | 22 | 17106.79 | -34165.57 | -33945.56 | 70783 |
|  | m3 | 0.3131 | 23 | 17511.97 | -34973.94 | -34744.76 | 70772 |
|  | m4 | 0.3143 | 27 | 17491.47 | -34924.94 | -34659.22 | 70455 |
| pa | m1 | 0.0790 | 34 | -313072.71 | 626217.42 | 626555.96 | 89683 |
|  | m2 | 0.0798 | 35 | -313033.58 | 626141.17 | 626489.12 | 89683 |
|  | m3 | 0.1452 | 36 | -309056.44 | 618188.89 | 618546.17 | 89512 |
|  | m4 | 0.1498 | 40 | -307970.79 | 616025.58 | 616420.35 | 89264 |

Note. p-value near 0 for all models, but modest contribution of smoking and alcohol status: CRF-vo2max ΔR^2^_m4-m3_ < 0.001, CRF-slope ΔR^2^ _m4-m3_ = 0.001, PA ΔR^2^ _m4-m3_ = 0.005

m1:	age + sex (for sex combined analyses) + array + centre + trend (CRF only) + category (CRF only)

*m2:	m1 + age^2^

m3:	m2 + BMI

m4:	m3 + smoking status + alcohol status

* model m2 used for all GWAS and post-GWAS analyses described in the manuscript

**Table S7. PA association statistics for rs564819152, chromosome 10**

| **STUDY** | **CHR** | **BP** | **SNP** | **A1** | **A0** | **A1 FREQ** | **BETA** | **SE** | **P** |
| --- | --- | --- | --- | --- | --- | --- | --- | --- | --- |
| Current | 10 | 21820650 | rs564819152 | A | G | 0.68 | 0.230 | 0.040 | 1.1x10^-8^ |
| Previous | 10 | 21820650 | *rs564819152 | A | G | 0.68 | 0.028 | 0.005 | 4.2x10^-9^ |

Note. Differences in magnitude of BETAs between the studies may differences in scale of the QCd PA phenotype, as well as differences in QC protocol (**Table S1**). Current = present study; Previous = Doherty, A., Smith-Byrne, K., Ferreira, T. et al. GWAS identifies 14 loci for device-measured physical activity and sleep duration. Nat Commun 9, 5257 (2018). https://doi.org/10.1038/s41467-018-07743-4. *The GWAS significant chr10 SNP we reported rs34719019 was in LD with rs564819152, *r*^2^ = 0.72 in the sample with PA data (and *r*^2^ = 0.78, Chi-sq = 781.39, p < 0.0001 in 1000 Genomes EUR)

**Table S8. Gene-based association test significant genes**

| **SEX** | **GENE** | **CHR** | **START** | **STOP** | **NSNPS** | **P** |
| --- | --- | --- | --- | --- | --- | --- |
| ***CRF-vo2max*** |  |  |  |  |  |  |
| combined | *SCN10A* | 3 | 38738293 | 38835501 | 319 | 4.03E-08 |
| combined | *ARHGAP27* | 17 | 43471275 | 43511787 | 118 | 2.11E-06 |
| combined | *CRHR1* | 17 | 43699267 | 43913194 | 1157 | 1.54E-07 |
| combined | *SPPL2C* | 17 | 43922256 | 43924438 | 17 | 2.67E-07 |
| combined | *MAPT* | 17 | 43971748 | 44105700 | 841 | 2.29E-07 |
| combined | *STH* | 17 | 44076616 | 44077060 | 1 | 4.20E-07 |
| combined | *KANSL1* | 17 | 44107282 | 44302733 | 1206 | 2.60E-07 |
| combined | *ARL17B* | 17 | 44352150 | 44439130 | 202 | 9.18E-08 |
| combined | *LRRC37A* | 17 | 44370099 | 44415160 | 15 | 1.11E-06 |
| combined | *NSF* | 17 | 44668035 | 44834830 | 84 | 8.38E-08 |
| combined | *WNT3* | 17 | 44839872 | 44910520 | 134 | 5.97E-09 |
| combined | *KIAA1755* | 20 | 36838890 | 36889174 | 152 | 1.25E-09 |
| male | *SCN10A* | 3 | 38738293 | 38835501 | 319 | 7.44E-07 |
| male | *KIAA1755* | 20 | 36838890 | 36889174 | 152 | 2.83E-07 |
| ***CRF-slope*** |  |  |  |  |  |  |
| combined | *TRIP6* | 7 | 100464760 | 100471076 | 16 | 7.61E-08 |
| combined | *MUC3A* | 7 | 100547257 | 100550424 | 16 | 1.65E-06 |
| combined | *MFHAS1* | 8 | 8640864 | 8751155 | 552 | 8.07E-07 |
| combined | *ERI1* | 8 | 8859657 | 8974256 | 526 | 1.82E-06 |
| combined | *MSRA* | 8 | 9911778 | 10286401 | 1635 | 1.66E-06 |
| combined | *RP1L1* | 8 | 10463859 | 10569697 | 314 | 7.51E-07 |
| combined | *C8orf74* | 8 | 10530147 | 10558103 | 41 | 3.39E-07 |
| combined | *SOX7* | 8 | 10581278 | 10697357 | 653 | 6.58E-07 |
| combined | *SOX7* | 8 | 10582909 | 10697357 | 648 | 1.26E-06 |
| combined | *PINX1* | 8 | 10622473 | 10697394 | 471 | 1.39E-06 |
| combined | *XKR6* | 8 | 10753555 | 11058875 | 1179 | 2.54E-09 |
| combined | *AF131215.5* | 8 | 10983980 | 10987745 | 13 | 4.12E-07 |
| combined | *BLK* | 8 | 11351510 | 11422113 | 315 | 3.81E-07 |
| combined | *RNF183* | 9 | 116059373 | 116065656 | 20 | 1.54E-06 |
| combined | *SYT10* | 12 | 33527173 | 33592754 | 230 | 9.95E-09 |
| ***PA*** |  |  |  |  |  |  |
| combined | *EBF1* | 5 | 158122928 | 158526769 | 863 | 9.37E-07 |
| combined | *MAPKAP1* | 9 | 128199672 | 128469513 | 517 | 1.85E-07 |
| combined | *SKIDA1* | 10 | 21802407 | 21814611 | 15 | 1.96E-07 |
| combined | *MLLT10* | 10 | 21823094 | 22032559 | 237 | 1.00E-09 |
| combined | *ARHGAP27* | 17 | 43471275 | 43511787 | 118 | 1.22E-08 |
| combined | *PLEKHM1* | 17 | 43513266 | 43568115 | 162 | 2.25E-08 |
| combined | *CRHR1* | 17 | 43699267 | 43913194 | 1157 | 5.65E-11 |
| combined | *SPPL2C* | 17 | 43922256 | 43924438 | 17 | 2.17E-10 |
| combined | *MAPT* | 17 | 43971748 | 44105700 | 841 | 3.60E-11 |
| combined | *STH* | 17 | 44076616 | 44077060 | 1 | 8.10E-11 |
| combined | *KANSL1* | 17 | 44107282 | 44302733 | 1206 | 5.30E-11 |
| combined | *ARL17B* | 17 | 44352150 | 44439130 | 202 | 2.12E-10 |
| combined | *LRRC37A* | 17 | 44370099 | 44415160 | 15 | 1.07E-08 |
| combined | *LRRC37A2* | 17 | 44588877 | 44633016 | 11 | 8.68E-09 |
| combined | *ARL17A* | 17 | 44594068 | 44657088 | 16 | 1.42E-07 |
| combined | *NSF* | 17 | 44668035 | 44834830 | 84 | 8.98E-09 |
| combined | *WNT3* | 17 | 44839872 | 44910520 | 134 | 1.23E-06 |
| combined | *L3MBTL2* | 22 | 41601209 | 41627275 | 98 | 1.96E-06 |
| male | *CNTNAP5** | 2 | 124782864 | 125672864 | 3136 | 5.46E-07 |
| female | *EBF1* | 5 | 158122928 | 158526769 | 863 | 4.06E-08 |
| female | *DOCK1** | 10 | 128593978 | 129250781 | 2064 | 2.58E-07 |
| female | *CLEC18A** | 16 | 69984810 | 69998141 | 28 | 5.45E-07 |
| female | *TAT** | 16 | 71599563 | 71611033 | 30 | 1.12E-06 |
| female | *PHLPP2** | 16 | 71671738 | 71758604 | 198 | 1.18E-06 |
| female | *ARHGAP27* | 17 | 43471275 | 43511787 | 118 | 2.09E-06 |
| female | *CRHR1* | 17 | 43699267 | 43913194 | 1157 | 2.49E-09 |
| female | *SPPL2C* | 17 | 43922256 | 43924438 | 17 | 4.70E-09 |
| female | *MAPT* | 17 | 43971748 | 44105700 | 841 | 1.84E-09 |
| female | *STH* | 17 | 44076616 | 44077060 | 1 | 4.50E-09 |
| female | *KANSL1* | 17 | 44107282 | 44302733 | 1206 | 2.59E-09 |
| female | *ARL17B* | 17 | 44352150 | 44439130 | 202 | 4.01E-09 |
| female | *LRRC37A* | 17 | 44370099 | 44415160 | 15 | 1.17E-07 |
| female | *LRRC37A2* | 17 | 44588877 | 44633016 | 11 | 3.50E-09 |
| female | *ARL17A* | 17 | 44594068 | 44657088 | 16 | 5.11E-08 |
| female | *NSF* | 17 | 44668035 | 44834830 | 84 | 4.92E-08 |
| female | *WNT3* | 17 | 44839872 | 44910520 | 134 | 1.34E-06 |
| female | *CCNE1** | 19 | 30302805 | 30315215 | 36 | 1.08E-07 |

Note. Input SNPs were mapped to 18988 protein coding genes. Genome wide significance P = 0.05/18988 = 2.633e-6. * not associated in the combined sample analysis

**Table S9. Genetic correlation (FDR-significant)**

|  |  |  |  | *combined* | | | *male* | | | *female* | | |
| --- | --- | --- | --- | --- | --- | --- | --- | --- | --- | --- | --- | --- |
| **Category** | **PMID: GWAS** | ***h*^2^** | ***h*^2^ se** | ***r*_g_** | **se** | **p_adj** | ***r*_g_** | **se** | **p_adj** | ***r*_g_** | **se** | **p_adj** |
| CRF-vo2max |  |  |  |  |  |  |  |  |  |  |  |  |
| aging | 27015805: Fathers age at death | 0.04 | 0.01 | 0.37 | 0.09 | 6.9E-04 | NA | NA | NA | 0.49 | 0.12 | 9.8E-04 |
| aging | 27015805: Parents age at death | 0.03 | 0.01 | NA | NA | NA | NA | NA | NA | 0.42 | 0.15 | 4.2E-02 |
| anthropometric | 20935630: Body mass index | 0.19 | 0.01 | -0.29 | 0.04 | 8.9E-09 | -0.25 | 0.06 | 1.2E-04 | -0.33 | 0.06 | 1.3E-05 |
| anthropometric | 26833246: Body fat | 0.11 | 0.01 | -0.44 | 0.06 | 2.5E-10 | -0.44 | 0.08 | 1.1E-06 | -0.44 | 0.08 | 2.6E-06 |
| anthropometric | 23563607: Extreme bmi | 0.70 | 0.05 | -0.30 | 0.06 | 1.2E-05 | -0.29 | 0.07 | 1.8E-03 | -0.32 | 0.09 | 4.7E-03 |
| anthropometric | 20881960: Height_2010 | 0.29 | 0.02 | 0.13 | 0.04 | 1.8E-02 | NA | NA | NA | 0.14 | 0.05 | 3.9E-02 |
| anthropometric | 23563607: Obesity class 1 | 0.22 | 0.01 | -0.34 | 0.05 | 6.9E-11 | -0.32 | 0.06 | 6.2E-06 | -0.36 | 0.06 | 1.7E-06 |
| anthropometric | 23563607: Obesity class 2 | 0.19 | 0.01 | -0.36 | 0.05 | 1.4E-09 | -0.32 | 0.07 | 9.1E-05 | -0.39 | 0.07 | 1.9E-06 |
| anthropometric | 23563607: Obesity class 3 | 0.12 | 0.01 | -0.38 | 0.07 | 3.8E-06 | -0.33 | 0.09 | 6.0E-03 | -0.41 | 0.10 | 8.8E-04 |
| anthropometric | 23563607: Overweight | 0.11 | 0.01 | -0.33 | 0.05 | 7.6E-10 | -0.30 | 0.06 | 6.0E-06 | -0.35 | 0.07 | 2.7E-05 |
| anthropometric | 25673412: Hip circumference | 0.13 | 0.01 | -0.26 | 0.04 | 1.3E-07 | -0.27 | 0.06 | 2.7E-04 | -0.26 | 0.06 | 1.1E-04 |
| anthropometric | 25673412: Waist circumference | 0.12 | 0.01 | -0.29 | 0.05 | 1.9E-08 | -0.26 | 0.06 | 8.7E-04 | -0.33 | 0.06 | 2.7E-06 |
| anthropometric | 25673412: Waist-to-hip ratio | 0.12 | 0.01 | -0.23 | 0.04 | 2.0E-06 | NA | NA | NA | -0.34 | 0.06 | 1.1E-06 |
| anthropometric | 27680694: Birth weight | 0.10 | 0.01 | 0.14 | 0.05 | 2.4E-02 | NA | NA | NA | 0.20 | 0.06 | 1.2E-02 |
| anthropometric | 31043758: Own birth weight | 0.10 | 0.01 | 0.12 | 0.04 | 2.6E-02 | NA | NA | NA | 0.17 | 0.05 | 7.0E-03 |
| anthropometric | 31043758: Own birth weight | 0.09 | 0.01 | 0.13 | 0.04 | 1.7E-02 | NA | NA | NA | 0.17 | 0.05 | 8.1E-03 |
| anthropometric | 23449627: Difference in height between childhood and adulthood; age 8 | 0.33 | 0.05 | NA | NA | NA | NA | NA | NA | 0.34 | 0.12 | 3.3E-02 |
| cardiometabolic | 26343387: Coronary artery disease | 0.08 | 0.01 | -0.15 | 0.05 | 1.5E-02 | NA | NA | NA | -0.24 | 0.06 | 6.4E-04 |
| cognitive | 28530673: Intelligence | 0.20 | 0.01 | 0.16 | 0.05 | 9.2E-03 | NA | NA | NA | 0.21 | 0.06 | 6.0E-03 |
| education | 23358156: Childhood IQ | 0.29 | 0.05 | 0.28 | 0.08 | 1.0E-02 | NA | NA | NA | 0.37 | 0.12 | 1.5E-02 |
| education | 27225129: Years of schooling 2016 | 0.13 | 0.00 | 0.28 | 0.04 | 3.6E-11 | 0.25 | 0.05 | 2.6E-05 | 0.31 | 0.05 | 4.8E-09 |
| education | 23722424: College completion | 0.08 | 0.01 | 0.32 | 0.05 | 8.2E-08 | 0.33 | 0.08 | 3.8E-04 | 0.33 | 0.07 | 1.7E-05 |
| education | 25201988: Years of schooling (proxy cognitive performance) | 0.11 | 0.01 | 0.28 | 0.05 | 6.0E-06 | 0.29 | 0.08 | 1.8E-03 | 0.30 | 0.07 | 1.7E-04 |
| education | 23722424: Years of schooling 2013 | 0.09 | 0.01 | 0.31 | 0.05 | 6.8E-07 | 0.31 | 0.07 | 5.4E-04 | 0.32 | 0.07 | 8.5E-05 |
| glycemic | 22581228: Fasting insulin main effect | 0.07 | 0.01 | -0.45 | 0.09 | 2.5E-05 | -0.53 | 0.12 | 1.7E-04 | -0.34 | 0.11 | 1.8E-02 |
| glycemic | 20858683: HbA1C | 0.06 | 0.01 | -0.25 | 0.08 | 3.1E-02 | -0.33 | 0.11 | 1.9E-02 | NA | NA | NA |
| glycemic | 20081858: HOMA-B | 0.09 | 0.01 | -0.40 | 0.09 | 4.1E-04 | -0.43 | 0.12 | 6.0E-03 | -0.35 | 0.12 | 3.5E-02 |
| glycemic | 20081858: HOMA-IR | 0.07 | 0.01 | -0.58 | 0.12 | 4.9E-05 | -0.67 | 0.16 | 5.7E-04 | -0.46 | 0.14 | 1.2E-02 |
| haemotological | 22139419: Platelet count | 0.12 | 0.01 | NA | NA | NA | -0.22 | 0.08 | 3.5E-02 | NA | NA | NA |
| hormone | 26833098: Leptin_adjBMI | 0.10 | 0.02 | -0.32 | 0.08 | 1.7E-03 | -0.38 | 0.12 | 1.4E-02 | NA | NA | NA |
| hormone | 26833098: Leptin_not_adjBMI | 0.10 | 0.02 | -0.41 | 0.08 | 2.8E-05 | -0.46 | 0.12 | 3.4E-03 | -0.35 | 0.11 | 1.8E-02 |
| lipids | 20686565: Triglycerides | 0.17 | 0.03 | -0.17 | 0.05 | 1.8E-03 | NA | NA | NA | -0.23 | 0.06 | 4.7E-03 |
| lung_function | 28166213: Forced expiratory volume in 1 second (FEV1) | 0.27 | 0.02 | 0.18 | 0.05 | 8.1E-03 | 0.26 | 0.06 | 1.0E-03 | NA | NA | NA |
| lung_function | 28166213: Forced Vital capacity(FVC) | 0.26 | 0.02 | 0.22 | 0.05 | 6.3E-04 | 0.26 | 0.06 | 1.3E-03 | 0.19 | 0.07 | 4.8E-02 |
| lung_function | 26635082: Forced expiratory volume in 1 second (FEV1) | 0.14 | 0.02 | 0.19 | 0.07 | 3.7E-02 | NA | NA | NA | 0.31 | 0.09 | 4.7E-03 |
| lung_function | 26635082: Forced Vital capacity(FVC) | 0.15 | 0.02 | 0.25 | 0.06 | 1.5E-03 | NA | NA | NA | 0.36 | 0.08 | 2.7E-04 |
| lung_function | 0: Forced expiratory volume in 1 second | 0.17 | 0.01 | 0.19 | 0.03 | 2.2E-06 | 0.18 | 0.04 | 6.6E-04 | 0.21 | 0.05 | 1.9E-04 |
| lung_function | 0: Forced vital capacity | 0.16 | 0.01 | 0.23 | 0.03 | 2.5E-09 | 0.20 | 0.04 | 1.1E-04 | 0.26 | 0.05 | 5.4E-07 |
| lung_function | 0: Peak expiratory flow | 0.14 | 0.01 | 0.12 | 0.03 | 4.7E-03 | 0.14 | 0.04 | 2.3E-02 | NA | NA | NA |
| lung_function | 0: Forced expiratory volume in 1 second | 0.18 | 0.01 | 0.18 | 0.04 | 2.3E-05 | 0.19 | 0.04 | 7.8E-04 | 0.19 | 0.05 | 1.8E-03 |
| lung_function | 0: Forced vital capacity | 0.18 | 0.01 | 0.22 | 0.04 | 5.3E-08 | 0.20 | 0.04 | 1.4E-04 | 0.25 | 0.05 | 6.4E-06 |
| lung_function | 0: Peak expiratory flow | 0.15 | 0.01 | 0.12 | 0.03 | 9.2E-03 | NA | NA | NA | NA | NA | NA |
| lung_function | 0: Forced expiratory volume in 1 second | 0.13 | 0.01 | 0.24 | 0.05 | 1.1E-04 | NA | NA | NA | 0.35 | 0.08 | 1.4E-04 |
| lung_function | 0: Forced vital capacity | 0.12 | 0.01 | 0.26 | 0.05 | 1.5E-05 | NA | NA | NA | 0.36 | 0.07 | 1.7E-05 |
| metabolites | 27005778: Glycoprotein acetyls; mainly a1-acid glycoprotein | 0.11 | 0.03 | -0.29 | 0.10 | 3.9E-02 | NA | NA | NA | NA | NA | NA |
| metabolites | 27005778: Acetoacetate | 0.07 | 0.03 | NA | NA | NA | NA | NA | NA | 0.42 | 0.15 | 3.9E-02 |
| psychiatric | 27089181: Depressive symptoms | 0.05 | 0.00 | -0.21 | 0.06 | 4.7E-03 | -0.22 | 0.07 | 3.3E-02 | -0.23 | 0.08 | 2.5E-02 |
| psychiatric | 24514567: Anorexia Nervosa | 0.55 | 0.03 | 0.14 | 0.05 | 3.9E-02 | NA | NA | NA | NA | NA | NA |
| psychiatric | 21926972: Bipolar disorder | 0.45 | 0.04 | 0.18 | 0.06 | 3.3E-02 | NA | NA | NA | NA | NA | NA |
| reproductive | 25231870: Age at Menarche | 0.21 | 0.01 | 0.11 | 0.04 | 3.1E-02 | NA | NA | NA | 0.16 | 0.05 | 9.2E-03 |
| reproductive | 27798627: Age of first birth | 0.07 | 0.00 | 0.22 | 0.05 | 3.8E-05 | 0.22 | 0.07 | 1.2E-02 | 0.22 | 0.05 | 8.4E-04 |
| sleeping | 28604731: Insomnia | 0.05 | 0.01 | -0.17 | 0.06 | 4.8E-02 | NA | NA | NA | NA | NA | NA |
| sleeping | 27992416: Insomnia | 0.14 | 0.01 | -0.21 | 0.06 | 6.0E-03 | NA | NA | NA | -0.28 | 0.08 | 7.0E-03 |
| smoking_behaviour | 20418890: Ever vs never smoked | 0.07 | 0.01 | NA | NA | NA | NA | NA | NA | -0.23 | 0.08 | 4.8E-02 |
| CRF-slope |  |  |  |  |  |  |  |  |  |  |  |  |
| anthropometric | 20935630: Body mass index | 0.19 | 0.01 | -0.18 | 0.05 | 1.8E-03 | -0.34 | 0.06 | 1.9E-06 | NA | NA | NA |
| anthropometric | 22484627: Childhood obesity | 0.41 | 0.05 | -0.24 | 0.06 | 3.4E-03 | -0.34 | 0.08 | 9.8E-04 | NA | NA | NA |
| anthropometric | 23449627: Height; Females at age 10 and males at age 12 | 0.44 | 0.05 | -0.26 | 0.07 | 3.4E-03 | NA | NA | NA | -0.34 | 0.09 | 3.4E-03 |
| anthropometric | 27680694: Birth weight | 0.10 | 0.01 | -0.15 | 0.05 | 3.5E-02 | NA | NA | NA | NA | NA | NA |
| anthropometric | 31043758: Own birth weight | 0.09 | 0.01 | -0.12 | 0.04 | 4.3E-02 | -0.15 | 0.06 | 4.7E-02 | NA | NA | NA |
| anthropometric | 23563607: Obesity class 1 | 0.22 | 0.01 | NA | NA | NA | -0.25 | 0.07 | 1.8E-03 | NA | NA | NA |
| anthropometric | 23563607: Obesity class 2 | 0.19 | 0.01 | NA | NA | NA | -0.28 | 0.08 | 6.0E-03 | NA | NA | NA |
| anthropometric | 23563607: Overweight | 0.11 | 0.01 | NA | NA | NA | -0.22 | 0.07 | 1.0E-02 | NA | NA | NA |
| anthropometric | 25673412: Hip circumference | 0.13 | 0.01 | NA | NA | NA | -0.19 | 0.06 | 1.7E-02 | NA | NA | NA |
| anthropometric | 25673412: Waist circumference | 0.12 | 0.01 | NA | NA | NA | -0.24 | 0.06 | 9.8E-04 | NA | NA | NA |
| anthropometric | 25673412: Waist-to-hip ratio | 0.12 | 0.01 | NA | NA | NA | -0.19 | 0.06 | 1.8E-02 | NA | NA | NA |
| bone | 26367794: Lumbar Spine bone mineral density | 0.14 | 0.02 | -0.21 | 0.08 | 4.2E-02 | NA | NA | NA | NA | NA | NA |
| bone | 22504420: Femoral neck bone mineral density | 0.31 | 0.03 | NA | NA | NA | -0.20 | 0.07 | 2.8E-02 | NA | NA | NA |
| brain_volume | 25607358: Mean Hippocampus | 0.16 | 0.04 | -0.42 | 0.12 | 9.2E-03 | -0.48 | 0.15 | 2.0E-02 | NA | NA | NA |
| education | 23722424: College completion | 0.08 | 0.01 | -0.17 | 0.05 | 7.0E-03 | NA | NA | NA | -0.22 | 0.07 | 1.0E-02 |
| education | 25201988: Years of schooling (proxy cognitive performance) | 0.11 | 0.01 | -0.15 | 0.05 | 3.1E-02 | NA | NA | NA | -0.22 | 0.06 | 6.0E-03 |
| education | 23722424: Years of schooling 2013 | 0.09 | 0.01 | -0.16 | 0.05 | 3.9E-02 | NA | NA | NA | -0.22 | 0.07 | 1.0E-02 |
| education | 23358156: Childhood IQ | 0.29 | 0.05 | NA | NA | NA | NA | NA | NA | -0.38 | 0.13 | 2.6E-02 |
| education | 27225129: Years of schooling 2016 | 0.13 | 0.00 | NA | NA | NA | NA | NA | NA | -0.16 | 0.05 | 2.4E-02 |
| glycemic | 22885922: Type 2 Diabetes | 0.09 | 0.01 | NA | NA | NA | -0.28 | 0.09 | 2.3E-02 | NA | NA | NA |
| haemotological | 23583979: Heart rate | 0.08 | 0.01 | -0.31 | 0.07 | 5.7E-04 | -0.36 | 0.08 | 4.9E-04 | NA | NA | NA |
| lung_function | 0: Forced vital capacity | 0.16 | 0.01 | -0.13 | 0.04 | 6.0E-03 | NA | NA | NA | -0.15 | 0.05 | 1.5E-02 |
| lung_function | 0: Peak expiratory flow | 0.14 | 0.01 | -0.14 | 0.04 | 1.8E-02 | NA | NA | NA | NA | NA | NA |
| lung_function | 0: Forced vital capacity | 0.18 | 0.01 | -0.13 | 0.04 | 1.5E-02 | NA | NA | NA | -0.14 | 0.05 | 3.5E-02 |
| lung_function | 0: Peak expiratory flow | 0.15 | 0.01 | -0.14 | 0.04 | 1.7E-02 | NA | NA | NA | NA | NA | NA |
| sleeping | 27494321: Sleep duration | 0.06 | 0.01 | NA | NA | NA | NA | NA | NA | -0.25 | 0.09 | 4.8E-02 |
| PA |  |  |  |  |  |  |  |  |  |  |  |  |
| aging | 27015805: Parents age at death | 0.03 | 0.01 | 0.28 | 0.08 | 7.0E-03 | NA | NA | NA | 0.48 | 0.10 | 8.1E-05 |
| aging | 27015805: Fathers age at death | 0.04 | 0.01 | 0.30 | 0.07 | 1.2E-03 | NA | NA | NA | 0.33 | 0.09 | 4.7E-03 |
| aging | 27015805: Mothers age at death | 0.04 | 0.01 | NA | NA | NA | NA | NA | NA | 0.28 | 0.09 | 3.1E-02 |
| anthropometric | 20935630: Body mass index | 0.19 | 0.01 | -0.20 | 0.03 | 2.3E-07 | -0.16 | 0.05 | 8.1E-03 | -0.23 | 0.04 | 5.4E-07 |
| anthropometric | 26833246: Body fat | 0.11 | 0.01 | -0.37 | 0.04 | 3.3E-15 | -0.25 | 0.06 | 5.9E-04 | -0.46 | 0.05 | 2.0E-14 |
| anthropometric | 23563607: Extreme bmi | 0.70 | 0.05 | -0.17 | 0.05 | 7.0E-03 | NA | NA | NA | -0.21 | 0.06 | 1.2E-02 |
| anthropometric | 23563607: Extreme height | 1.23 | 0.11 | -0.15 | 0.05 | 1.4E-02 | -0.19 | 0.06 | 2.1E-02 | NA | NA | NA |
| anthropometric | 20881960: Height_2010 | 0.29 | 0.02 | -0.12 | 0.03 | 2.4E-04 | -0.12 | 0.04 | 8.1E-03 | -0.12 | 0.04 | 1.6E-02 |
| anthropometric | 23563607: Obesity class 1 | 0.22 | 0.01 | -0.23 | 0.04 | 2.5E-08 | -0.14 | 0.05 | 2.3E-02 | -0.29 | 0.04 | 8.9E-09 |
| anthropometric | 23563607: Obesity class 2 | 0.19 | 0.01 | -0.23 | 0.04 | 3.0E-06 | NA | NA | NA | -0.30 | 0.05 | 2.8E-07 |
| anthropometric | 23563607: Obesity class 3 | 0.12 | 0.01 | -0.22 | 0.06 | 3.4E-03 | NA | NA | NA | -0.26 | 0.07 | 4.7E-03 |
| anthropometric | 23563607: Overweight | 0.11 | 0.01 | -0.23 | 0.04 | 7.9E-08 | -0.16 | 0.05 | 2.1E-02 | -0.28 | 0.04 | 3.1E-08 |
| anthropometric | 25673412: Hip circumference | 0.13 | 0.01 | -0.31 | 0.03 | 1.1E-18 | -0.24 | 0.05 | 6.4E-06 | -0.37 | 0.04 | 7.9E-16 |
| anthropometric | 25673412: Waist circumference | 0.12 | 0.01 | -0.31 | 0.03 | 1.2E-18 | -0.24 | 0.04 | 2.0E-06 | -0.35 | 0.04 | 1.4E-14 |
| anthropometric | 25673412: Waist-to-hip ratio | 0.12 | 0.01 | -0.18 | 0.03 | 3.0E-06 | -0.14 | 0.04 | 1.6E-02 | -0.20 | 0.04 | 4.5E-05 |
| autoimmune | 26192919: Crohns disease | 0.51 | 0.06 | -0.11 | 0.04 | 3.5E-02 | NA | NA | NA | NA | NA | NA |
| autoimmune | 26192919: Inflammatory Bowel Disease (Euro) | 0.34 | 0.04 | NA | NA | NA | NA | NA | NA | -0.13 | 0.05 | 3.9E-02 |
| cardiometabolic | 26343387: Coronary artery disease | 0.08 | 0.01 | -0.21 | 0.04 | 3.3E-07 | -0.20 | 0.05 | 2.6E-04 | -0.20 | 0.05 | 3.8E-04 |
| education | 27225129: Years of schooling 2016 | 0.13 | 0.00 | 0.11 | 0.03 | 3.4E-03 | NA | NA | NA | 0.14 | 0.03 | 8.7E-04 |
| education | 25201988: Years of schooling (proxy cognitive performance) | 0.11 | 0.01 | 0.14 | 0.04 | 1.6E-02 | NA | NA | NA | NA | NA | NA |
| glycemic | 22885922: Type 2 Diabetes | 0.09 | 0.01 | -0.23 | 0.05 | 8.5E-05 | -0.20 | 0.07 | 3.5E-02 | -0.26 | 0.06 | 9.6E-04 |
| glycemic | 22581228: Fasting insulin main effect | 0.07 | 0.01 | -0.31 | 0.07 | 1.1E-04 | -0.25 | 0.08 | 3.0E-02 | -0.35 | 0.08 | 7.0E-04 |
| glycemic | 20081858: HOMA-IR | 0.07 | 0.01 | -0.29 | 0.07 | 9.8E-04 | NA | NA | NA | -0.29 | 0.10 | 3.7E-02 |
| hormone | 26833098: Leptin_adjBMI | 0.10 | 0.02 | -0.21 | 0.07 | 2.0E-02 | NA | NA | NA | NA | NA | NA |
| hormone | 26833098: Leptin_not_adjBMI | 0.10 | 0.02 | -0.37 | 0.07 | 2.0E-06 | -0.36 | 0.09 | 7.0E-04 | -0.35 | 0.08 | 4.8E-04 |
| lipids | 20686565: HDL cholesterol | 0.13 | 0.03 | 0.28 | 0.04 | 4.6E-10 | 0.30 | 0.06 | 9.4E-06 | 0.27 | 0.05 | 2.0E-06 |
| lipids | 20686565: Triglycerides | 0.17 | 0.03 | -0.20 | 0.04 | 2.2E-06 | -0.16 | 0.05 | 1.5E-02 | -0.22 | 0.05 | 1.5E-04 |
| lung_function | 28166213: Forced Vital capacity(FVC) | 0.26 | 0.02 | 0.13 | 0.04 | 7.0E-03 | NA | NA | NA | 0.14 | 0.05 | 2.2E-02 |
| lung_function | 0: Forced expiratory volume in 1 second | 0.17 | 0.01 | 0.10 | 0.02 | 1.2E-03 | NA | NA | NA | 0.12 | 0.03 | 6.4E-04 |
| lung_function | 0: Forced vital capacity | 0.16 | 0.01 | 0.14 | 0.03 | 1.1E-06 | 0.11 | 0.04 | 2.9E-02 | 0.16 | 0.03 | 9.0E-07 |
| lung_function | 0: Forced expiratory volume in 1 second | 0.18 | 0.01 | 0.10 | 0.03 | 9.8E-04 | NA | NA | NA | 0.12 | 0.03 | 2.8E-04 |
| lung_function | 0: Forced vital capacity | 0.18 | 0.01 | 0.15 | 0.03 | 1.0E-06 | 0.12 | 0.04 | 1.8E-02 | 0.17 | 0.03 | 9.0E-07 |
| lung_function | 0: FEV1/FVC | 0.19 | 0.01 | -0.08 | 0.03 | 2.3E-02 | NA | NA | NA | NA | NA | NA |
| metabolites | 27005778: Acetate | 0.05 | 0.02 | 0.36 | 0.13 | 4.8E-02 | NA | NA | NA | NA | NA | NA |
| metabolites | 27005778: Apolipoprotein A-I | 0.06 | 0.03 | 0.35 | 0.13 | 4.6E-02 | NA | NA | NA | NA | NA | NA |
| metabolites | 27005778: Total cholesterol in HDL | 0.09 | 0.03 | 0.33 | 0.09 | 3.4E-03 | NA | NA | NA | 0.38 | 0.11 | 6.0E-03 |
| metabolites | 27005778: Total cholesterol in large HDL | 0.10 | 0.03 | 0.29 | 0.08 | 4.7E-03 | NA | NA | NA | 0.35 | 0.10 | 7.0E-03 |
| metabolites | 27005778: Cholesterol esters in large HDL | 0.12 | 0.03 | 0.28 | 0.08 | 4.7E-03 | NA | NA | NA | 0.31 | 0.10 | 1.8E-02 |
| metabolites | 27005778: Free cholesterol in large HDL | 0.10 | 0.03 | 0.27 | 0.08 | 8.1E-03 | NA | NA | NA | 0.33 | 0.10 | 7.0E-03 |
| metabolites | 27005778: Total lipids in large HDL | 0.12 | 0.03 | 0.27 | 0.08 | 7.0E-03 | NA | NA | NA | 0.30 | 0.09 | 1.7E-02 |
| metabolites | 27005778: Concentration of large HDL particles | 0.12 | 0.03 | 0.26 | 0.08 | 8.1E-03 | NA | NA | NA | 0.29 | 0.09 | 1.8E-02 |
| metabolites | 27005778: Phospholipids in large HDL | 0.12 | 0.03 | 0.27 | 0.08 | 7.0E-03 | NA | NA | NA | 0.31 | 0.10 | 1.5E-02 |
| metabolites | 27005778: Total lipids in large VLDL | 0.14 | 0.03 | -0.22 | 0.08 | 3.7E-02 | NA | NA | NA | NA | NA | NA |
| metabolites | 27005778: Phospholipids in large VLDL | 0.12 | 0.03 | -0.24 | 0.08 | 3.1E-02 | NA | NA | NA | -0.26 | 0.09 | 4.8E-02 |
| metabolites | 27005778: Total cholesterol in medium HDL | 0.05 | 0.03 | 0.42 | 0.15 | 3.3E-02 | NA | NA | NA | 0.53 | 0.18 | 3.3E-02 |
| metabolites | 27005778: Free cholesterol in medium HDL | 0.07 | 0.02 | 0.38 | 0.12 | 1.6E-02 | NA | NA | NA | 0.46 | 0.14 | 1.8E-02 |
| metabolites | 27005778: Phospholipids in medium HDL | 0.06 | 0.02 | 0.33 | 0.12 | 3.9E-02 | NA | NA | NA | 0.41 | 0.14 | 3.3E-02 |
| metabolites | 27005778: Cholesterol esters in large VLDL | 0.16 | 0.04 | NA | NA | NA | NA | NA | NA | -0.22 | 0.08 | 4.9E-02 |
| metabolites | 27005778: Concentration of large VLDL particles | 0.13 | 0.04 | NA | NA | NA | NA | NA | NA | -0.27 | 0.09 | 2.9E-02 |
| reproductive | 25231870: Age at Menarche | 0.21 | 0.01 | 0.10 | 0.03 | 1.6E-02 | NA | NA | NA | 0.13 | 0.04 | 1.5E-02 |
| reproductive | 27798627: Age of first birth | 0.07 | 0.00 | 0.13 | 0.04 | 3.4E-03 | NA | NA | NA | 0.17 | 0.05 | 6.0E-03 |
| reproductive | 27798627: Number of children ever born | 0.03 | 0.00 | NA | NA | NA | -0.18 | 0.06 | 1.2E-02 | NA | NA | NA |
| sleeping | 27494321: Chronotype | 0.10 | 0.01 | 0.12 | 0.04 | 3.3E-02 | NA | NA | NA | 0.16 | 0.05 | 8.1E-03 |
| sleeping | 27992416: Insomnia | 0.14 | 0.01 | -0.16 | 0.04 | 4.7E-03 | NA | NA | NA | -0.19 | 0.06 | 1.0E-02 |
| smoking_behaviour | 20418890: Former vs Current smoker | 0.06 | 0.01 | 0.27 | 0.07 | 3.4E-03 | NA | NA | NA | 0.32 | 0.09 | 4.7E-03 |
| smoking_behaviour | 20418890: Cigarettes smoked per day | 0.06 | 0.02 | NA | NA | NA | NA | NA | NA | -0.31 | 0.11 | 3.6E-02 |

Note. *h*^2^ = observed scale heritability; p_adj = FDR-adjusted p-value; NA = missing, i.e., *r*_g_ not significant

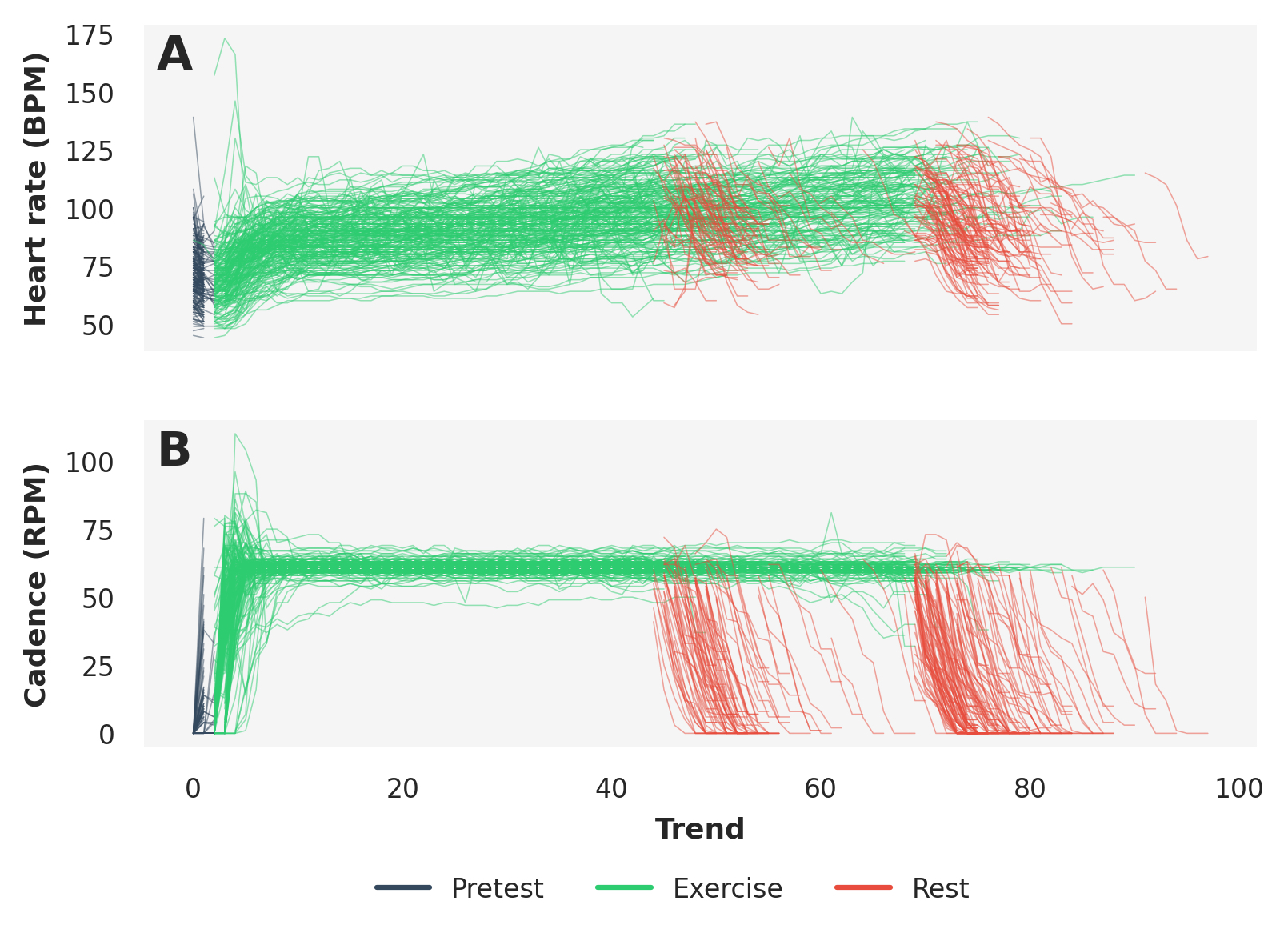

**Figure S1. Raw heart rate and cadence.** Panel **A** shows the heart rate and panel **B** the cadence for 1000 random samples drawn from the initial assessment and first repeat assessment combined. Trend = repeated measurement points during the submaximal cycle ramp test (a proxy for time); Heart Rate (BPM) = 4-lead electrocardiograph (ECG) heart rate data; Cadence (RPM) = stationary bicycle pedal speed – instruction was to cycle at 60 revolutions per minute

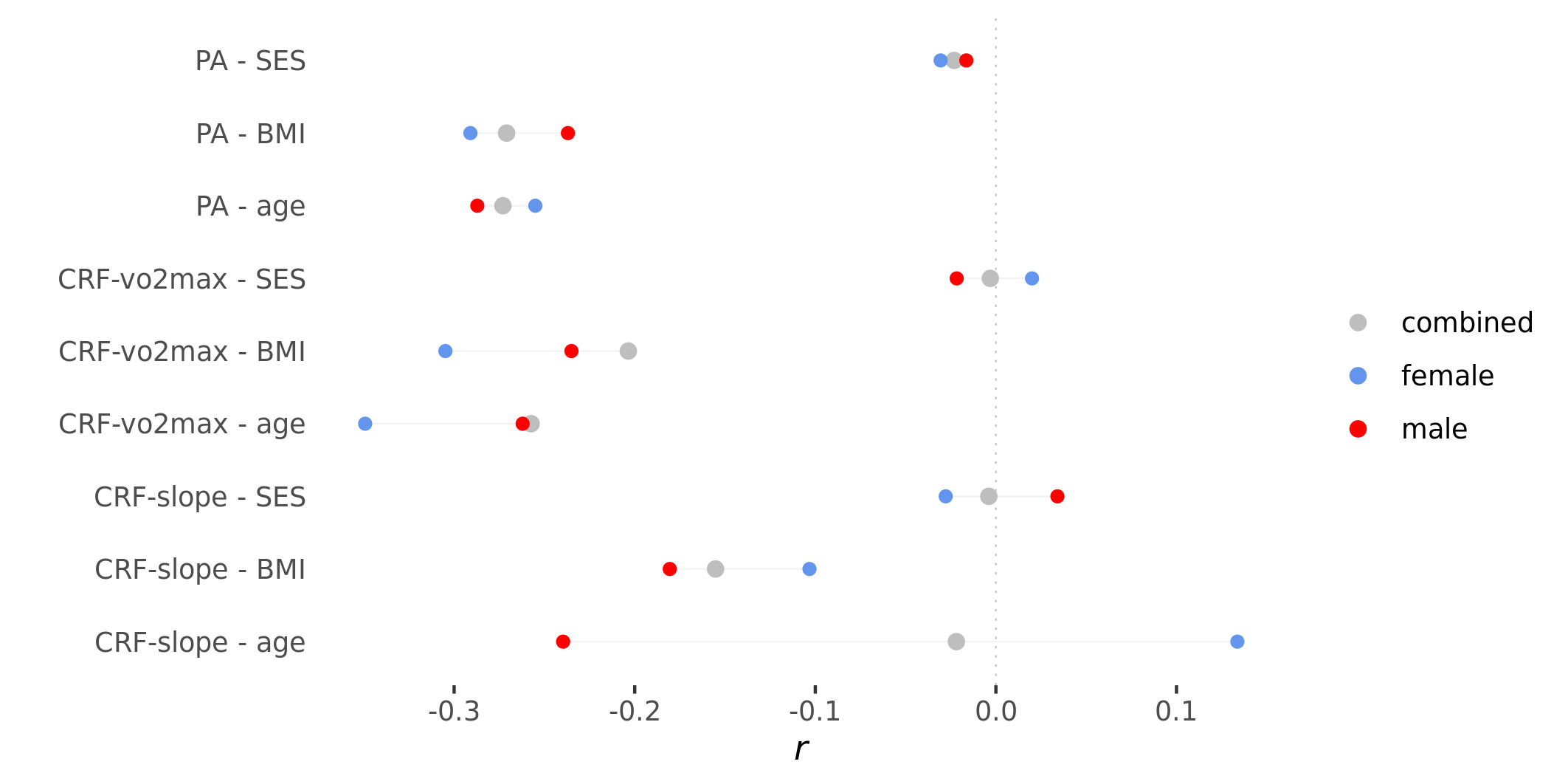

**Figure S2. Phenotypic correlations between CRF and PA, and age, BMI, and SES.** *r* = Pearson’s product moment correlation coefficient; SES = socioeconomic status, measured with the Townsend Deprivation Index (larger positive values indicate greater deprivation); BMI = body mass index

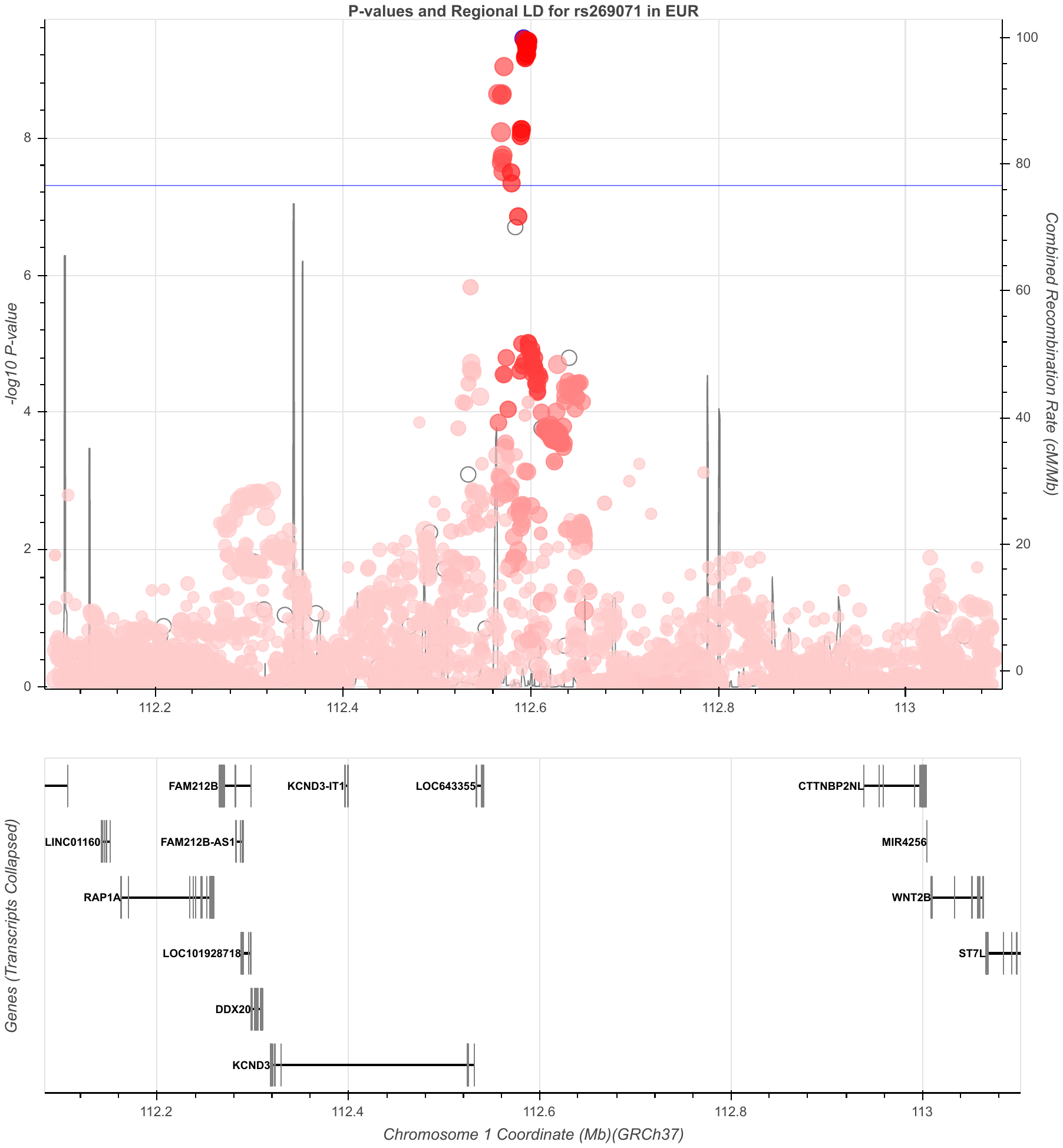

**Figure S3. CRF-vo2max chr1 rs269071 region**

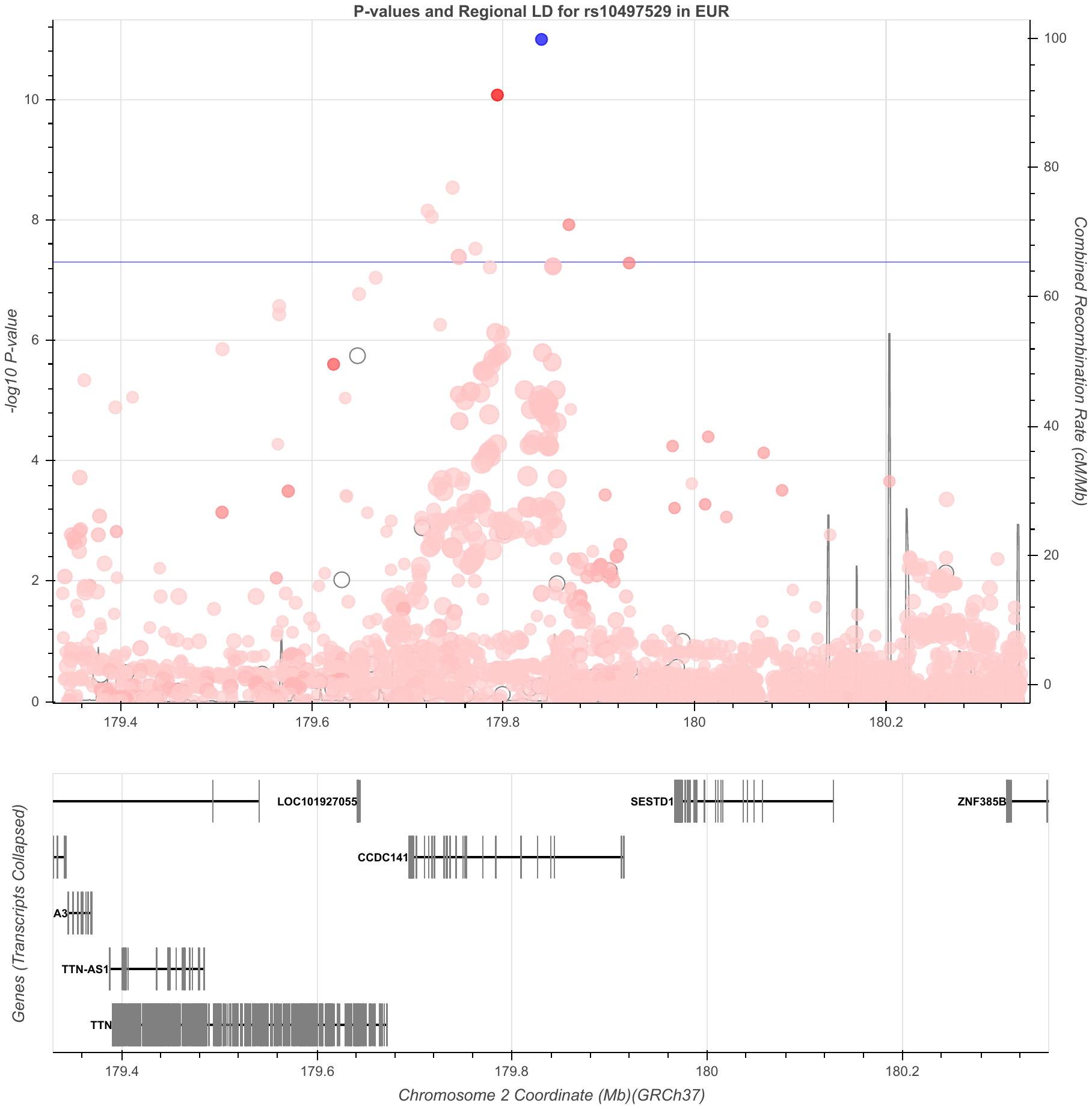

**Figure S4. CRF-vo2max chr2 rs1047529 region**

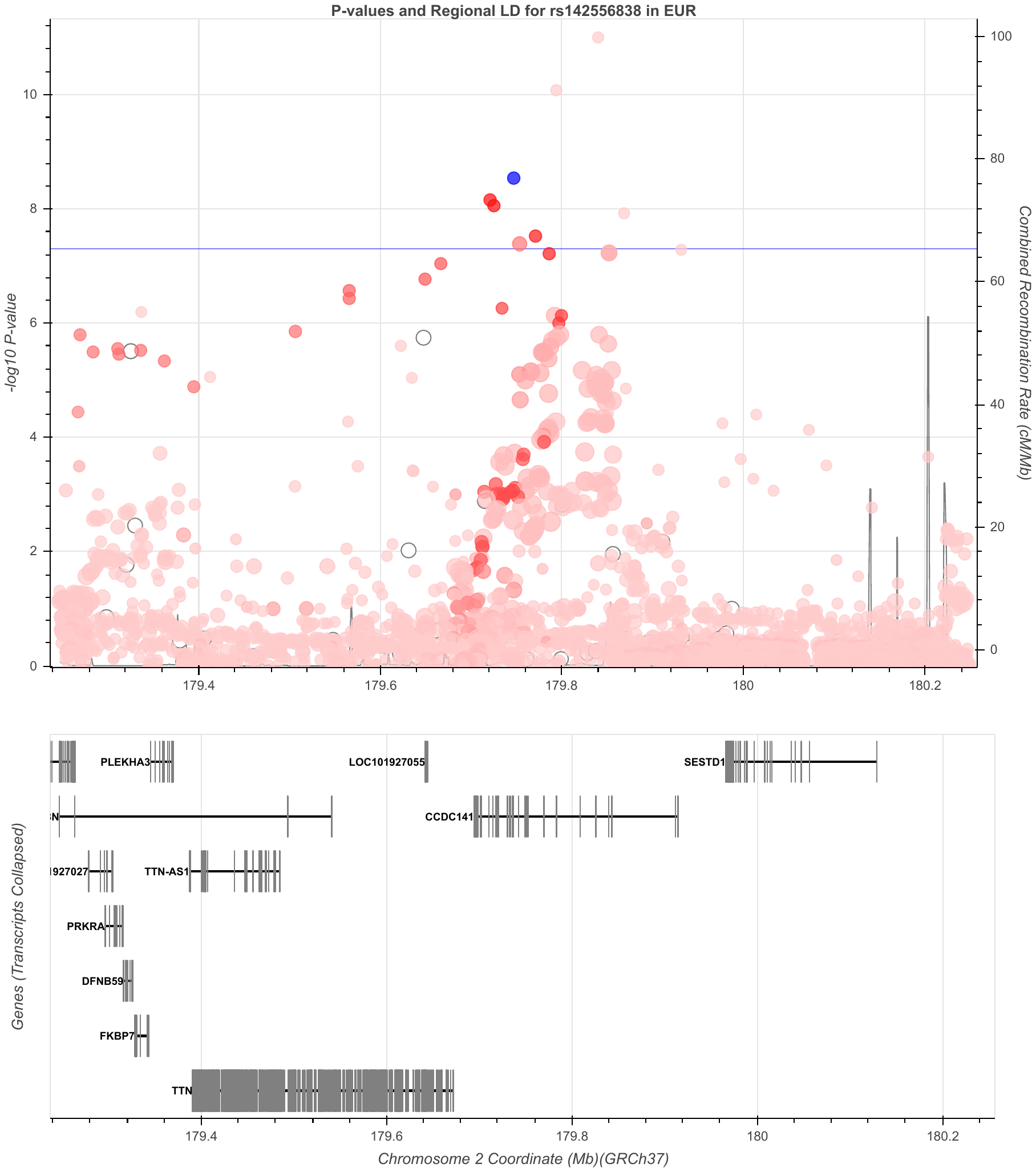
 **Figure S5. CRF-vo2max chr2 rs142556838 region**

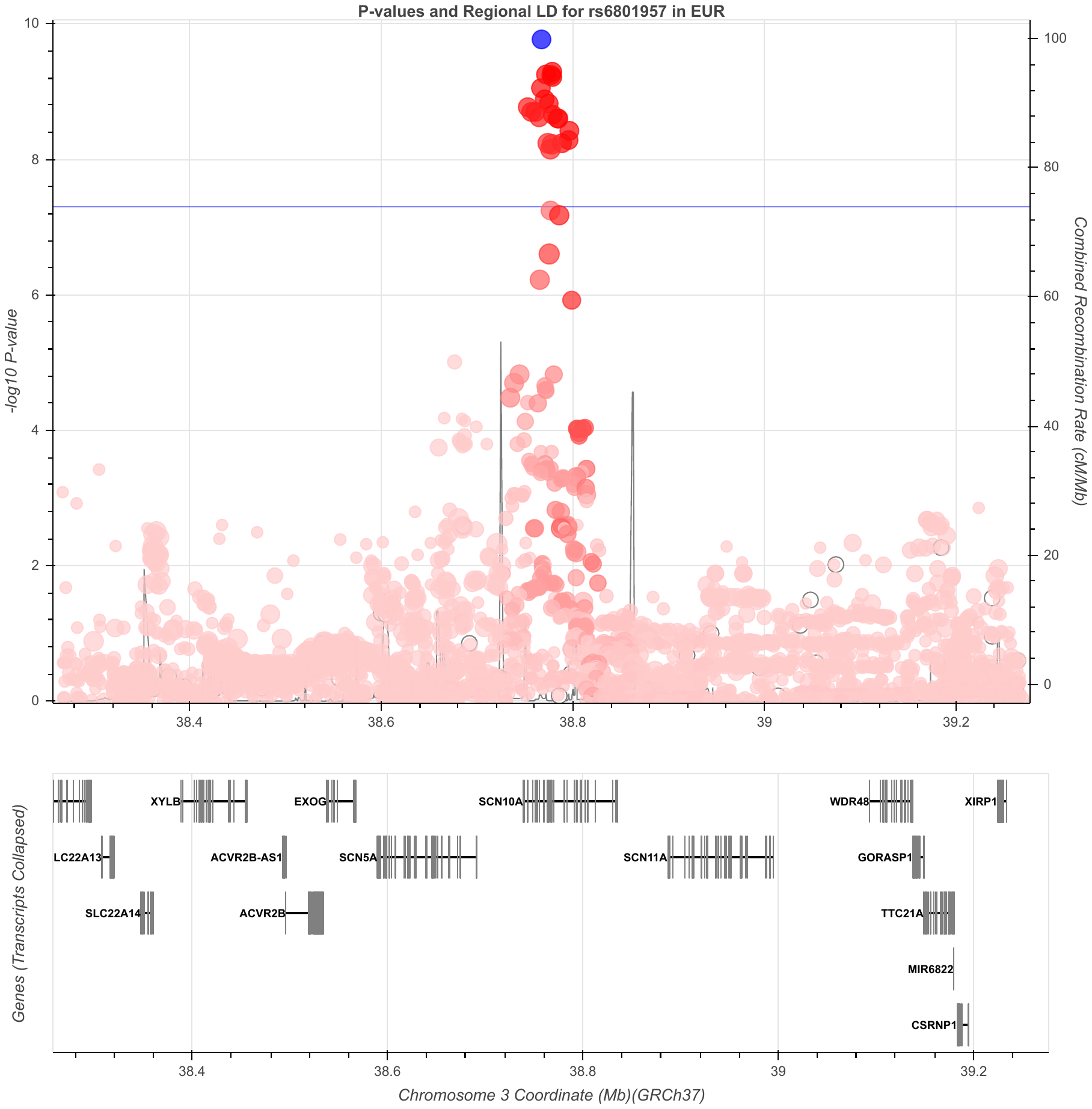
 **Figure S6. CRF-vo2max chr3 rs6801957 region**
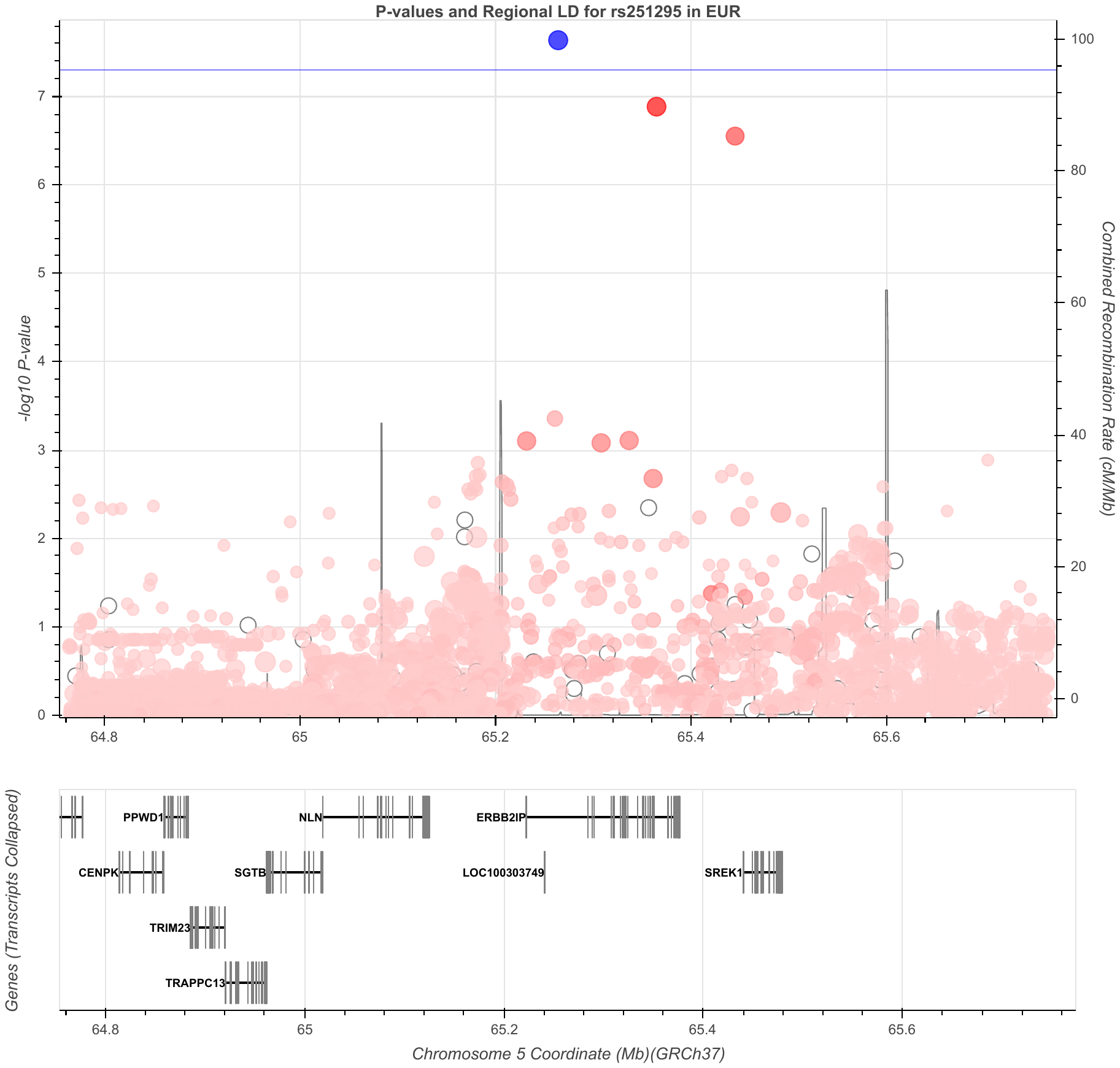
 **Figure S7. CRF-vo2max chr5 rs251295 region**
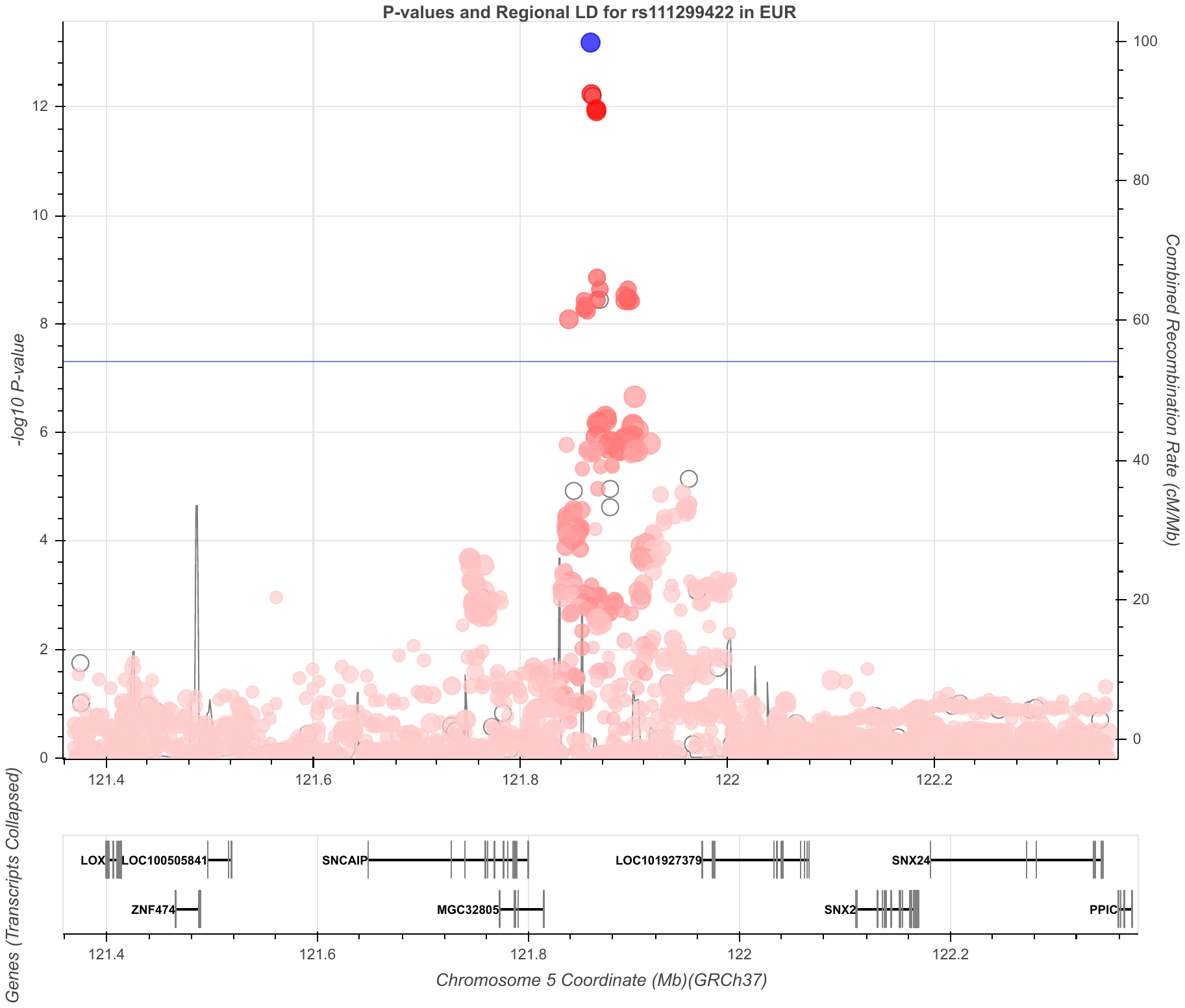
 **Figure S8. CRF-vo2max chr5 rs111299422 region**

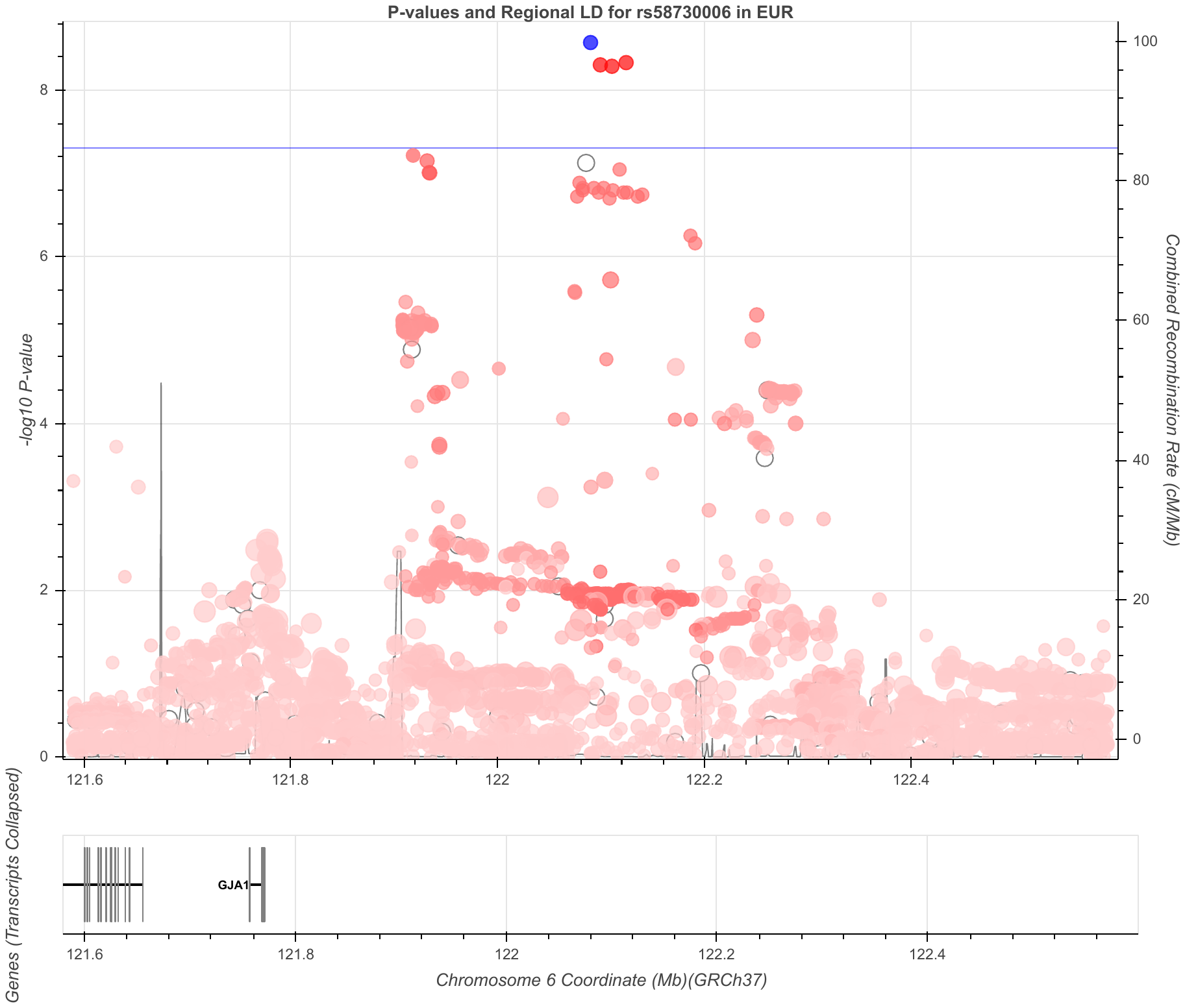
 **Figure S9. CRF-vo2max chr6 rs58730006 region**
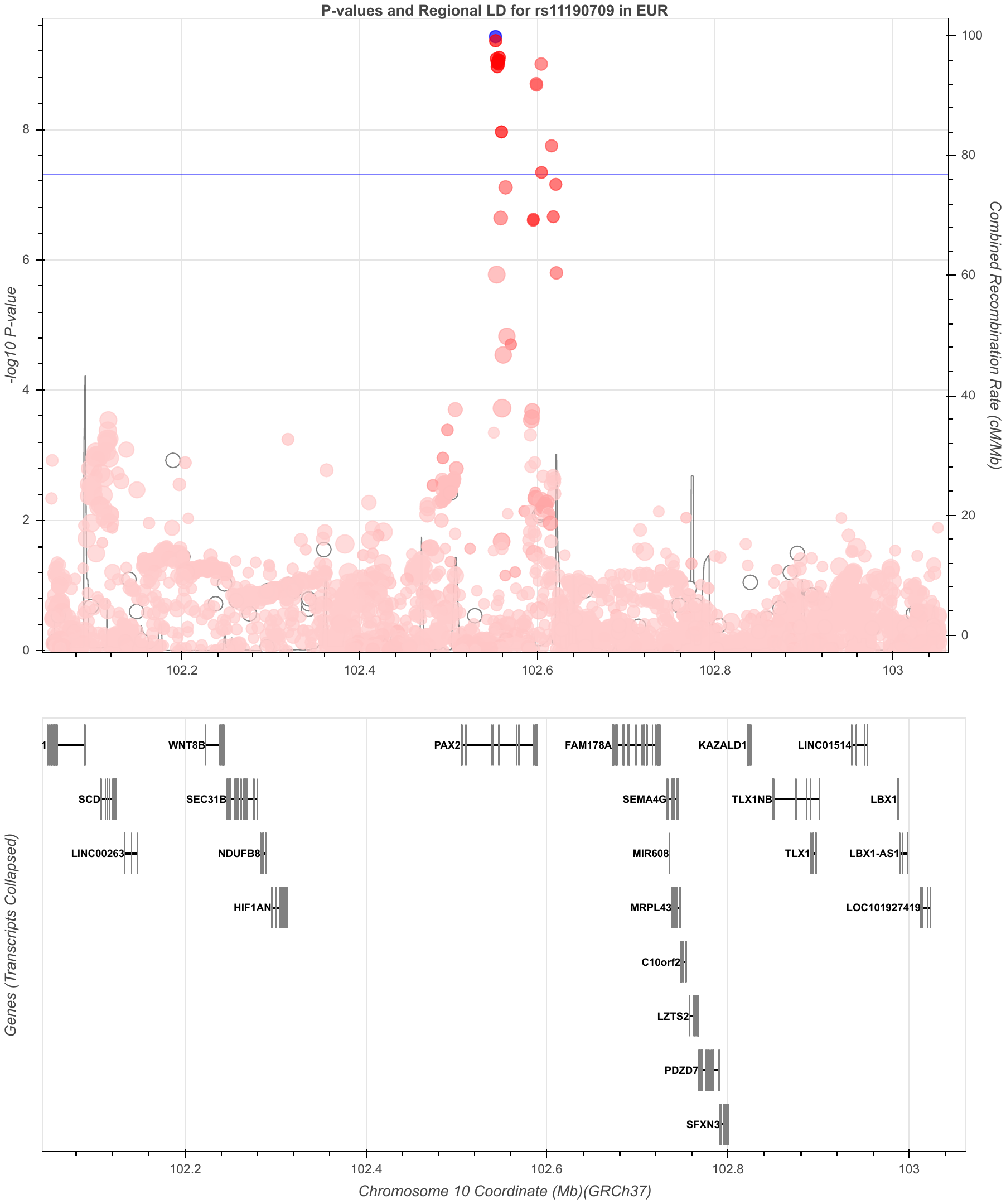
 **Figure S10. CRF-vo2max chr10 rs11190709 region**
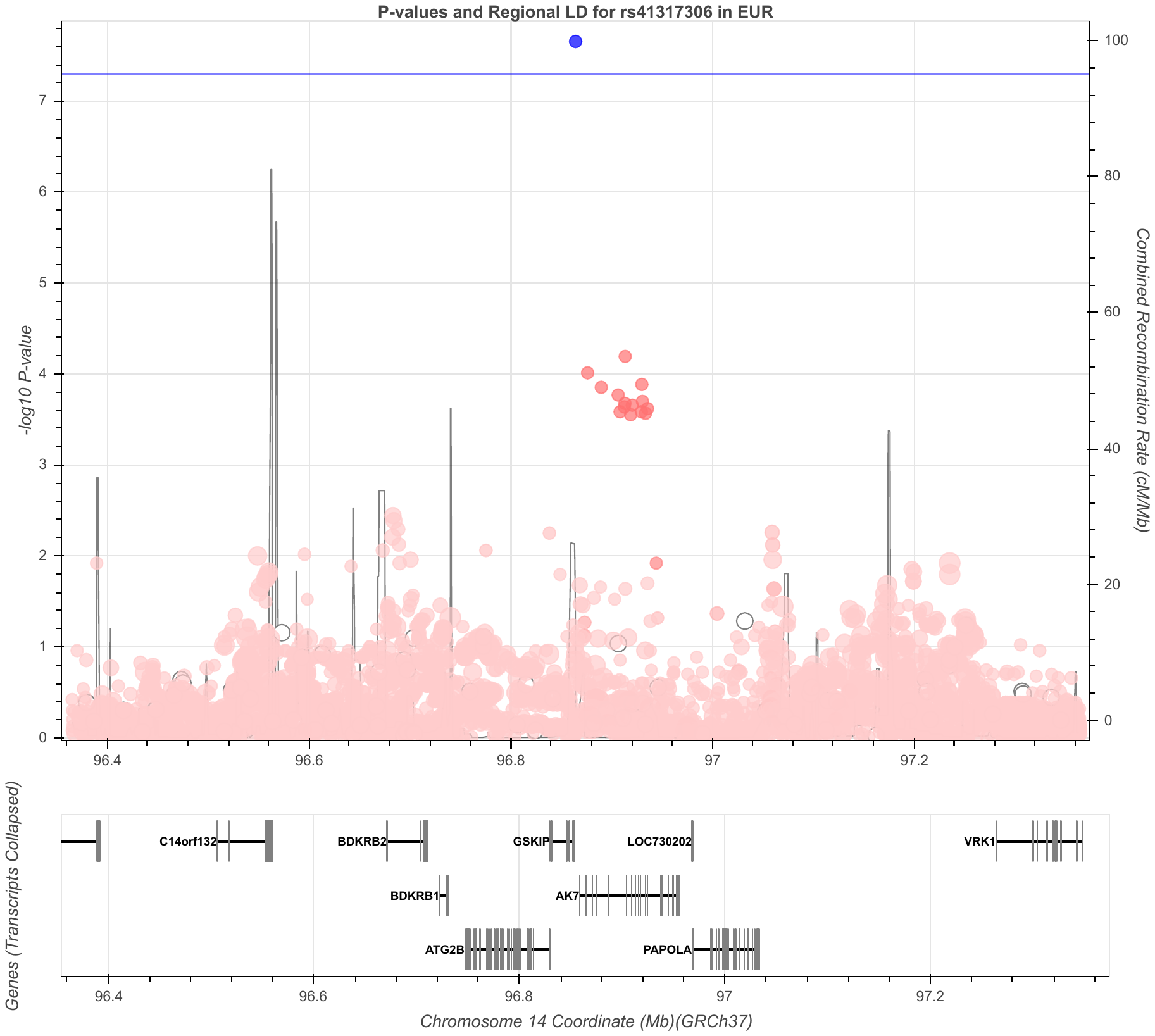
 **Figure S11. CRF-vo2max chr14 rs41317306 region (male)**
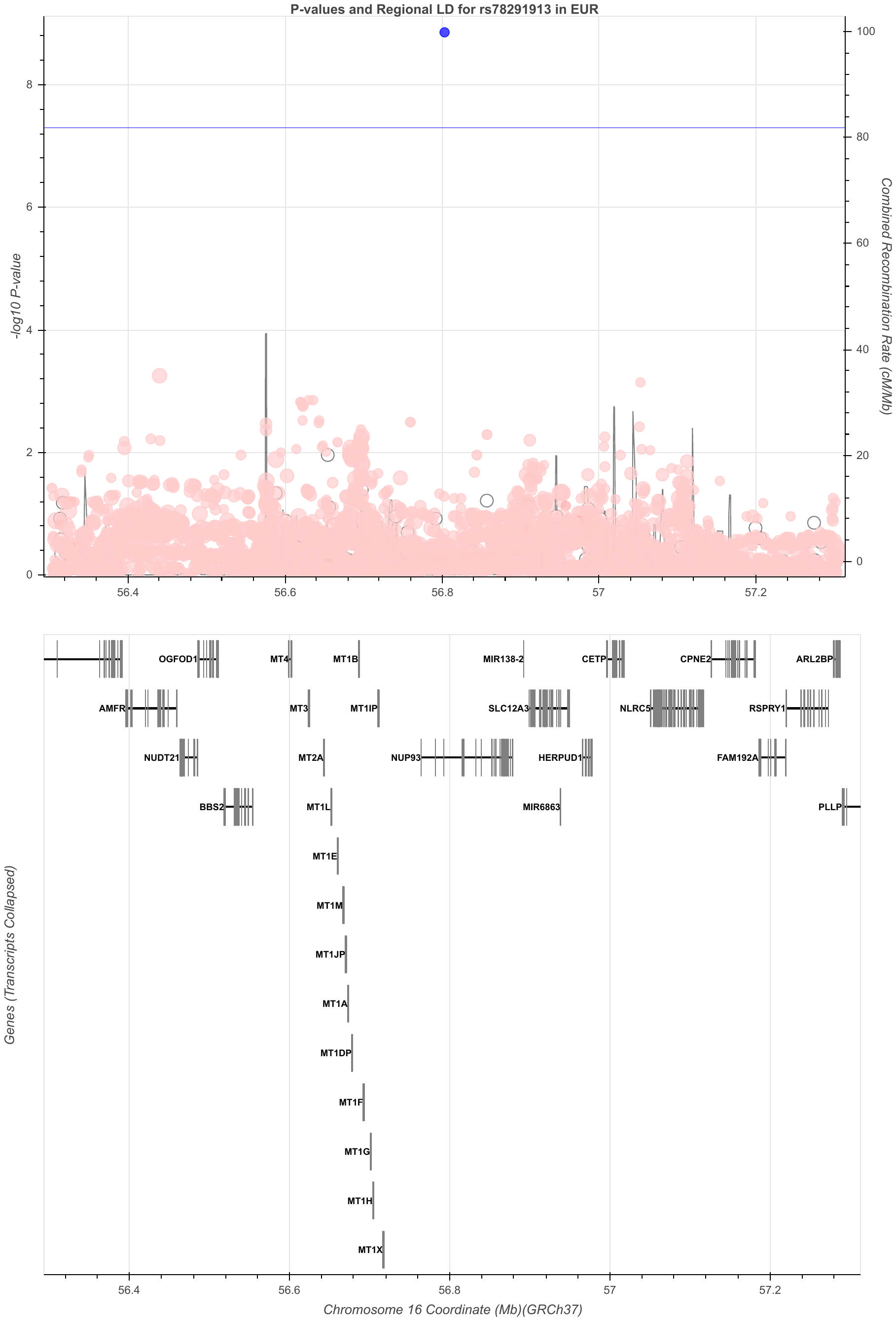
 **Figure S12. CRF-vo2max chr16 rs78291913 region**
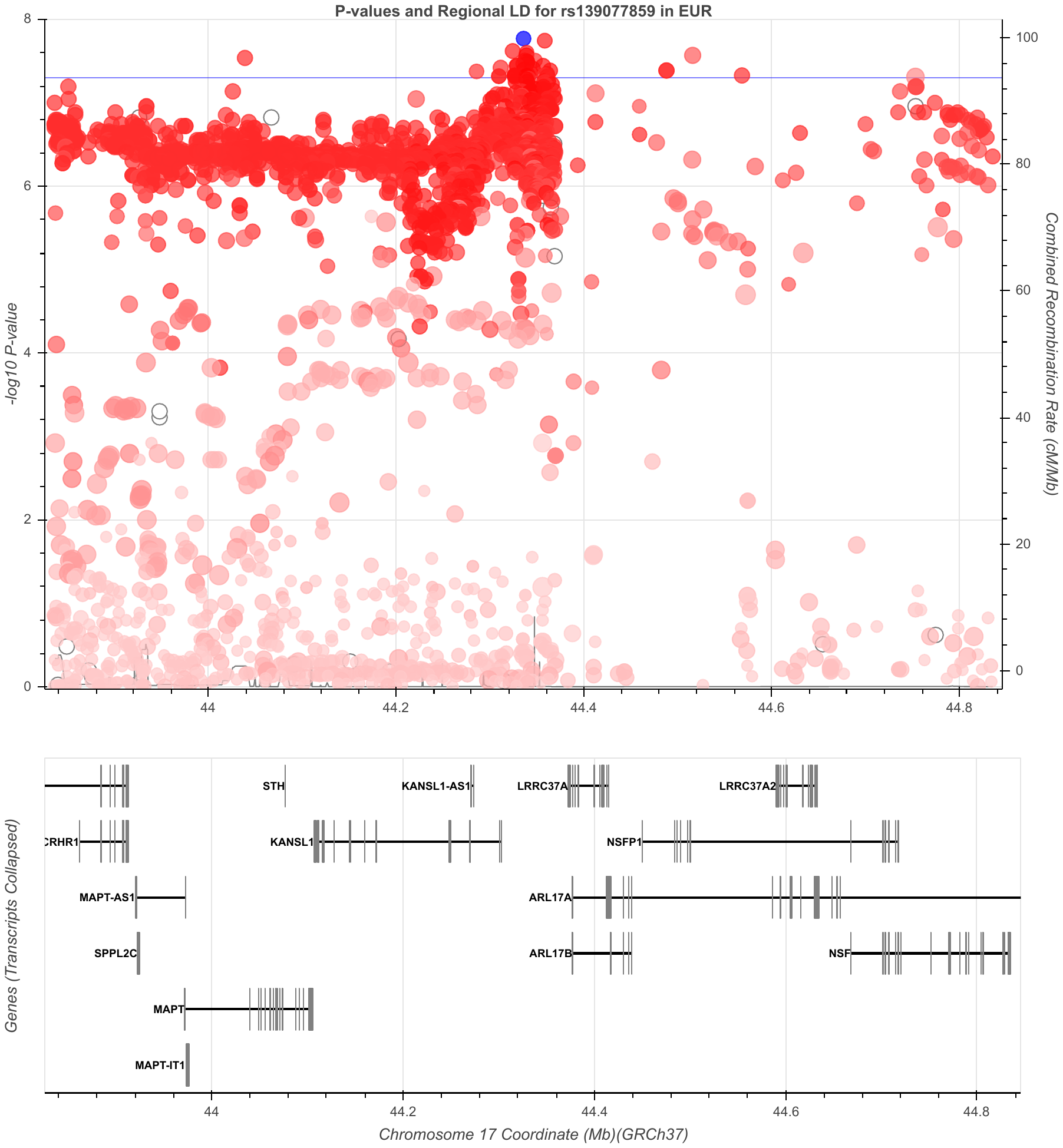
 **Figure S13. CRF-vo2max chr17 rs139077859 region**
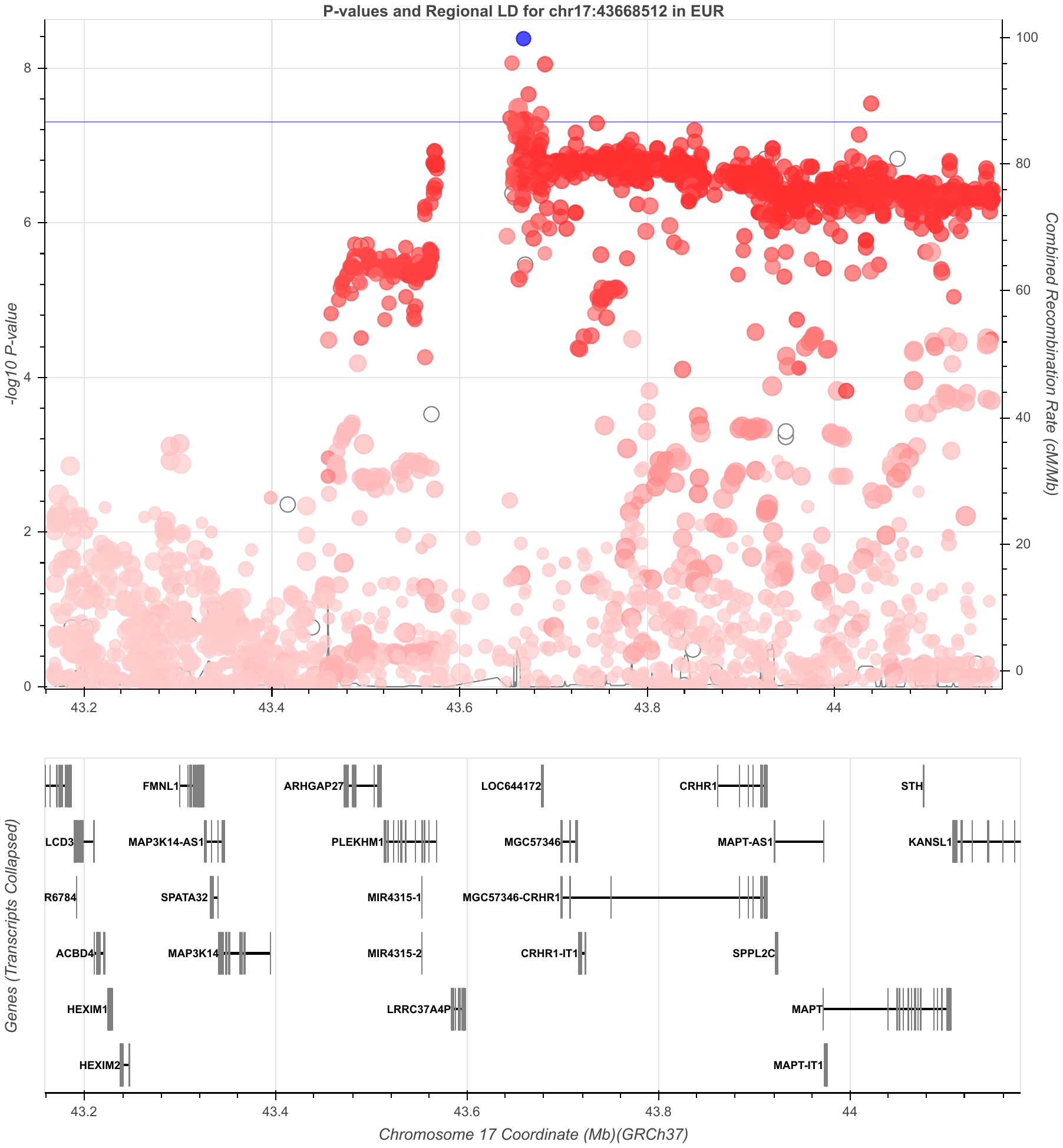
 **Figure S14. CRF-vo2max chr17:43668512 (rs527325496) region**
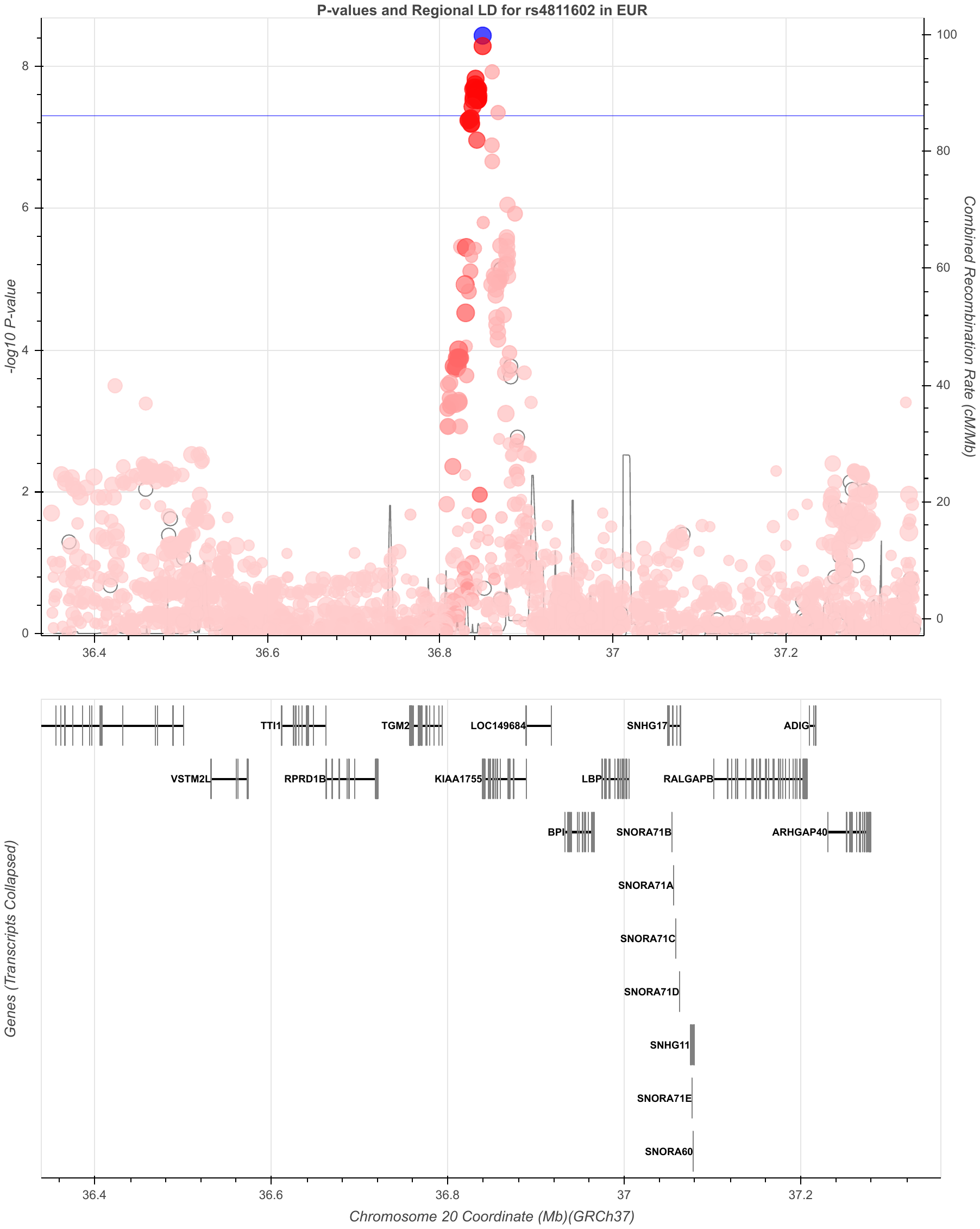
 **Figure S15. CRF-vo2max chr20 rs4811602 region**

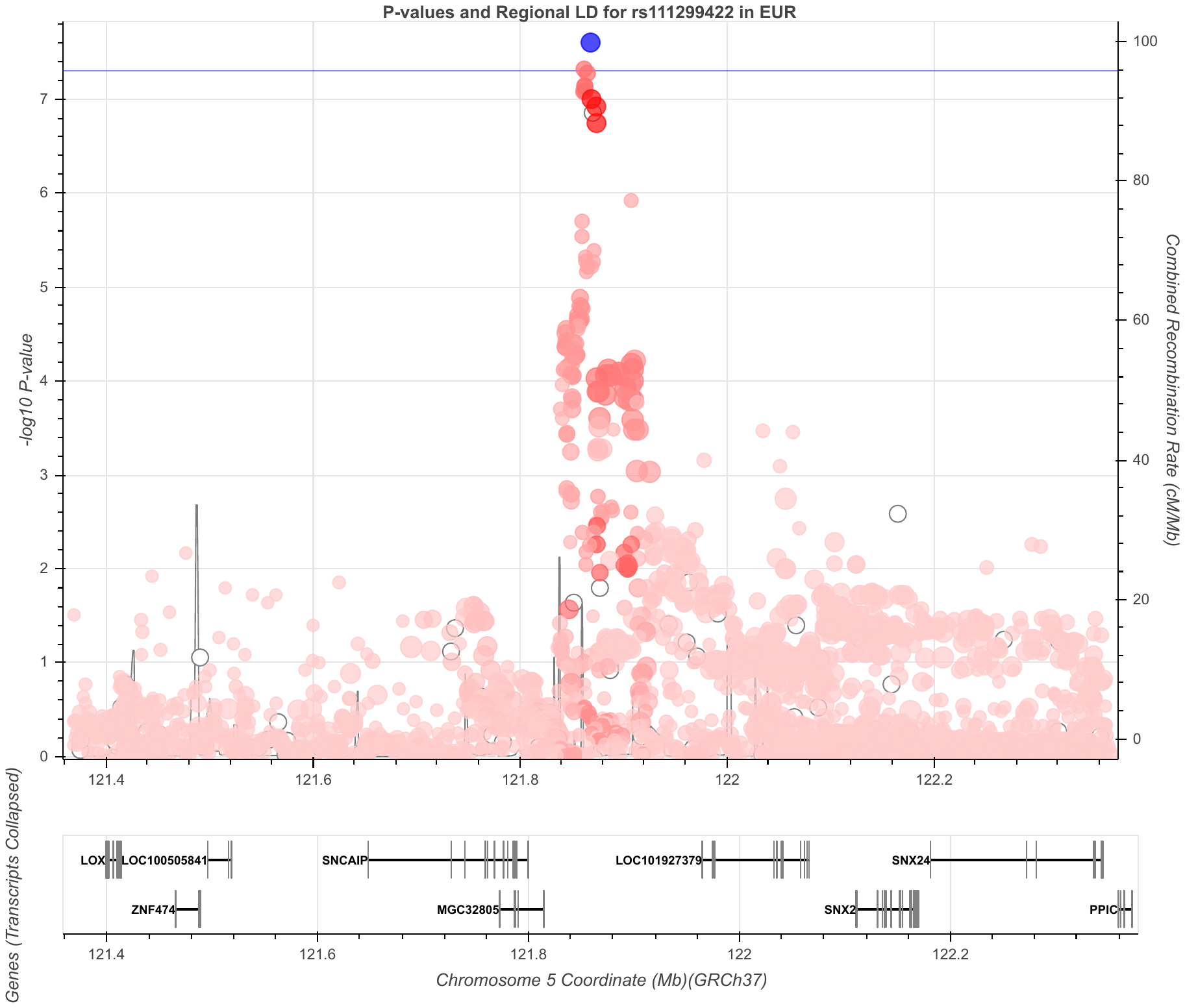
 **Figure S16. CRF-slope chr5 rs111299422 region (female)**
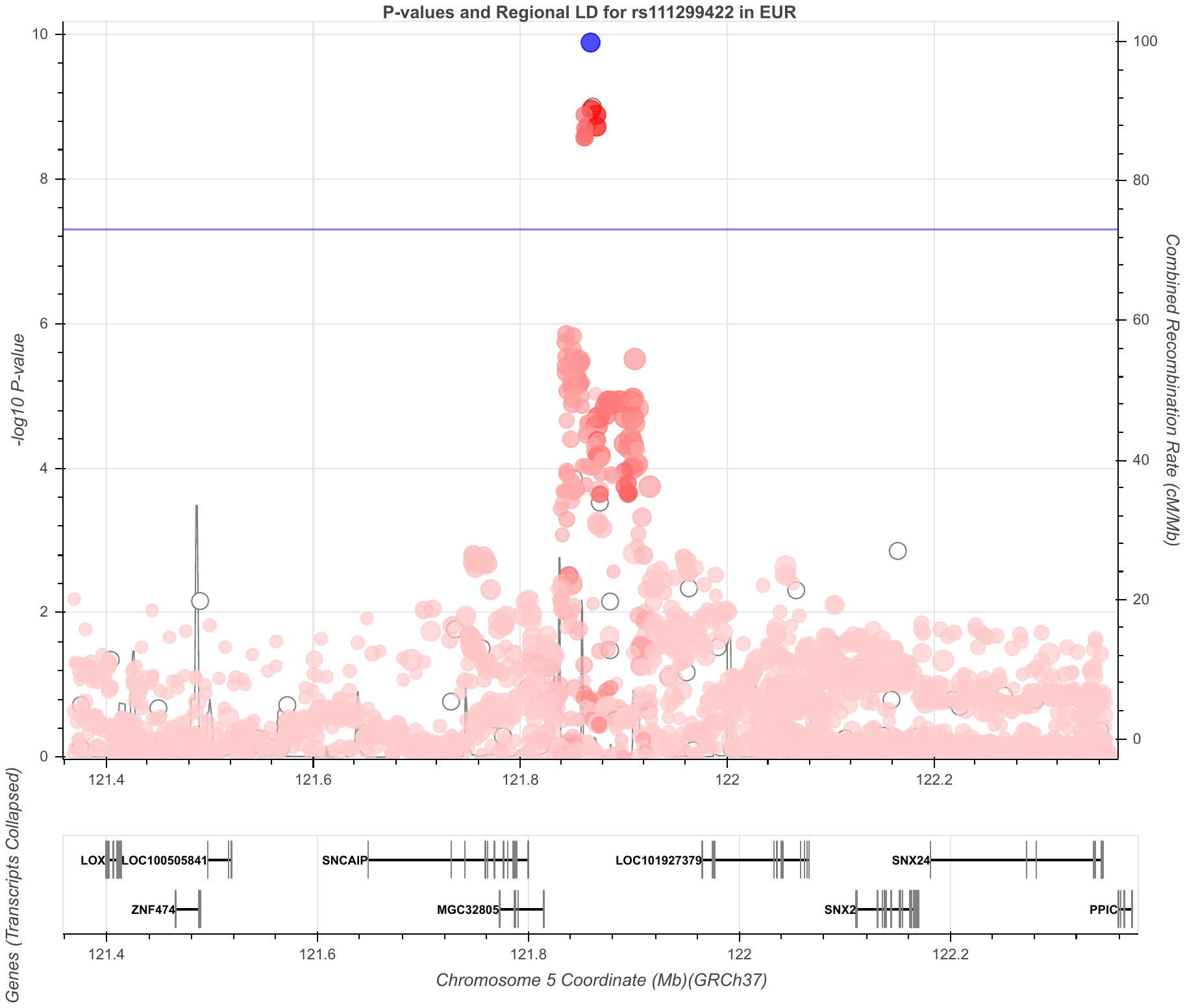
 **Figure S17. CRF-slope chr5 rs111299422 region**
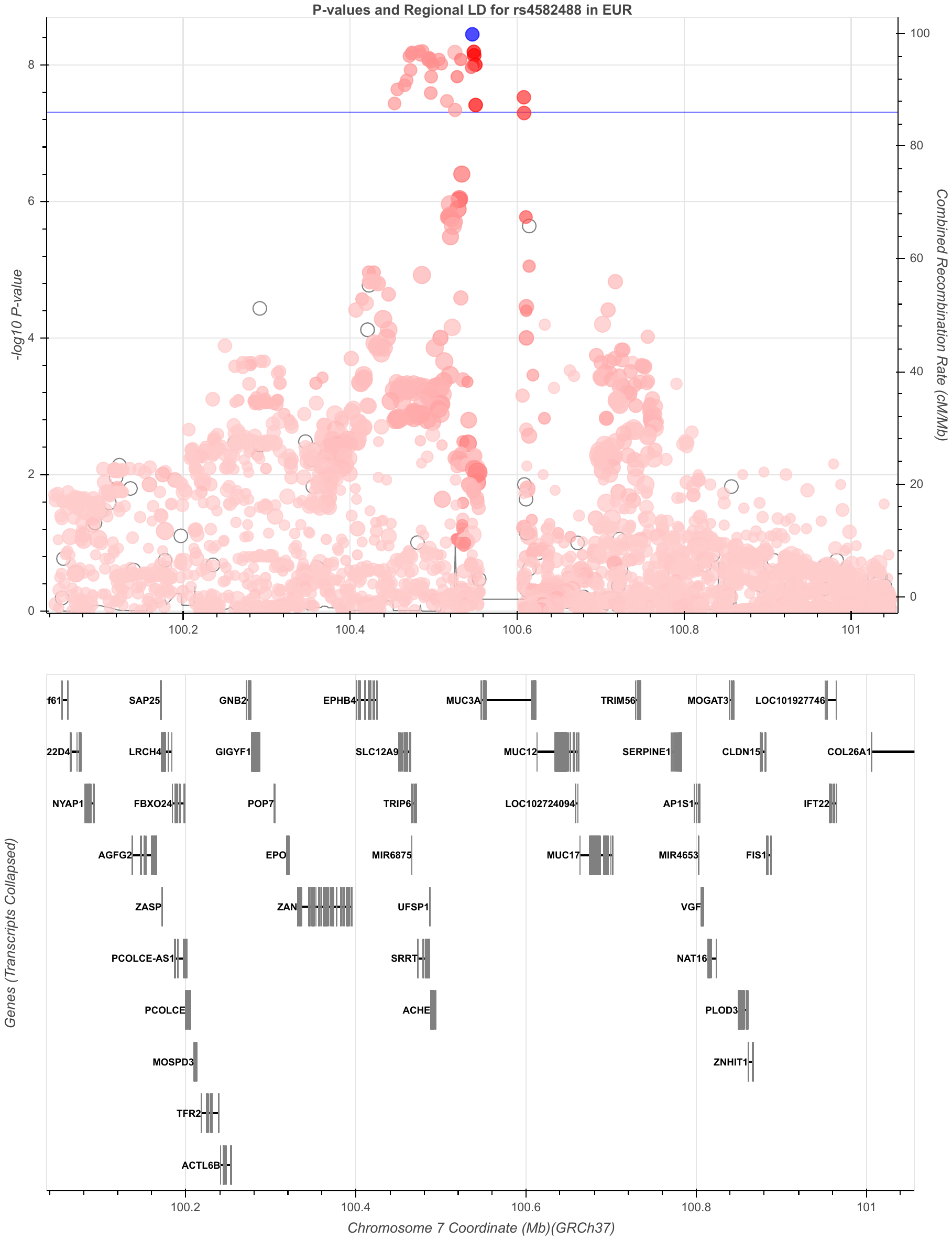
 **Figure S18. CRF-slope chr7 rs4582488 region**
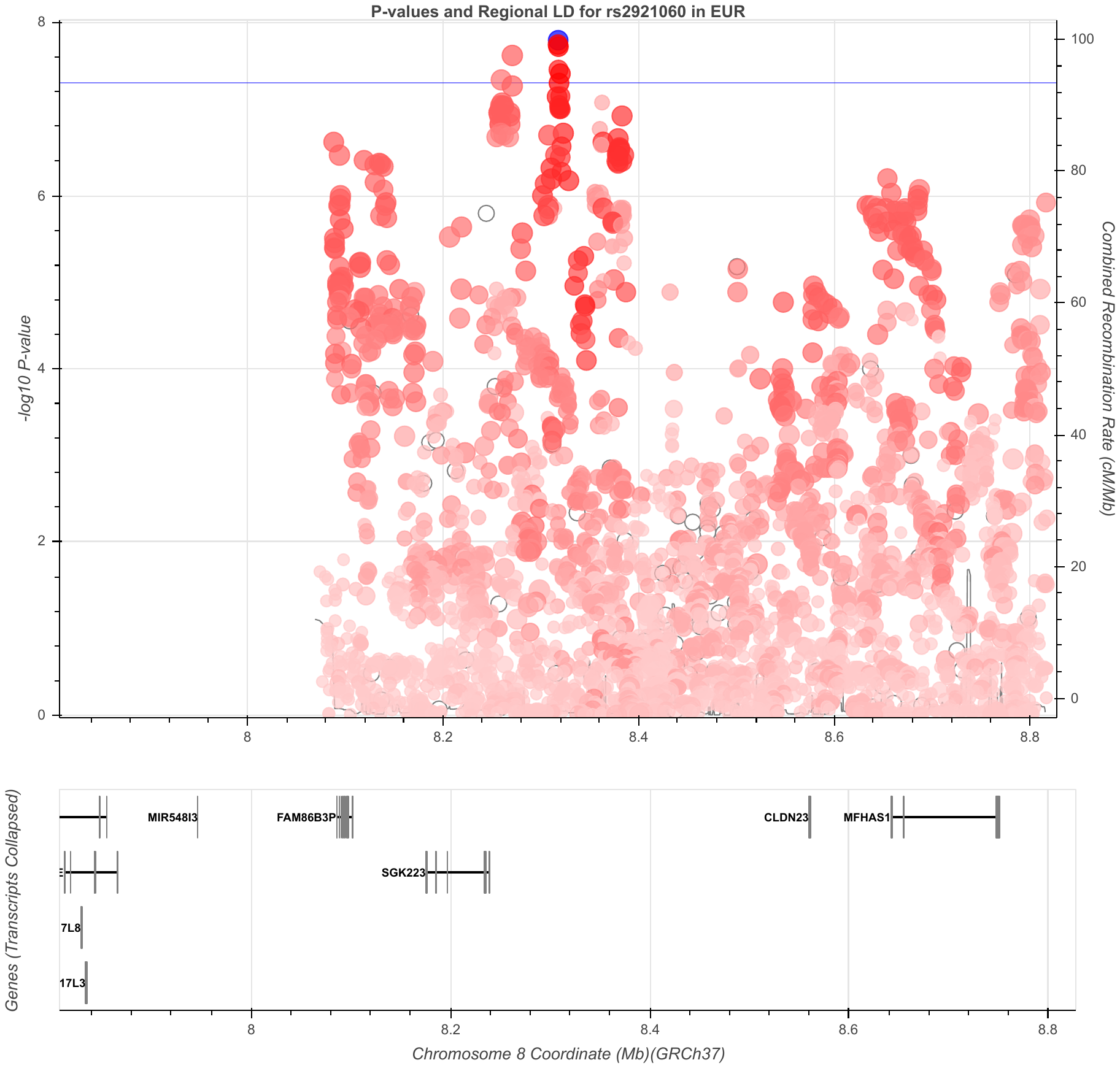
 **Figure S19. CRF-slope chr8 rs2921060 region**
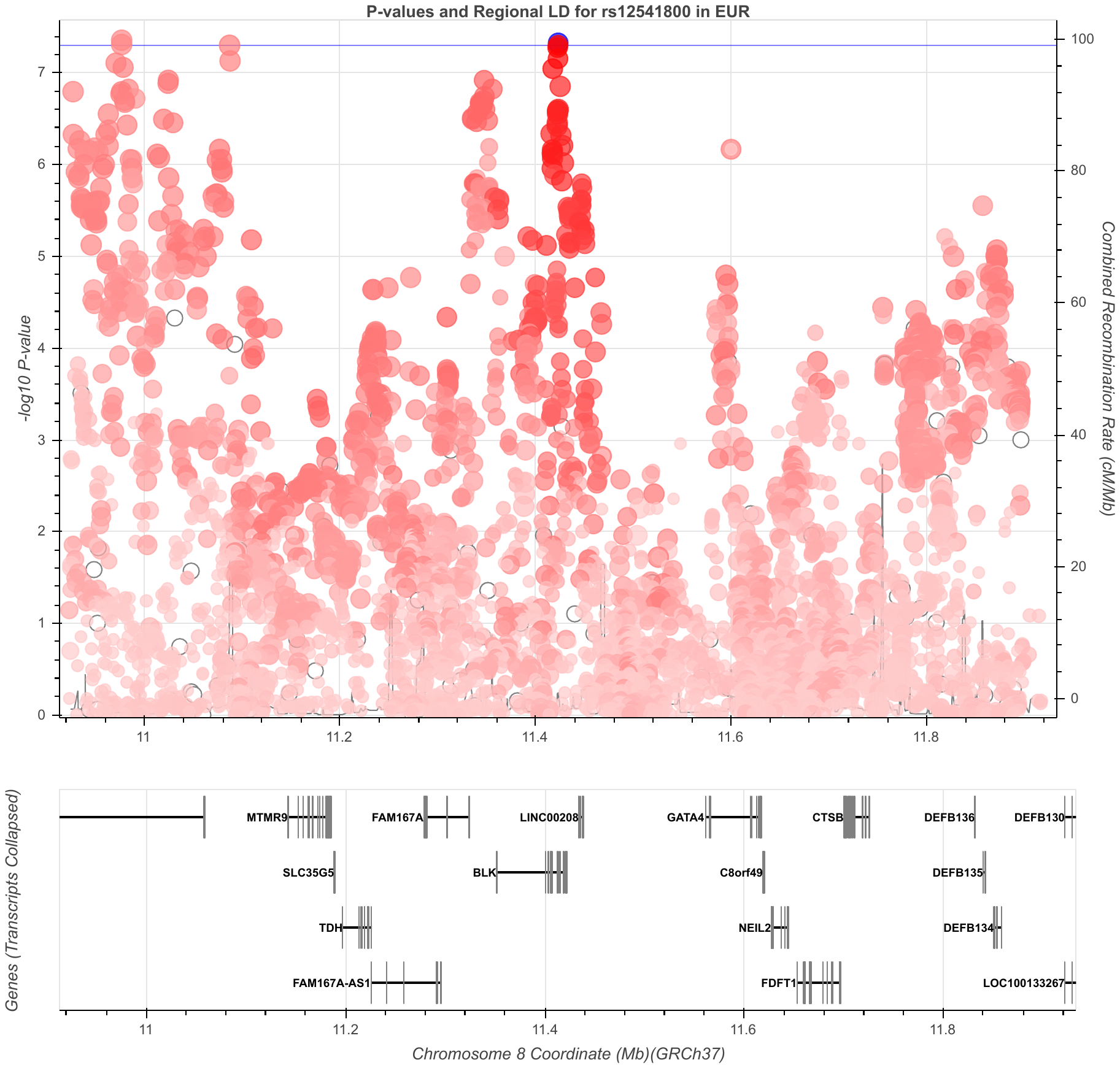
 **Figure S20. CRF-slope chr8 rs12541800 region**
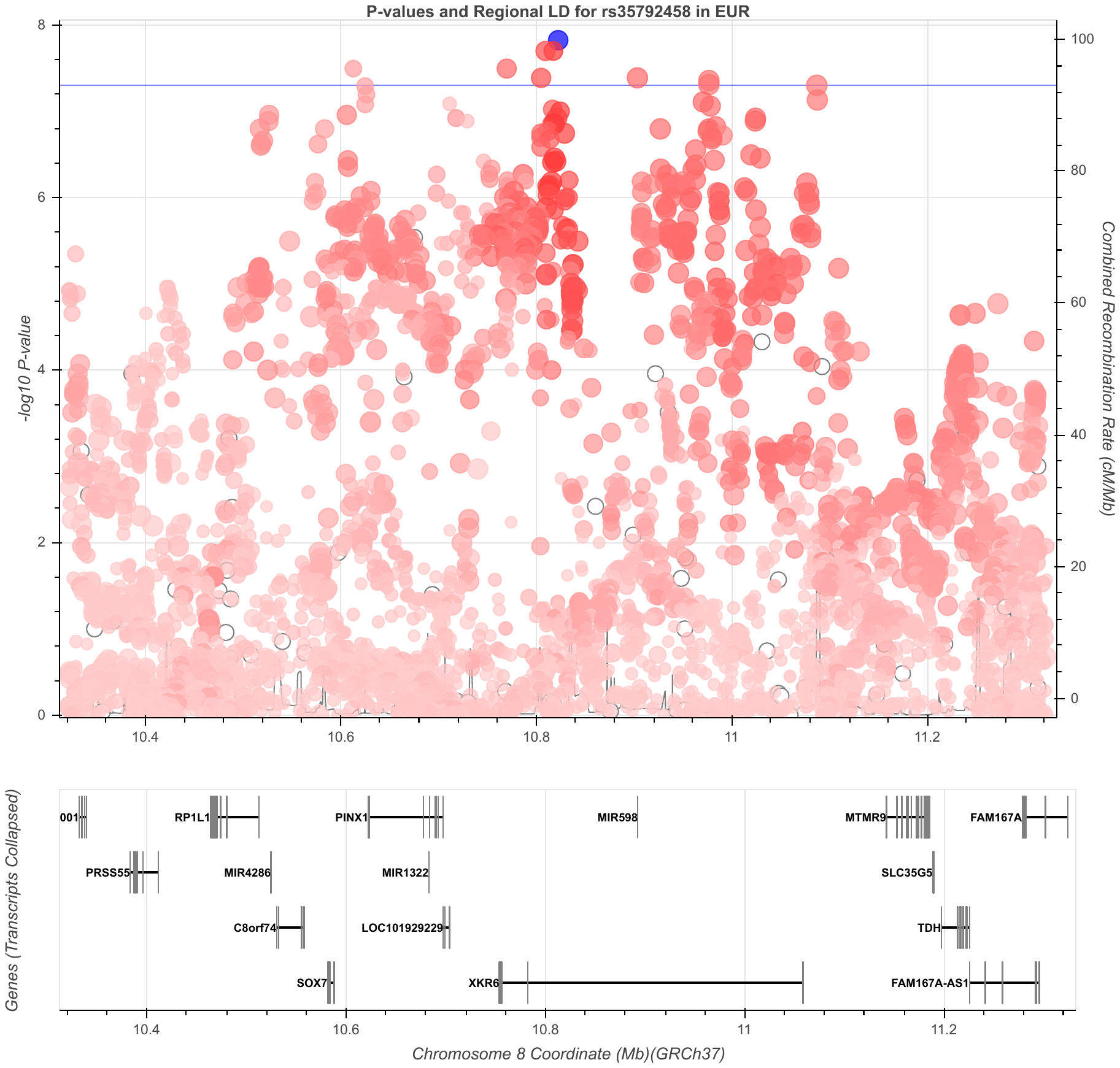
 **Figure S21. CRF-slope chr8 rs35792458 region**
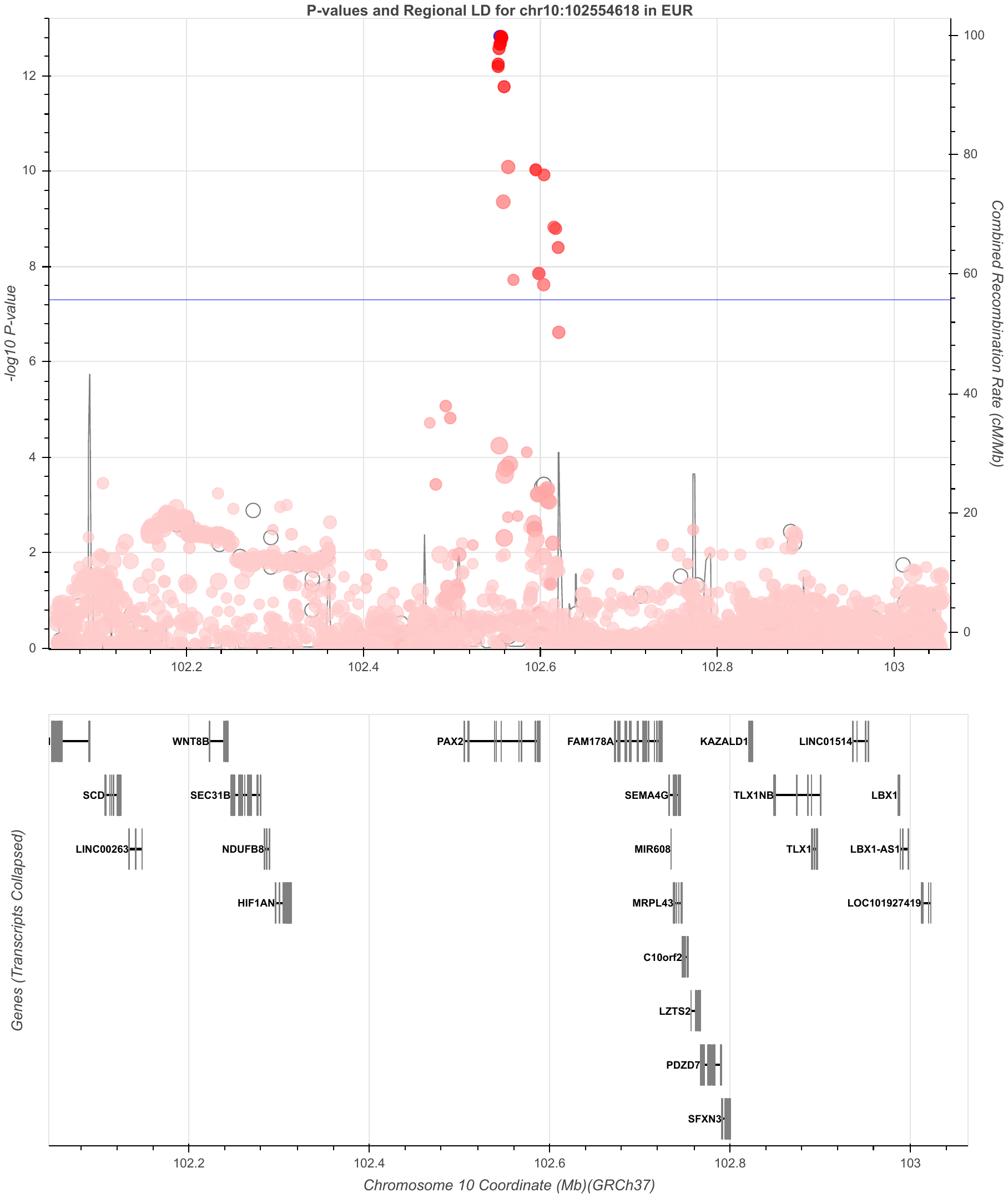
 **Figure S22. CRF-slope chr10 10:102554618_AT_A region**
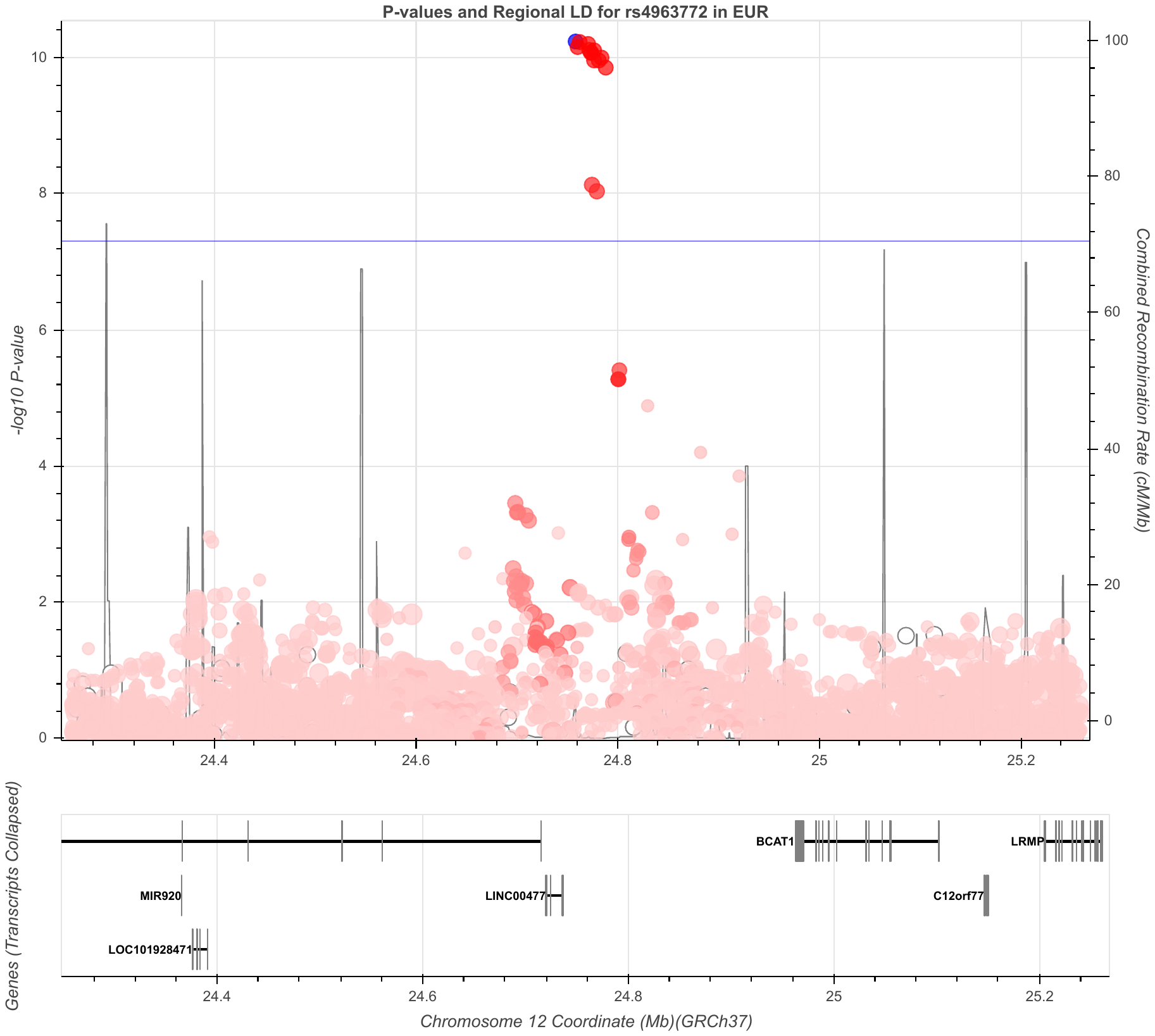
 **Figure S23. CRF-slope chr12 rs4963772 region**
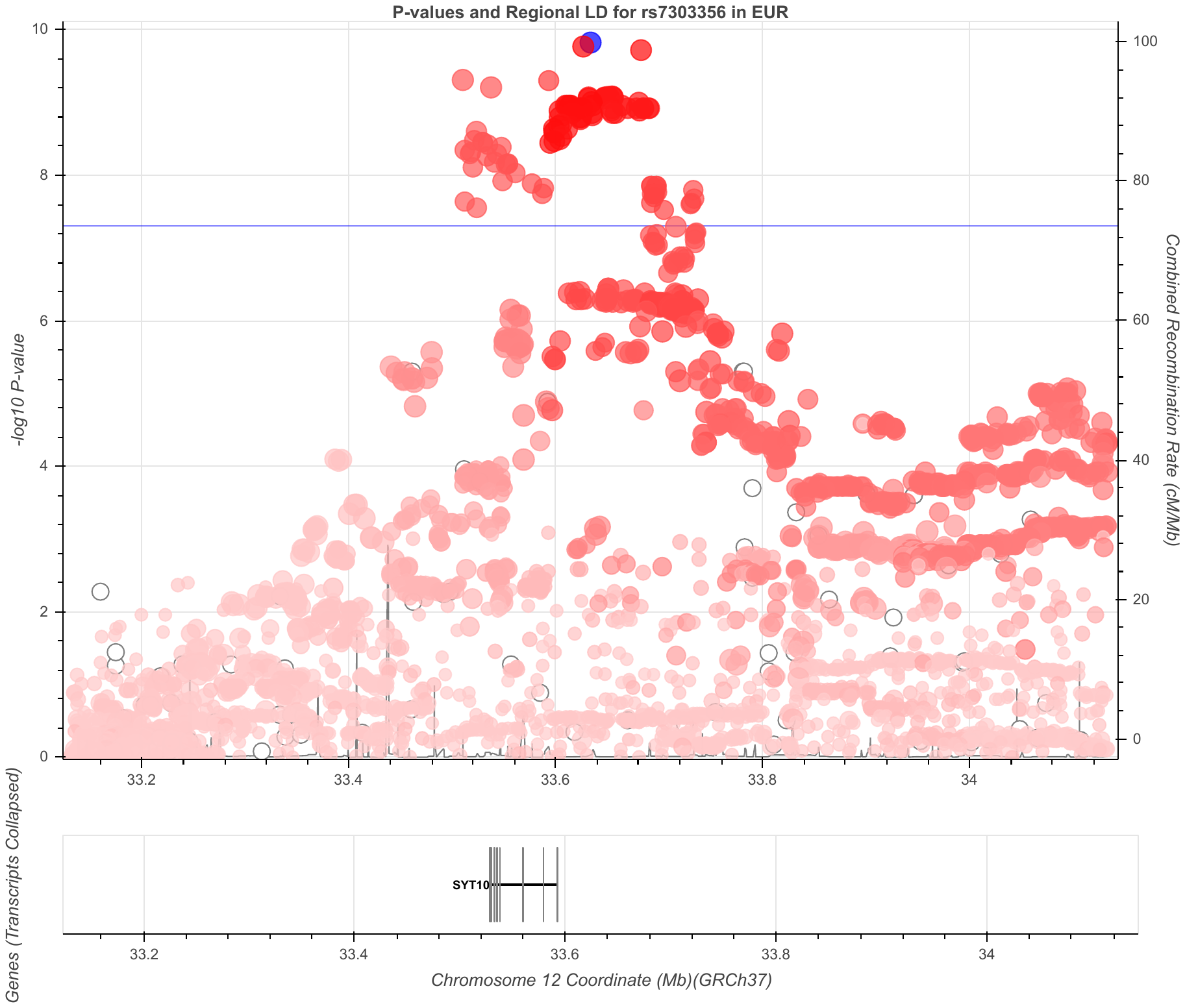
 **Figure S24. CRF-slope chr12 rs7303356 region**
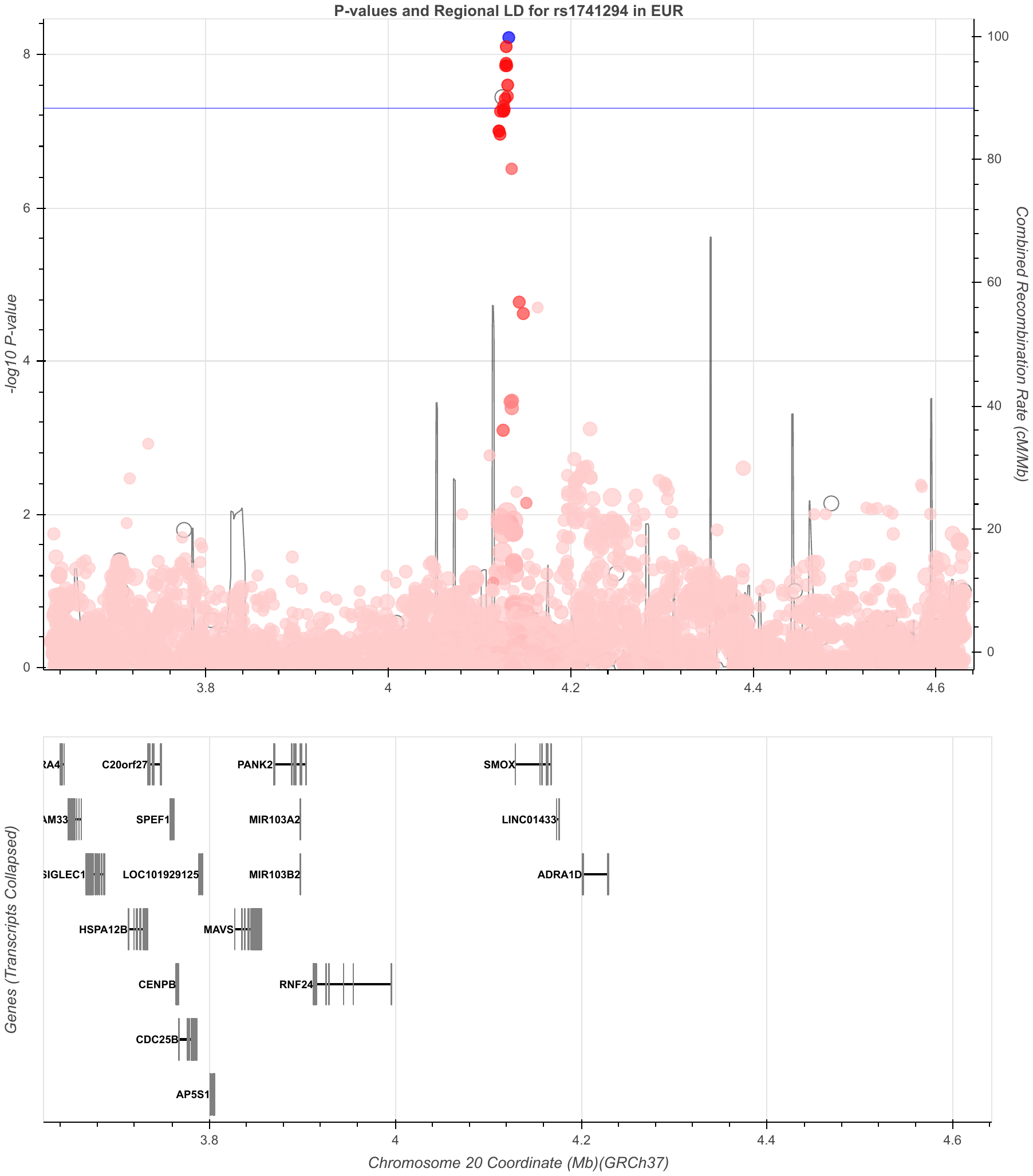
 **Figure S25. CRF-slope chr20 rs1741294 region (male)**

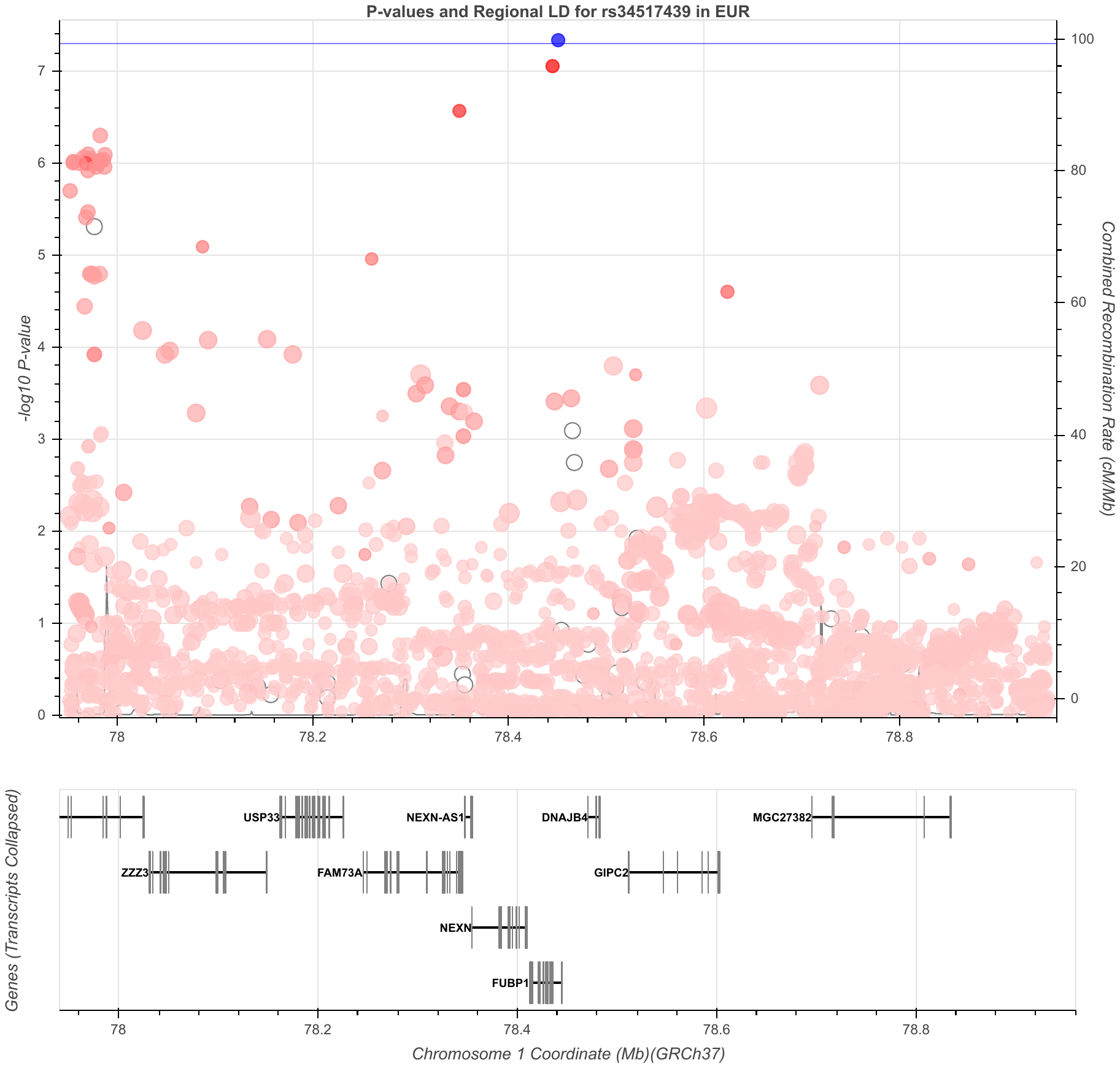
 **Figure S26. PA chr1 rs34517439 region**
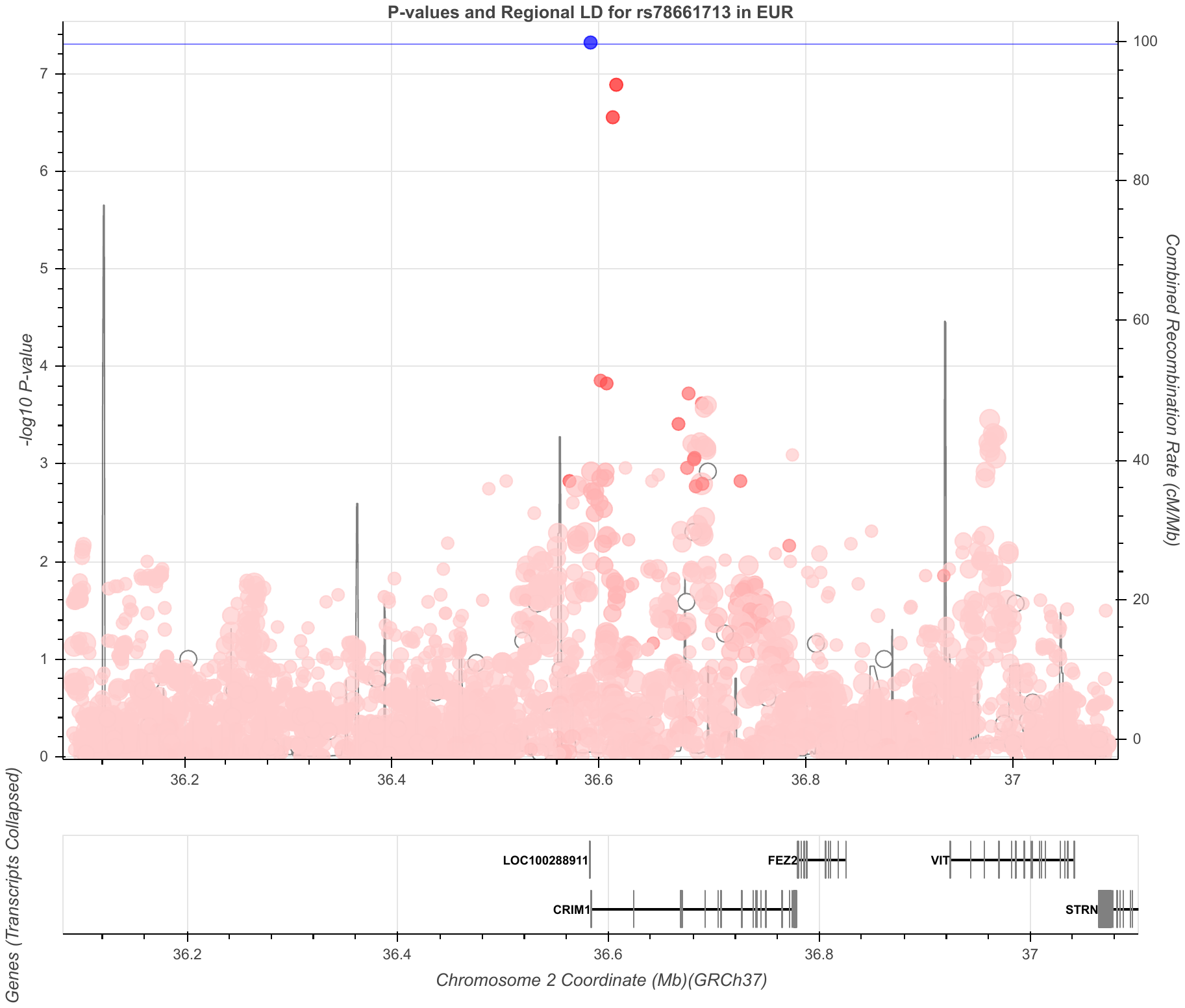
 **Figure S27. PA chr2 rs78661713 region (male)**
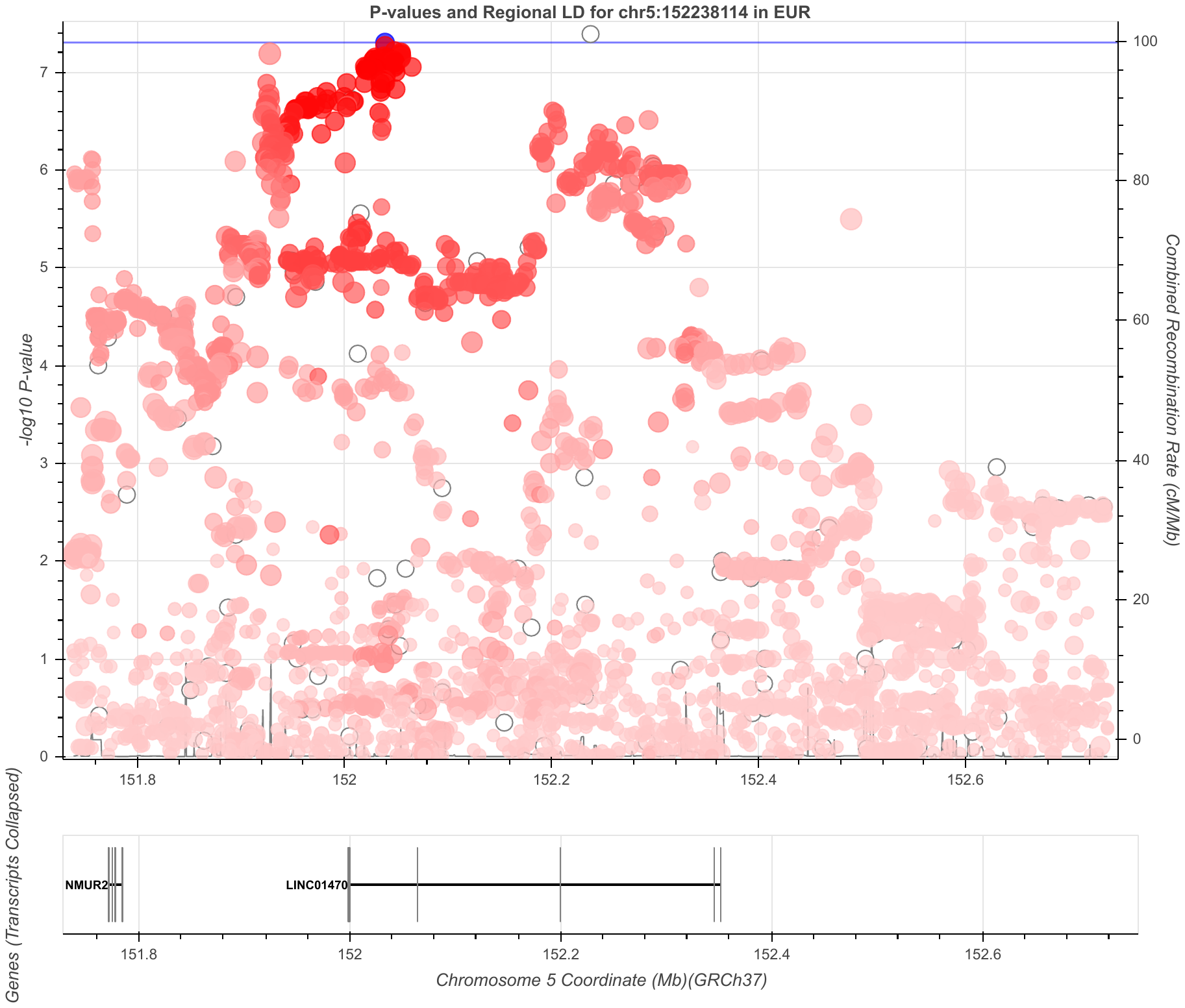
 **Figure S28. PA chr5 5:1527381114_TTTTTTTTTTTTC_T region**
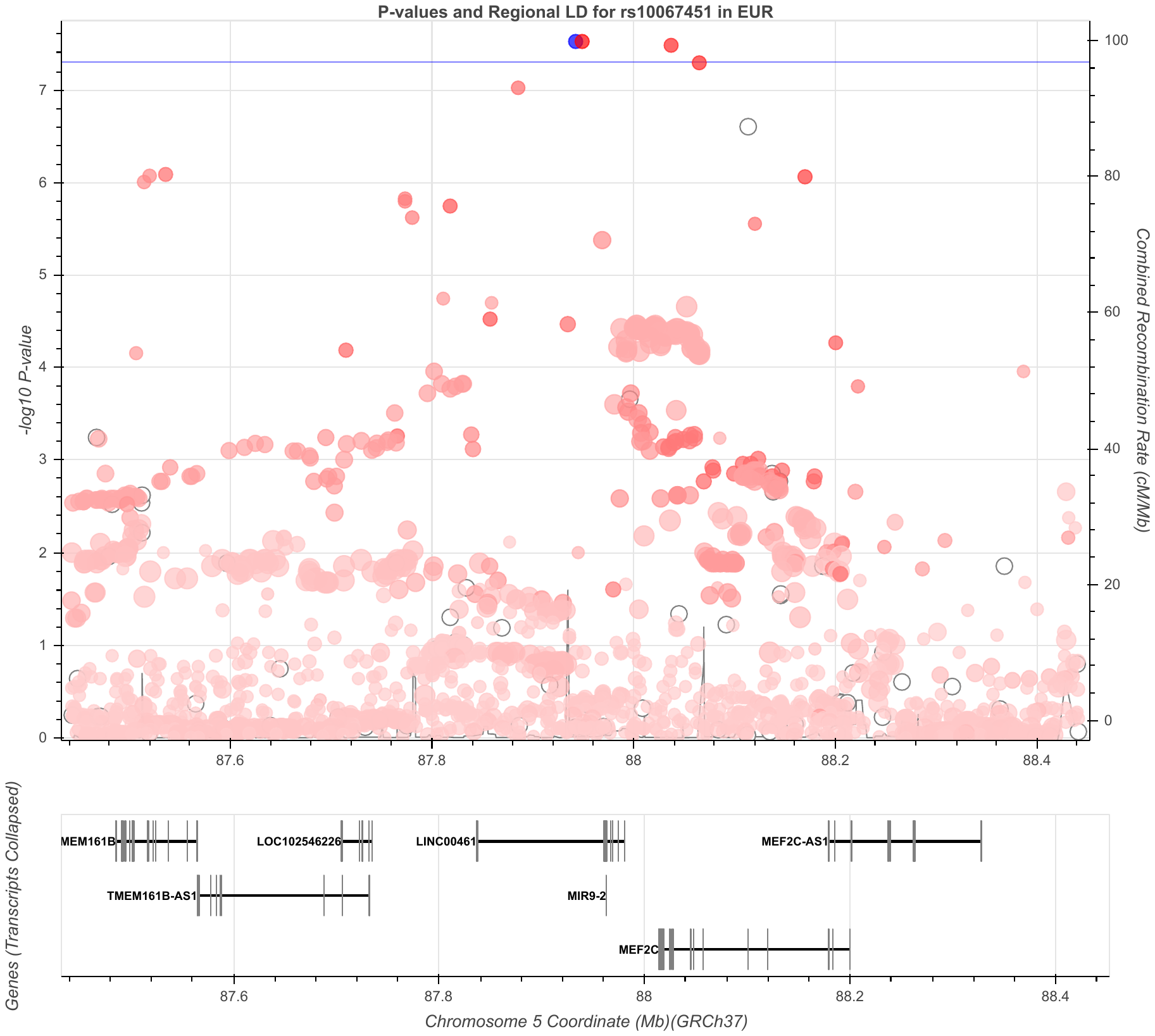
 **Figure S29. PA chr5 rs10067451 region**
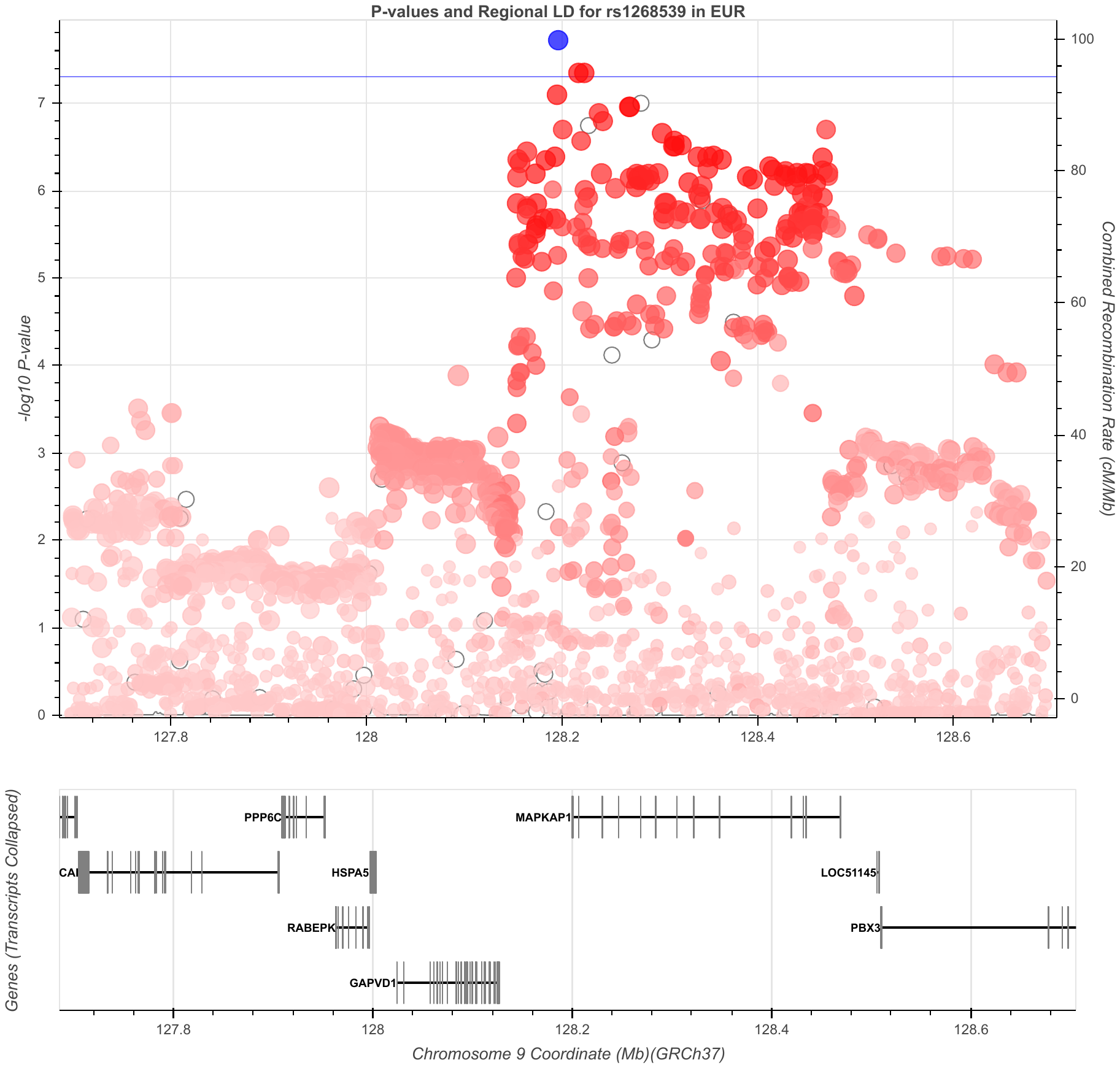
 **Figure S30. PA chr9 rs1268539 region**
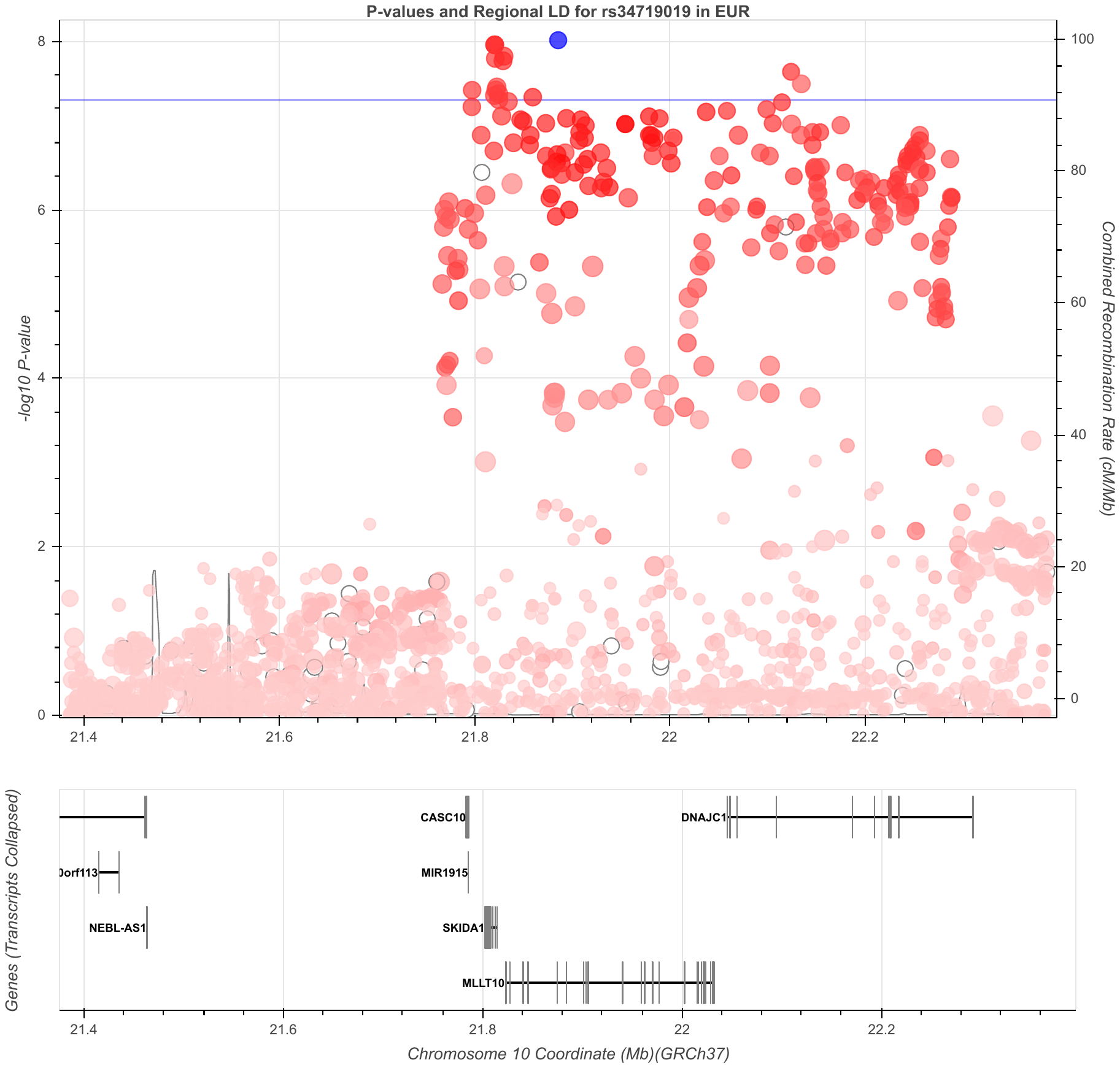
 **Figure S31. PA chr10 rs34719019 region**
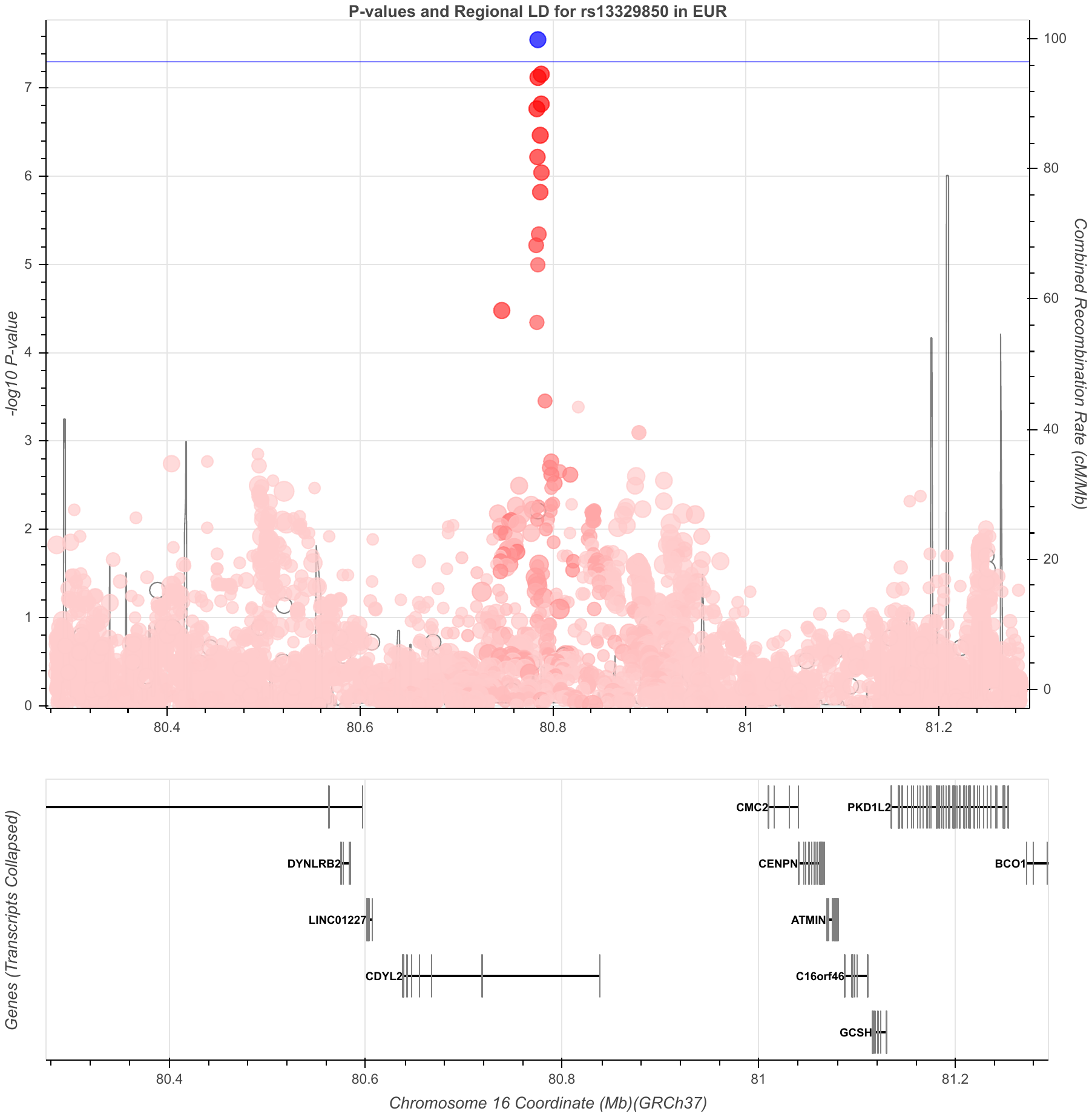
 **Figure S32. PA chr16 rs13329850 region (female)**
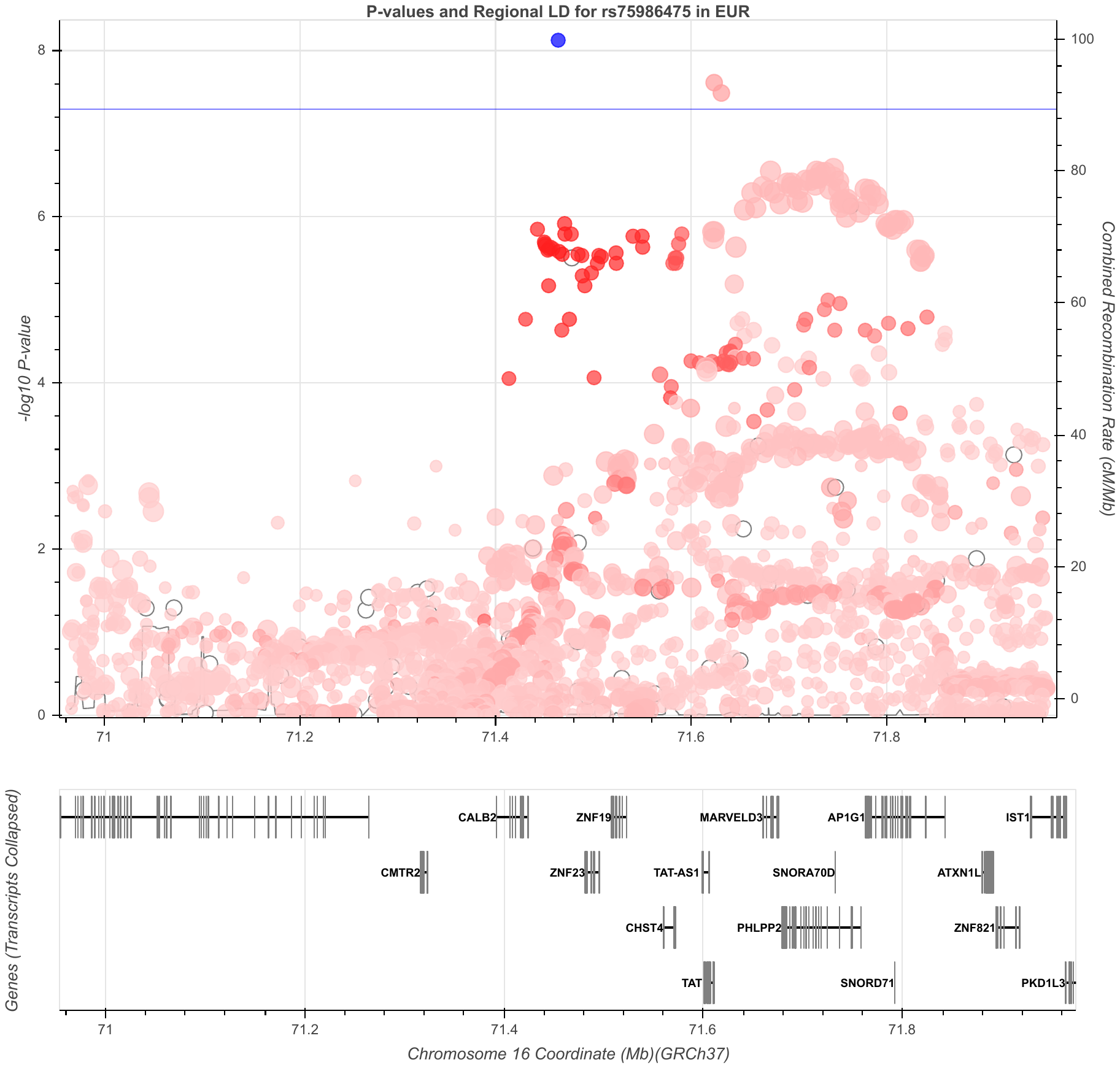
 **Figure S33. PA chr16 rs75986475 region (female)**
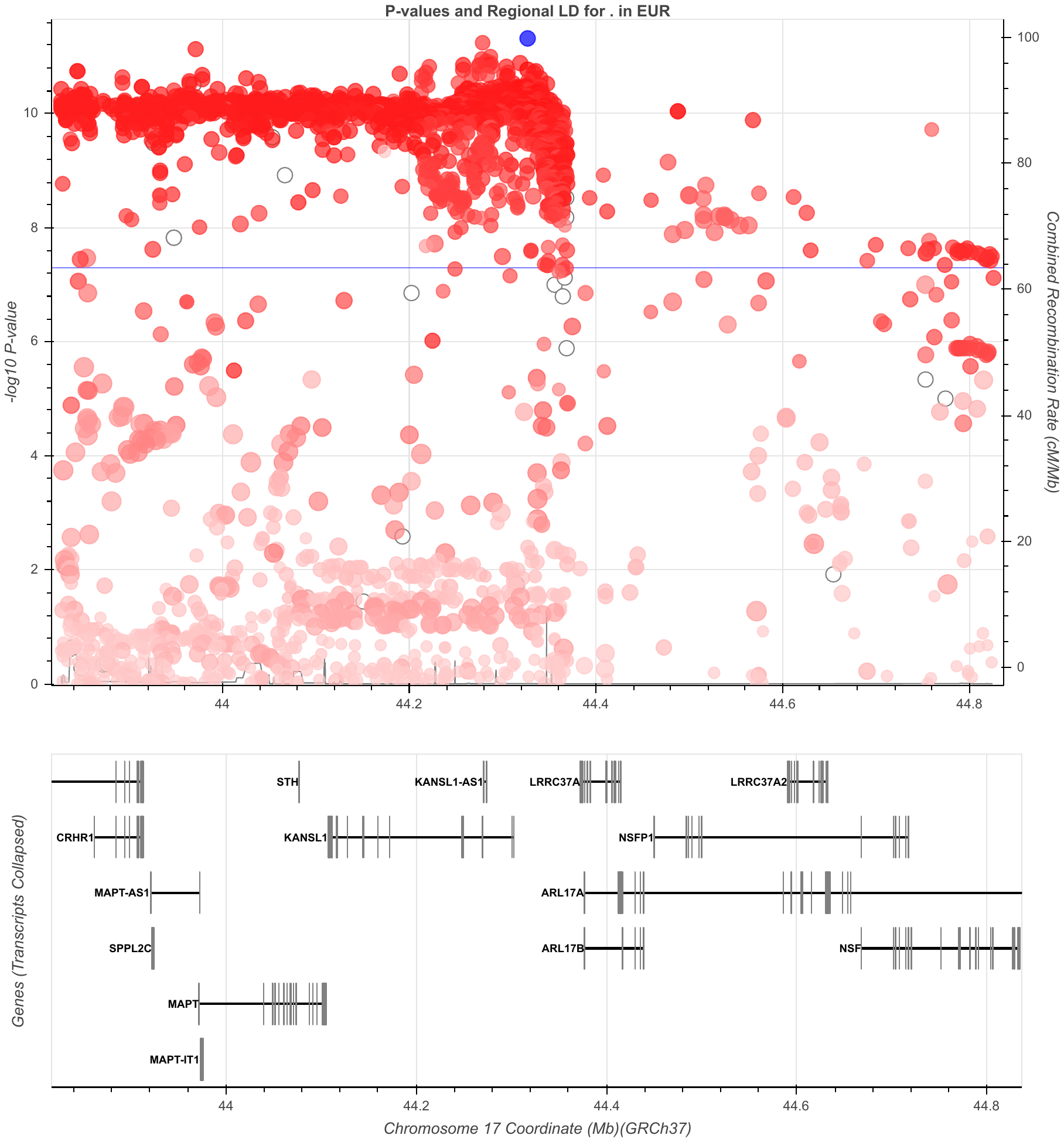
 **Figure S34. PA chr17 17:44326864 (rs2696625) region**
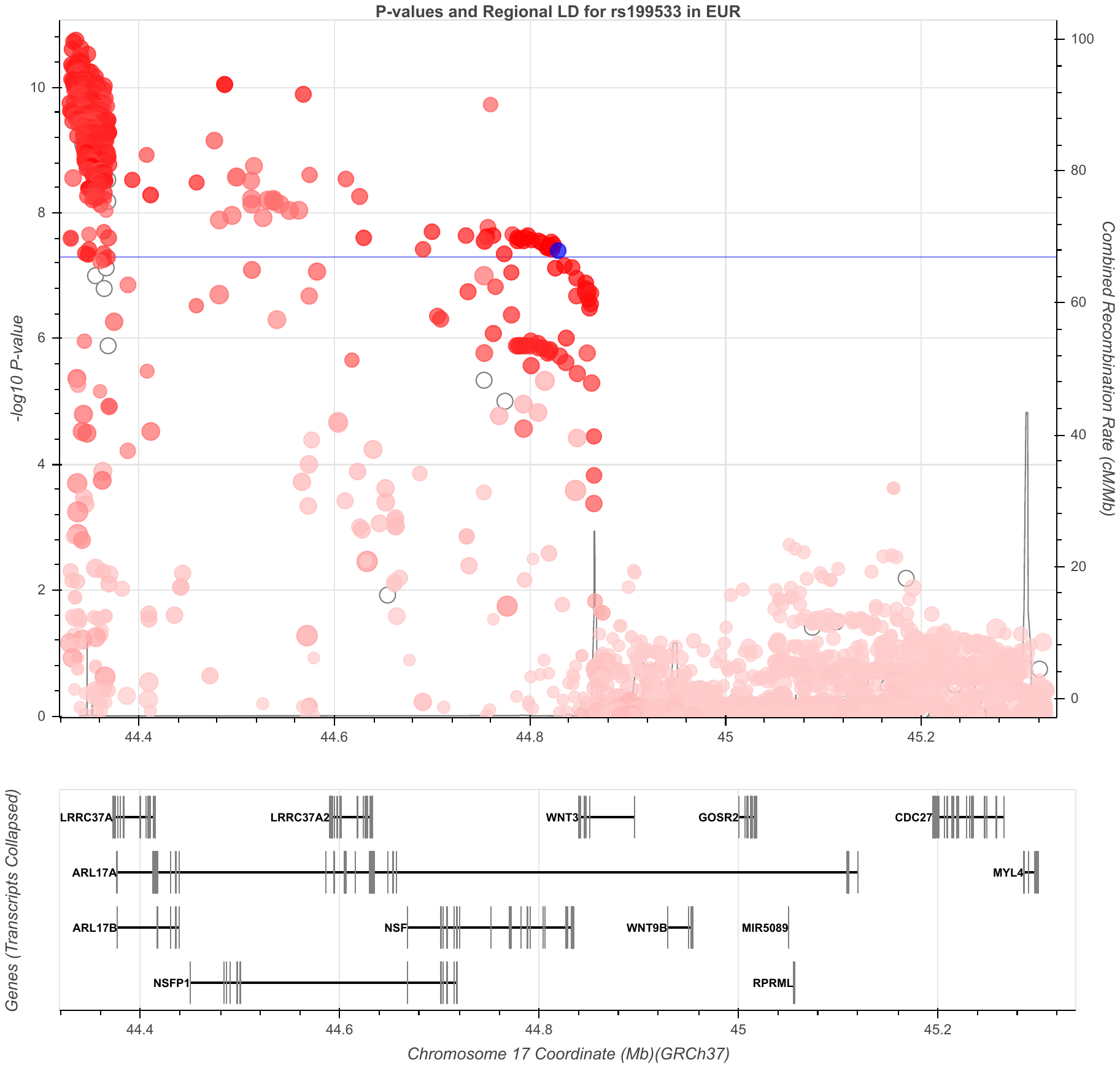
 **Figure S35. PA chr17 rs199533 region**
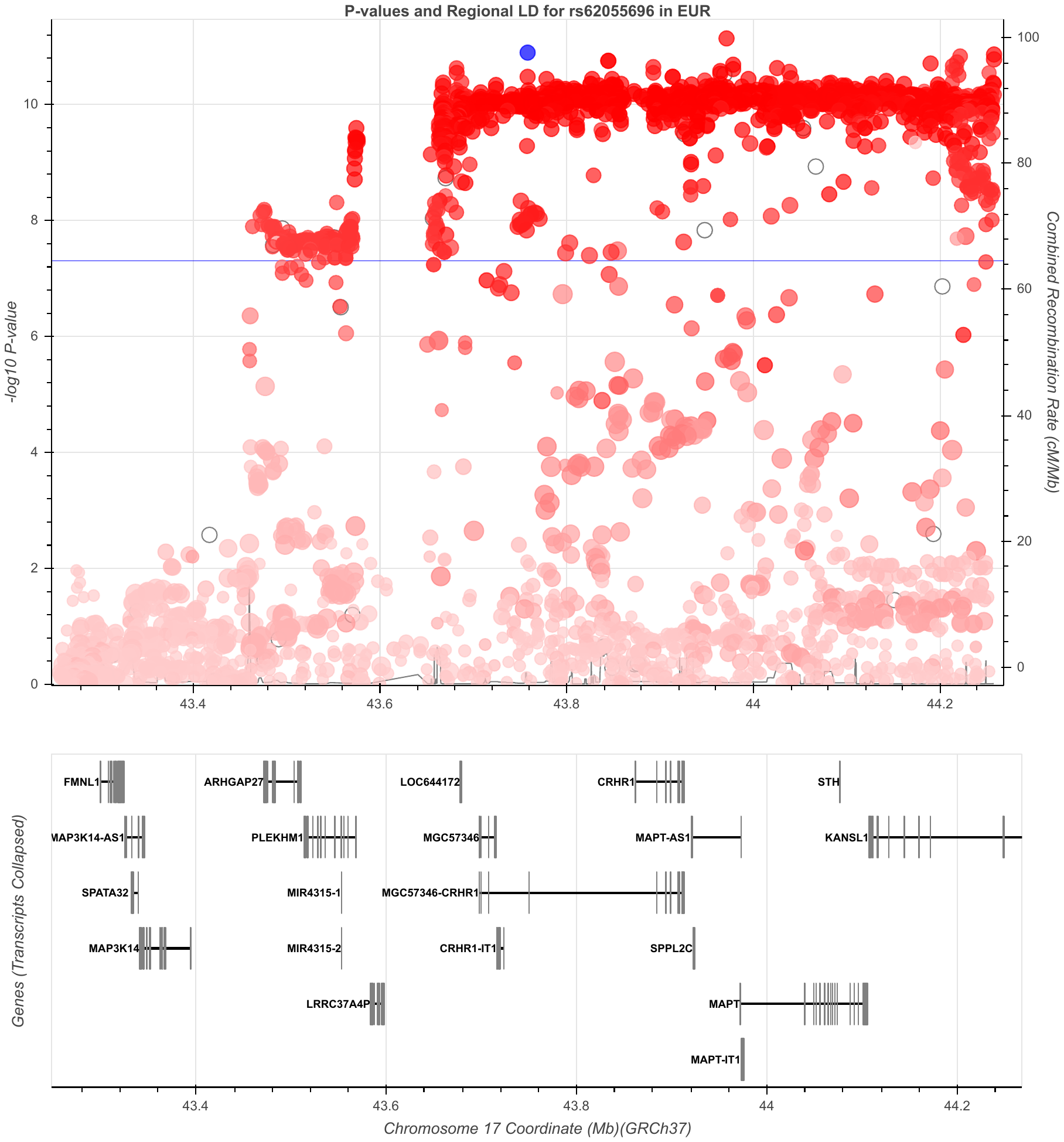
 **Figure S36. PA chr17 rs62055696 region**
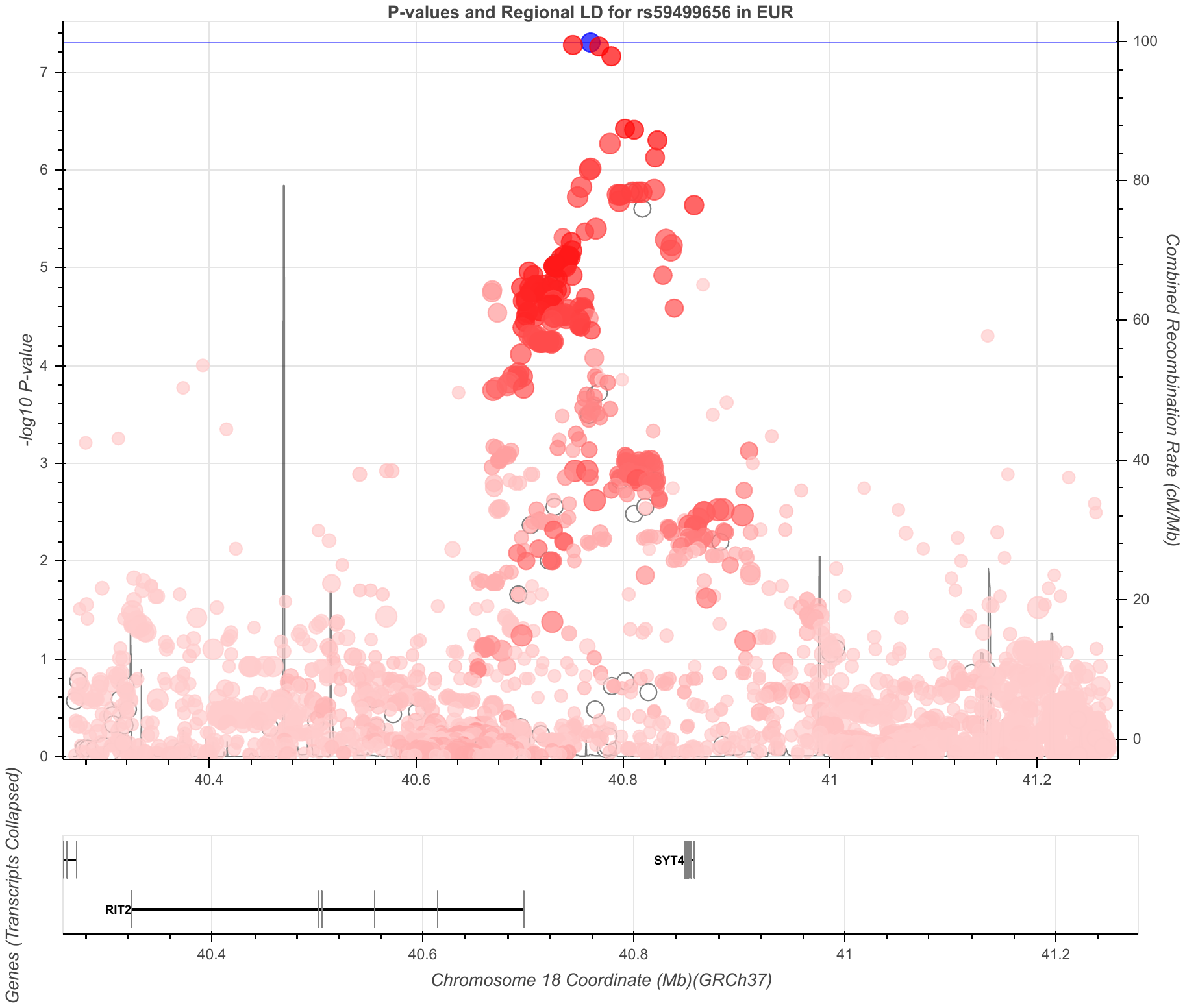

**Figure S37. PA chr18 rs59499656 region**

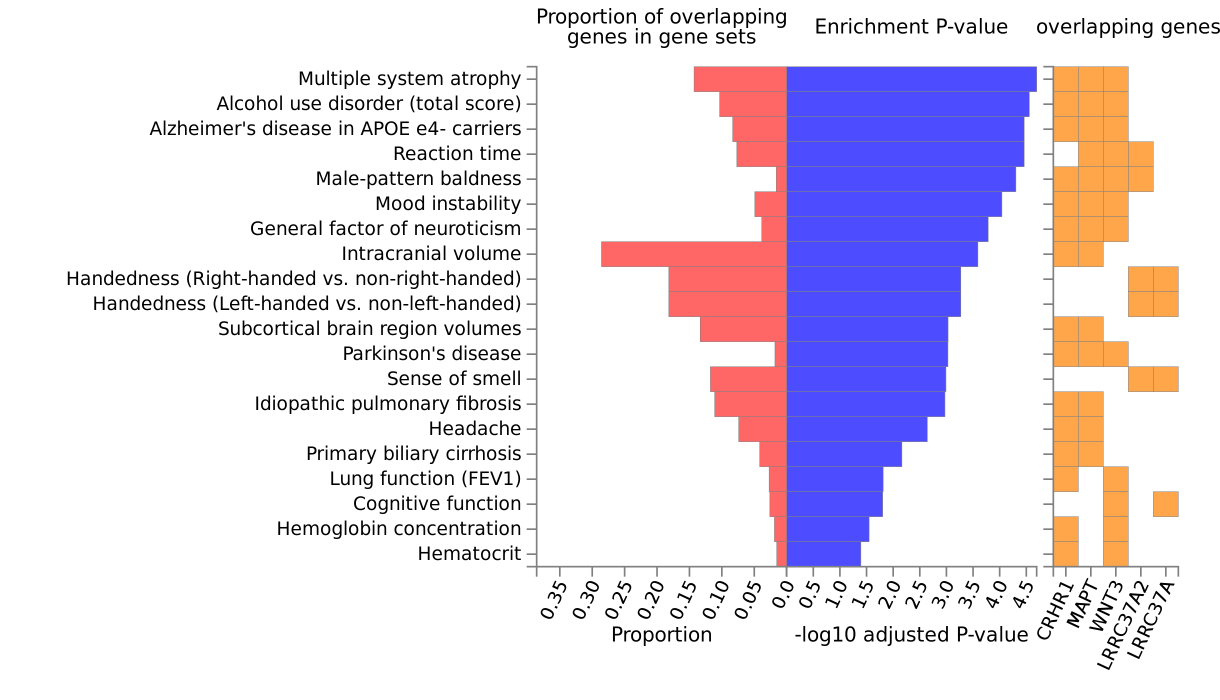

**Figure S38. CRF-vo2max functional candidates enriched in GWAS catalog traits**. TWAS-significant genes listed were enriched in all GWAS shown (FDR-adjusted p < 0.05).

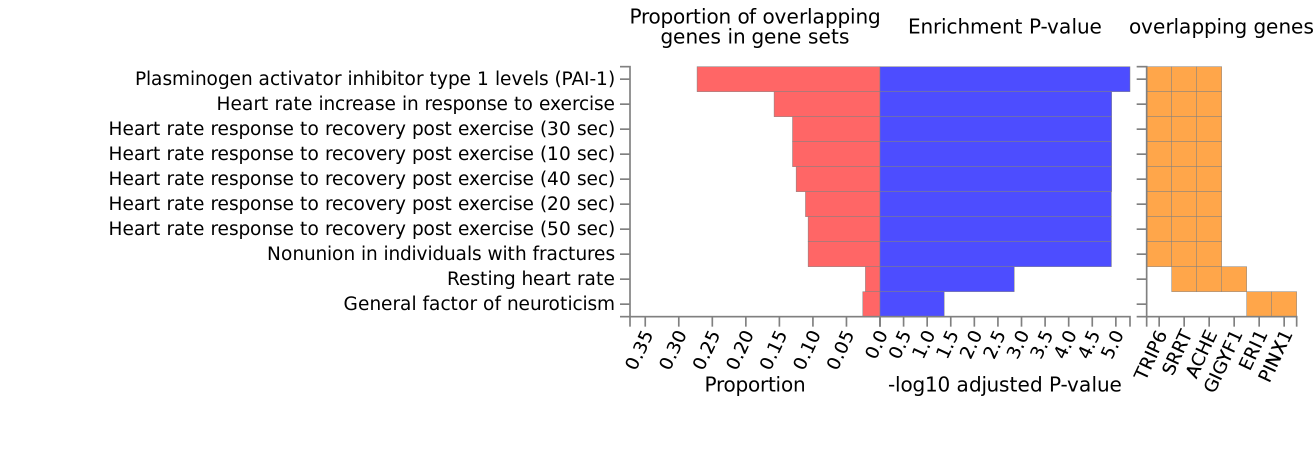

**Figure S39. CRF-slope functional candidates enriched in GWAS catalog traits**. TWAS-significant genes listed were enriched in all GWAS shown (FDR-adjusted p < 0.05).

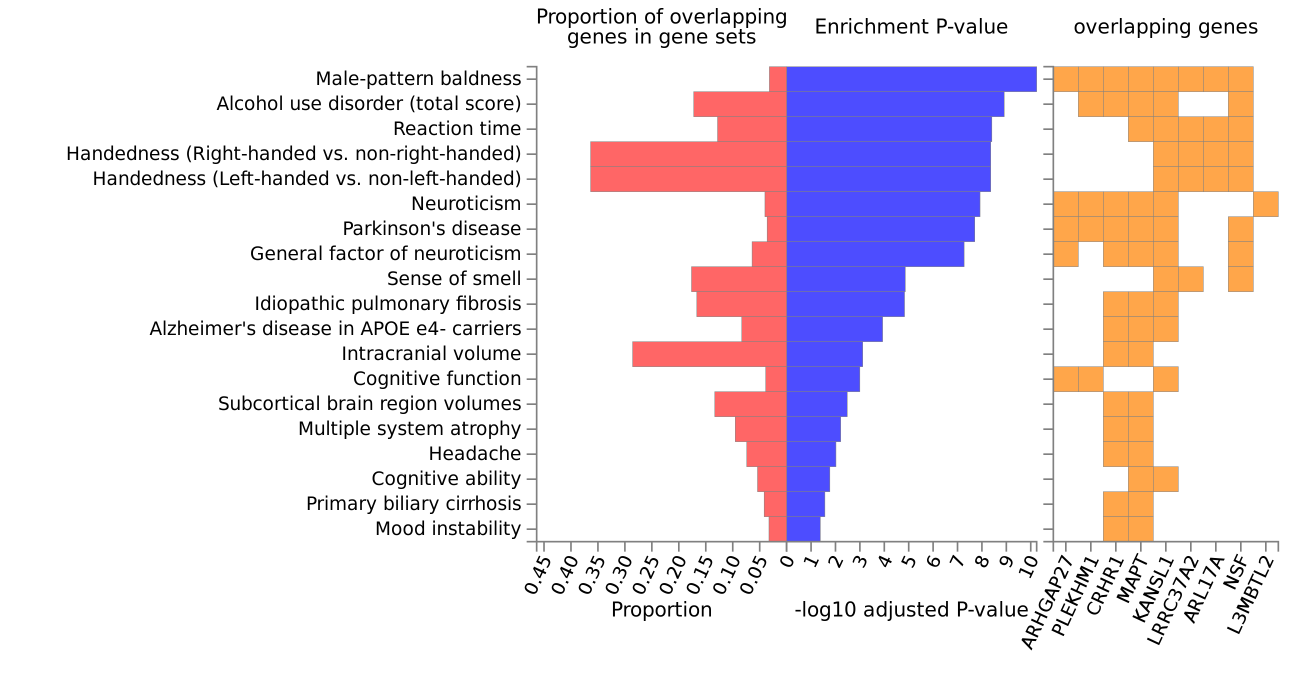

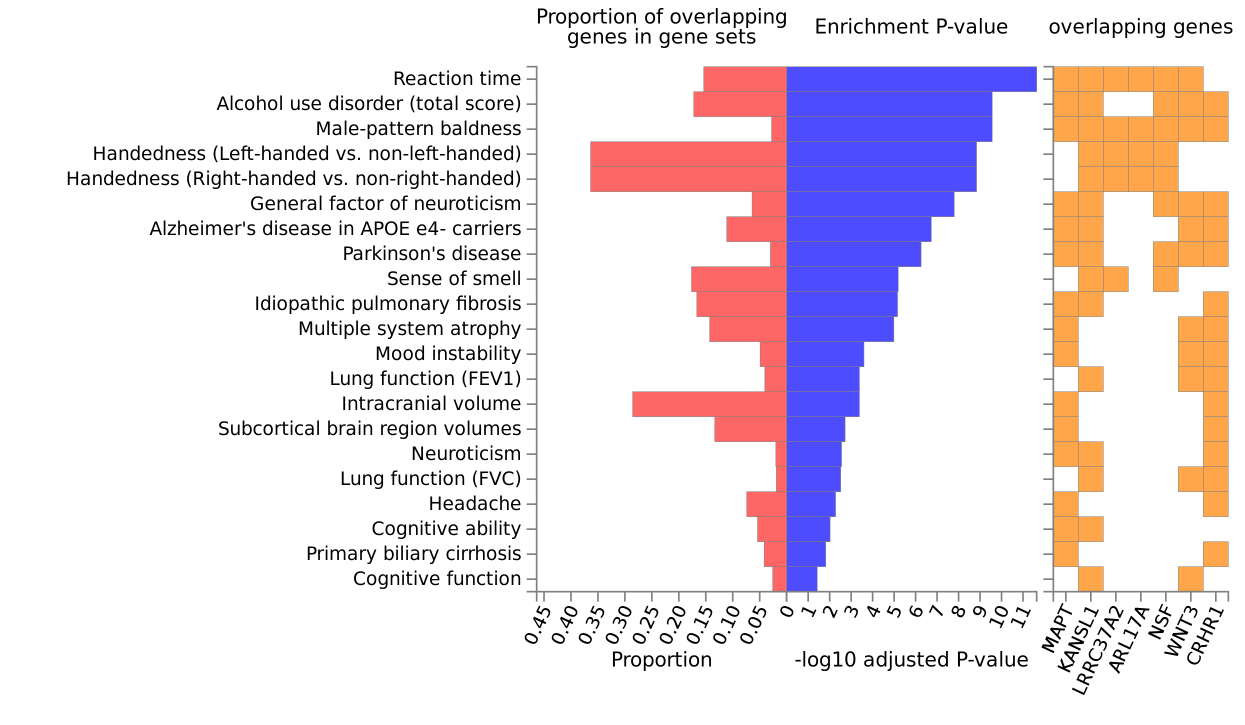
 **Figure S40. PA functional candidates enriched in GWAS catalog traits**. TWAS-significant genes listed were enriched in all GWAS shown (FDR-adjusted p < 0.05). Sex-combined PA functional candidates (top) and female PA functional candidates (below).

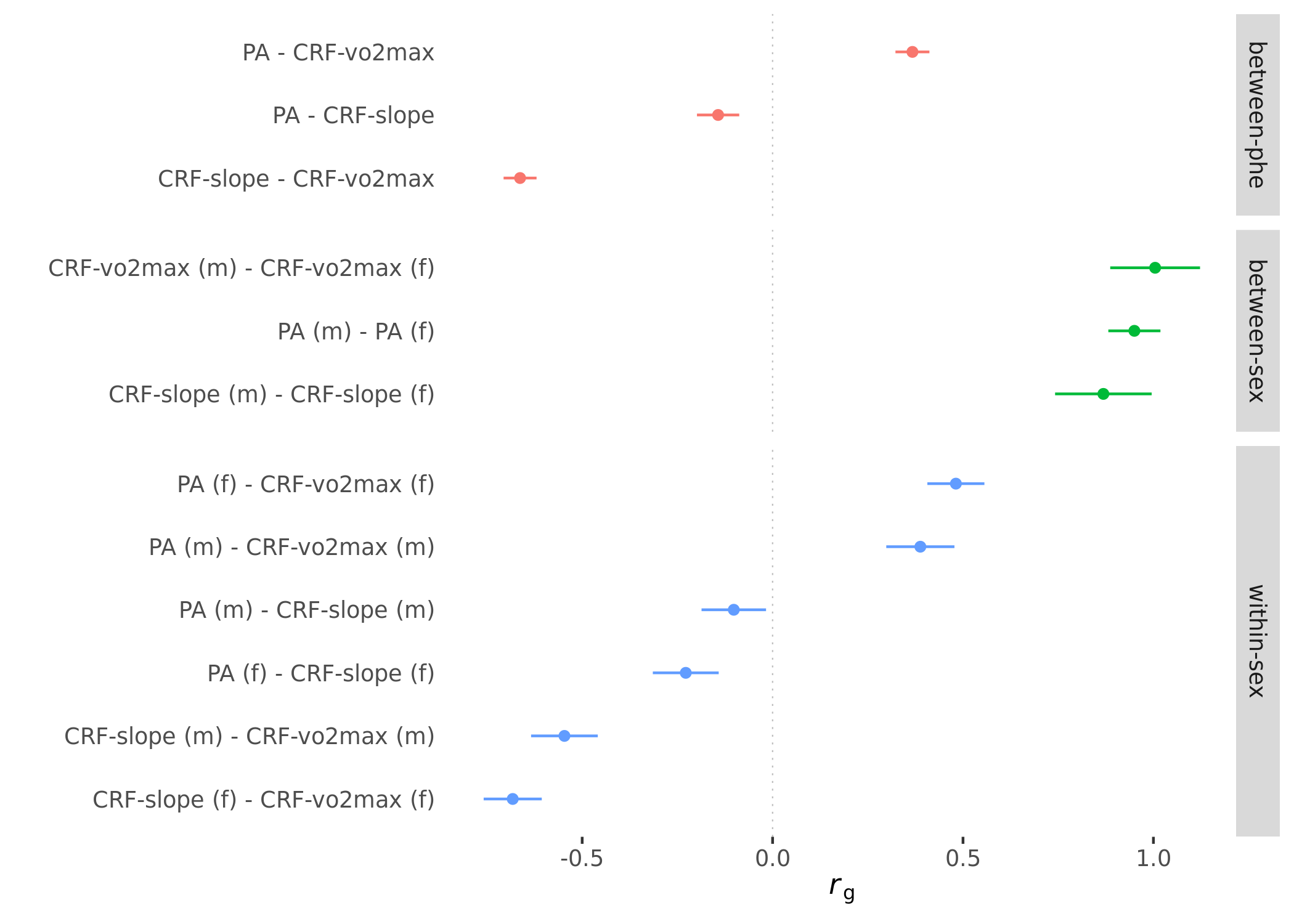

**Figure S41. Genetic correlations between CRF and PA by sex**. Summary statistics derived genetic correlations with error bars representing standard errors. The genetic correlation is not bounded between -1 and 1 and can validly be a little lower or higher than these values. Genetic correlations are shown for males and females combined **between-phenotype**, within-phenotype **between-sex**, and between-phenotype **within-sex**. (m) = male; (f) = female; *r*_g_ = genetic correlation.

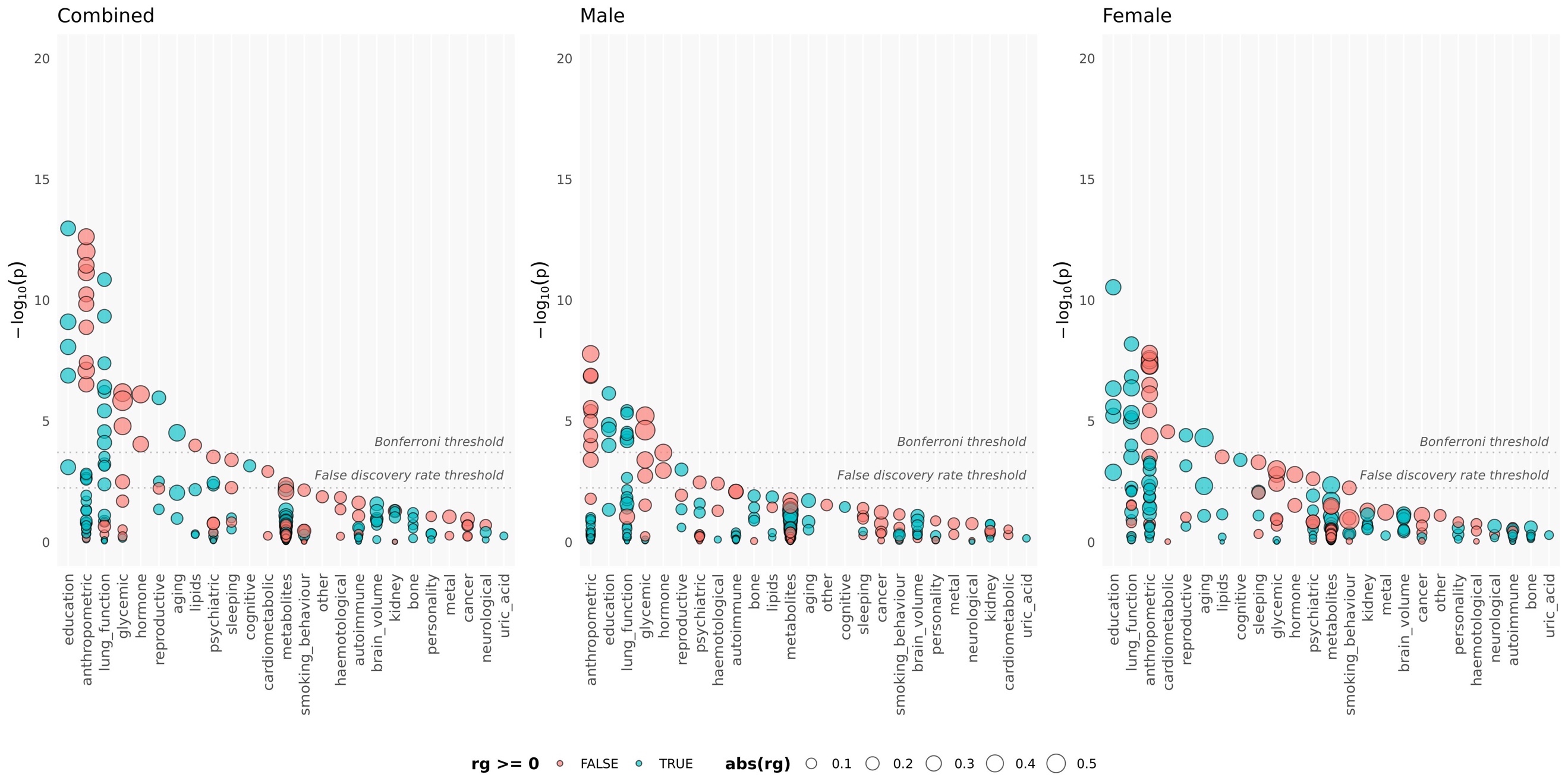

**Figure S42**. Genetic correlations between CRF-vo2max and LD Hub traits by sex

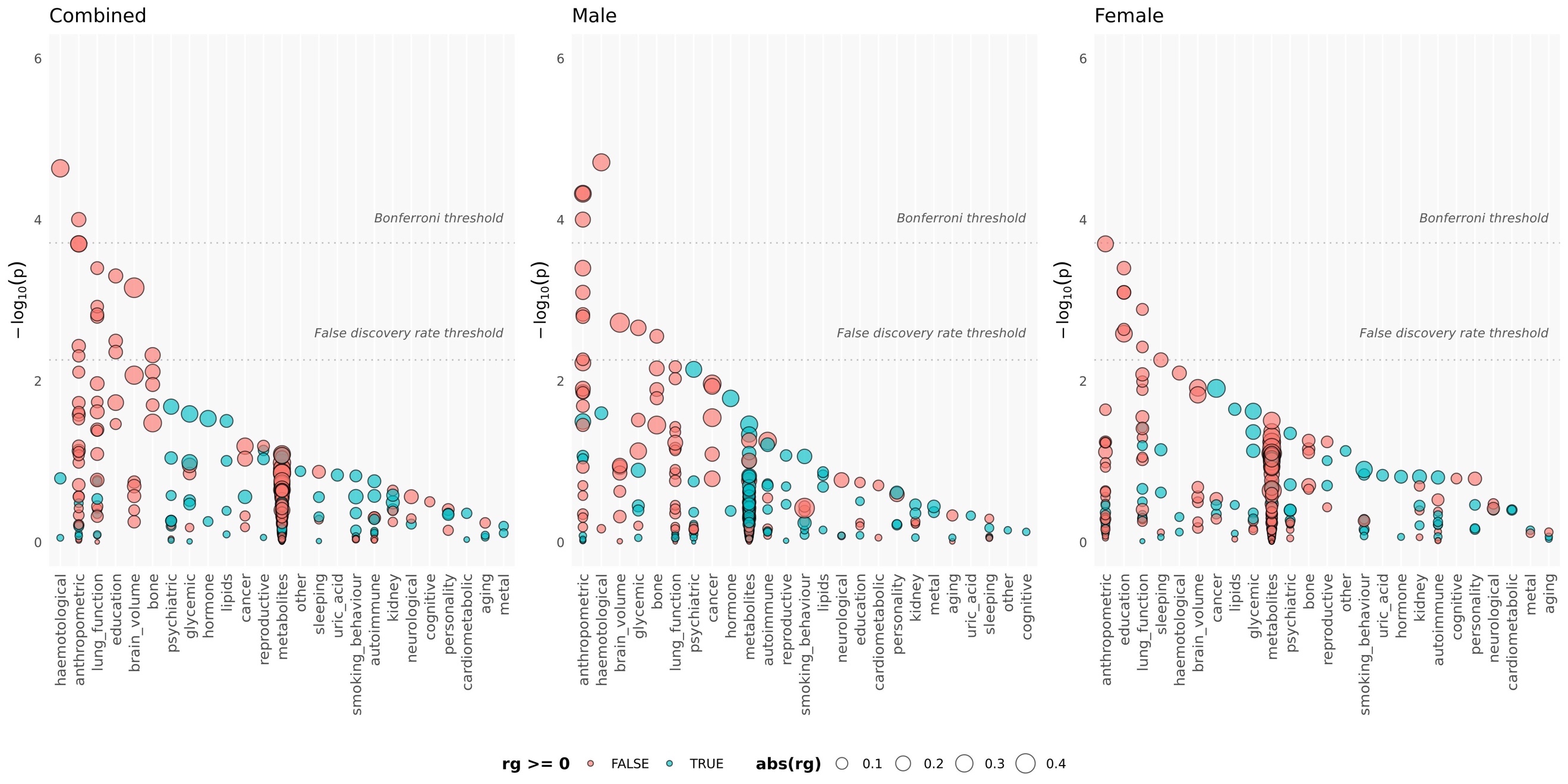

**Figure S43**. Genetic correlations between CRF-slope and LD Hub traits by sex

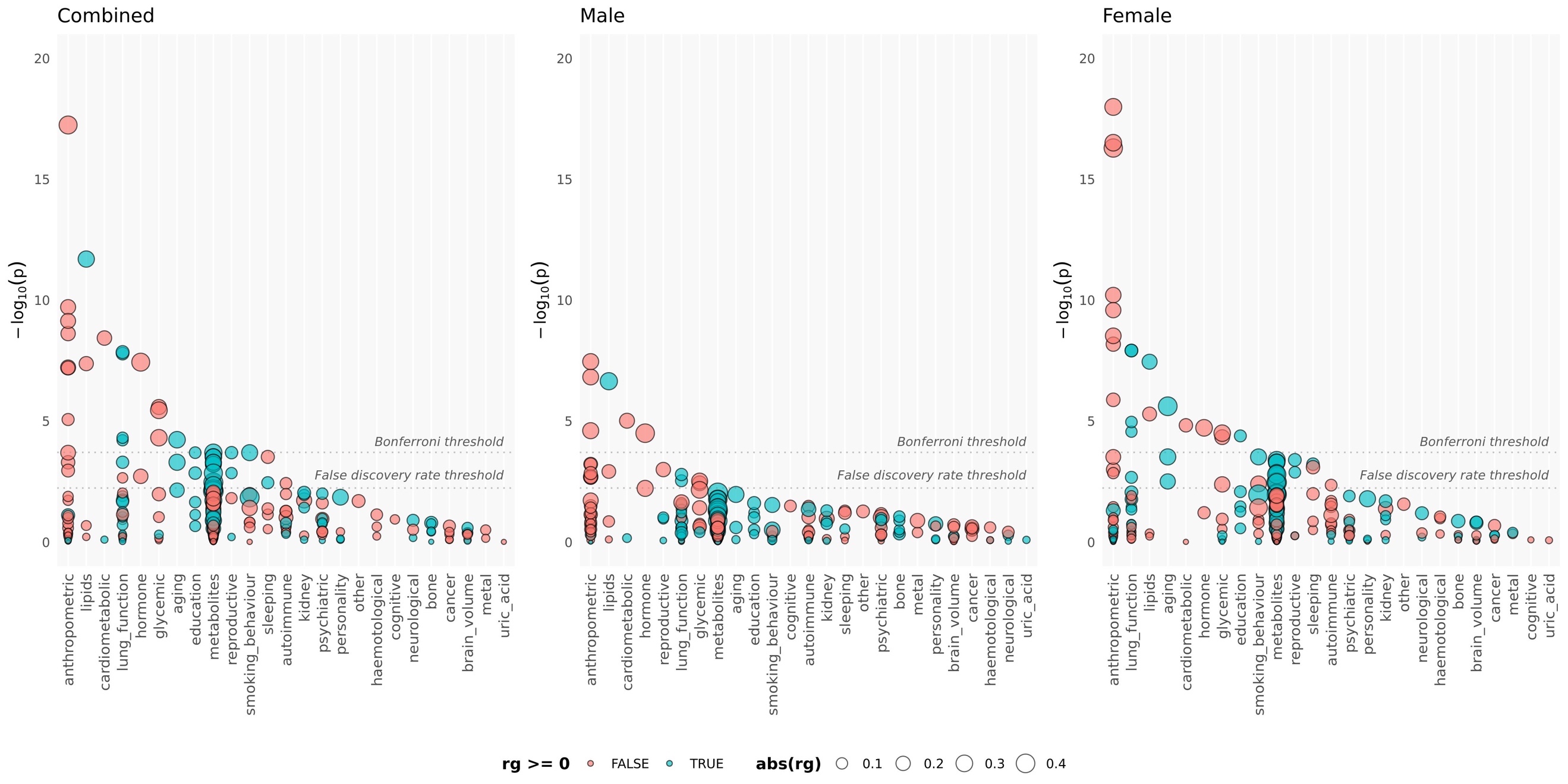

**Figure S44**. Genetic correlations between PA and LD Hub traits by sex

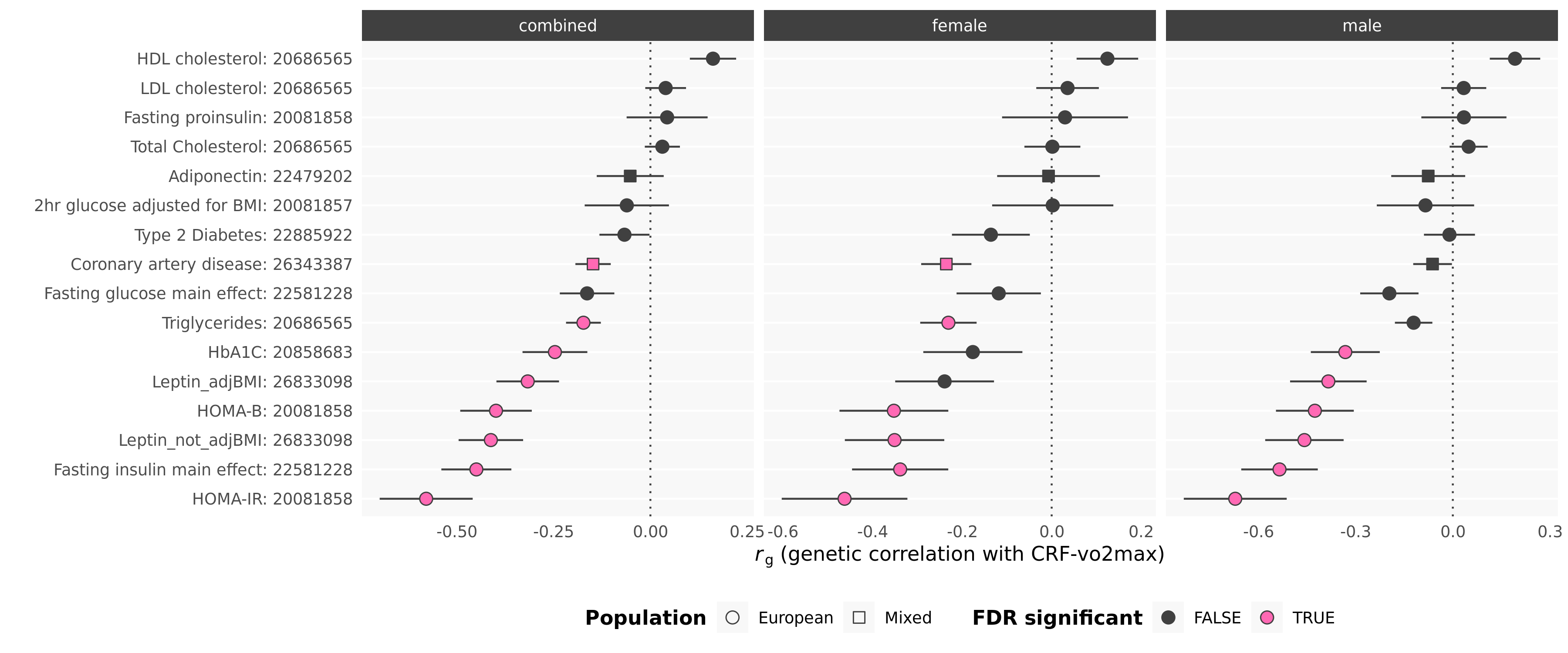

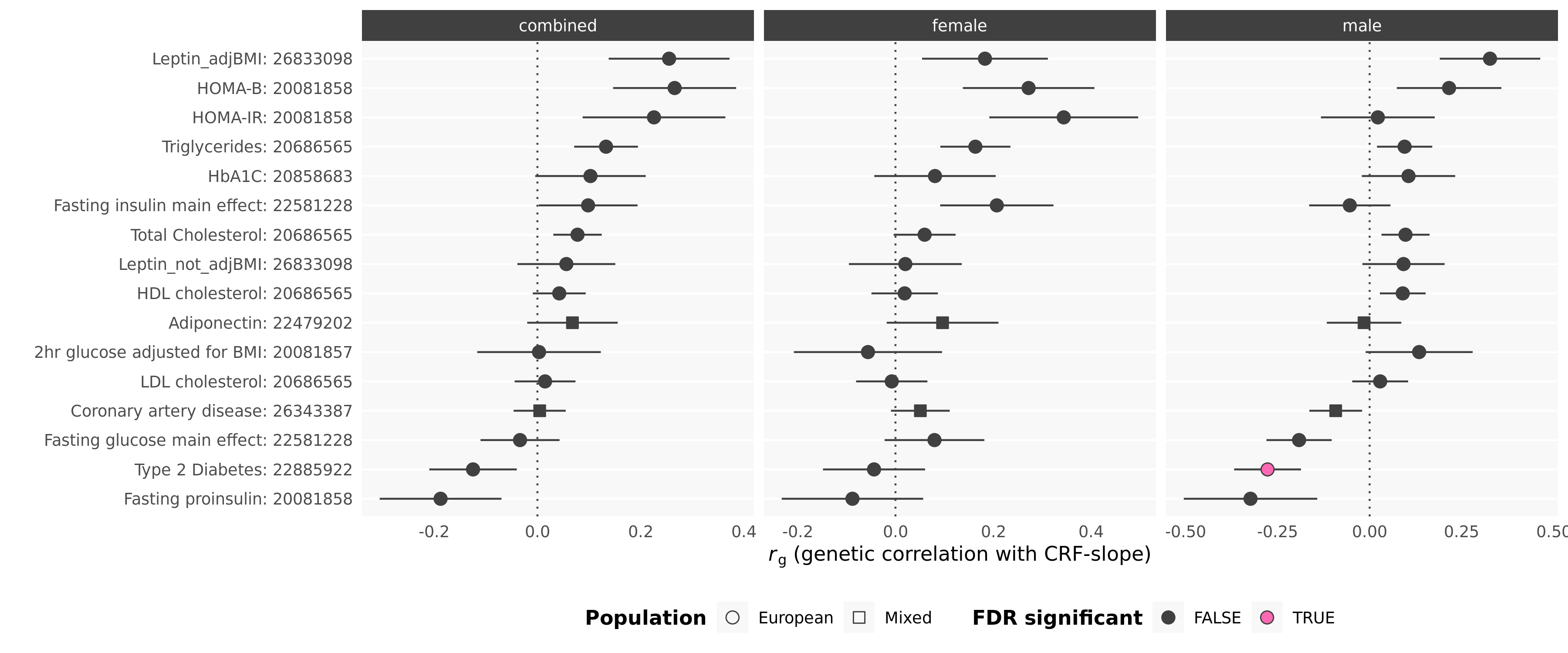

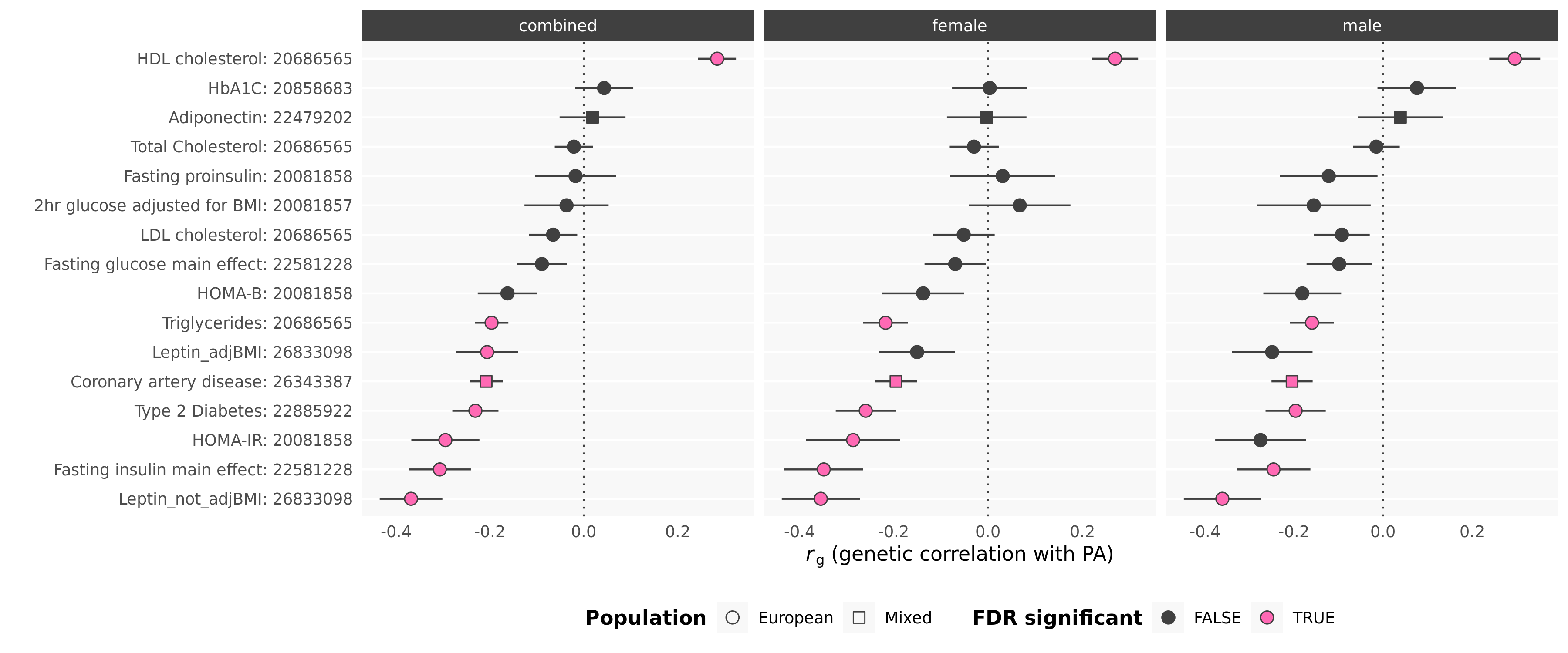

**Figure S45. Genetic correlations with glycaemic, cardiometabolic, and lipid traits.** GWASs listed as GWAS Trait: PMID (where PMID was not available it is listed as 0)

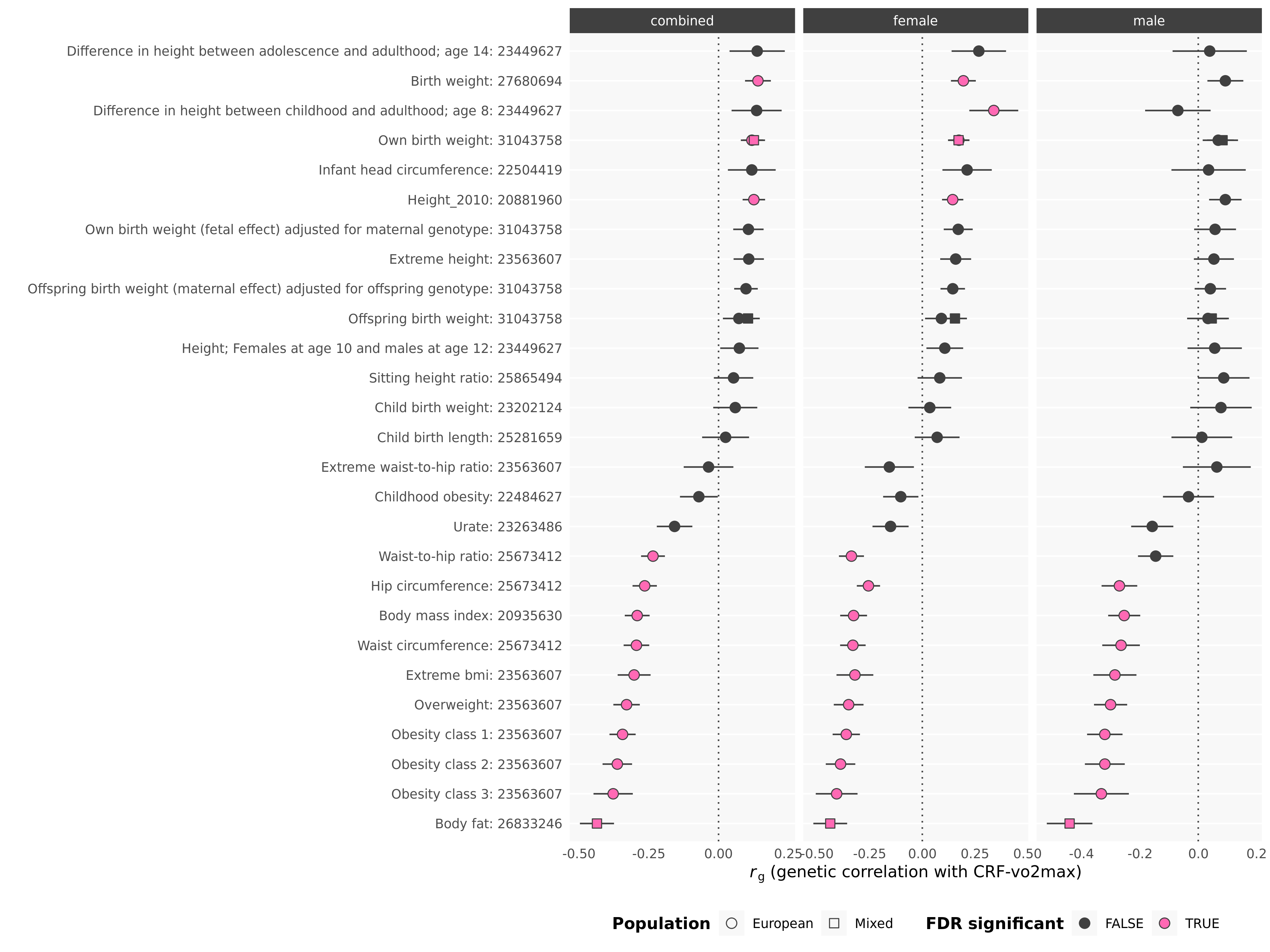

**Figure S46. Genetic correlations with anthropometric measures.** GWASs listed as GWAS Trait: PMID (where PMID was not available it is listed as 0)

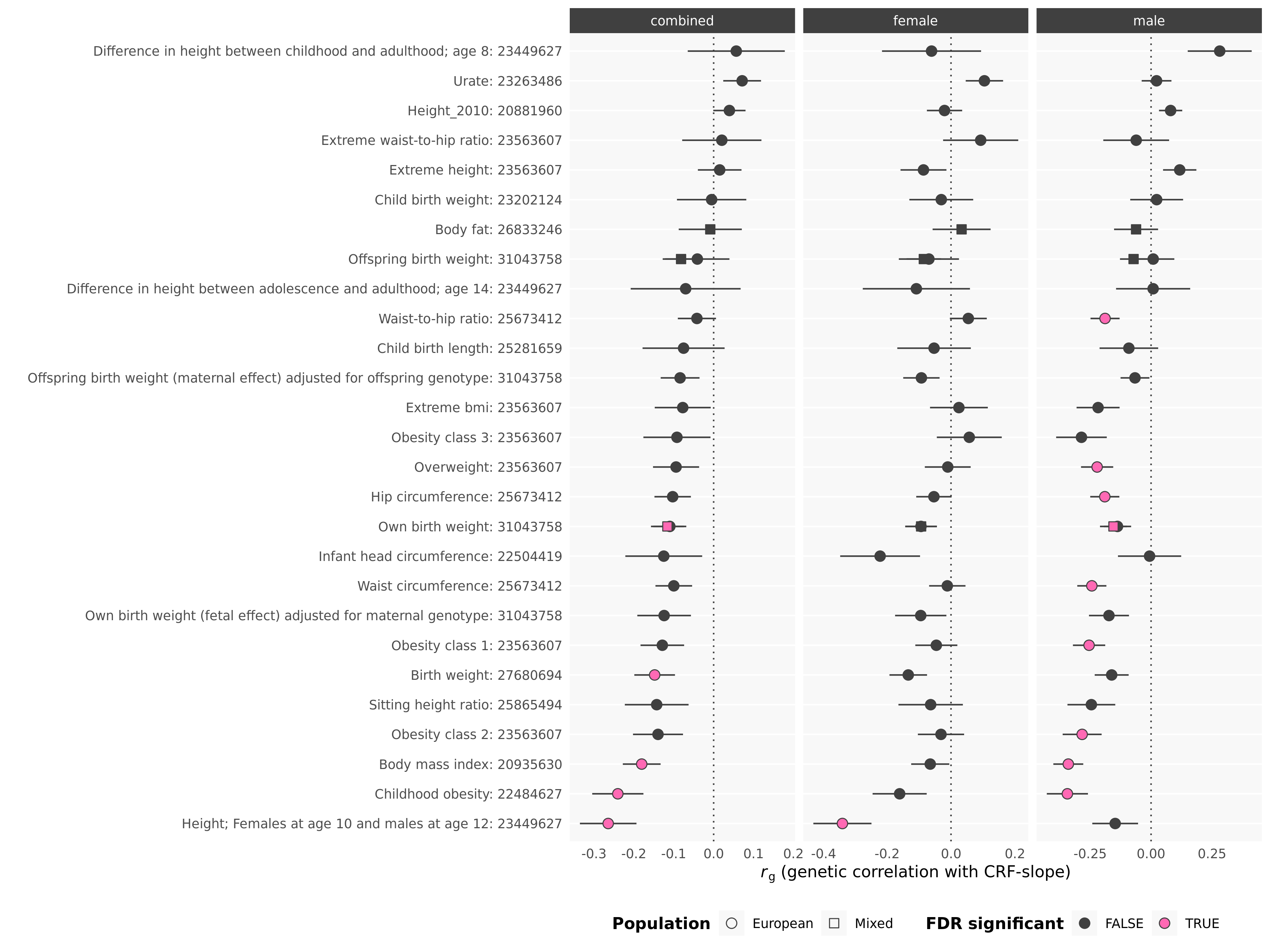

**Figure S47. Genetic correlations with anthropometric measures.** GWASs listed as GWAS Trait: PMID (where PMID was not available it is listed as 0)

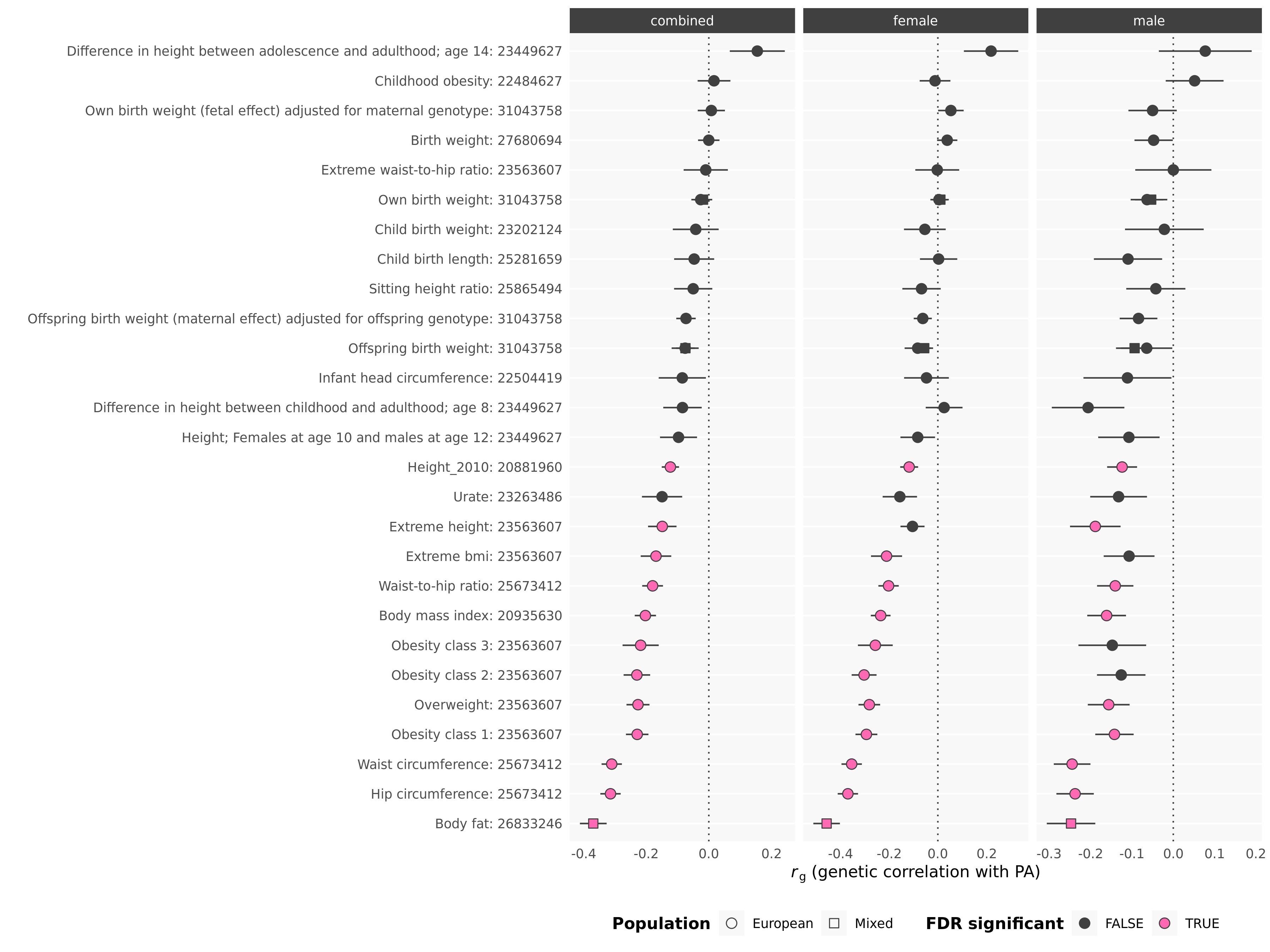

**Figure S48. Genetic correlations with anthropometric measures.** GWASs listed as GWAS Trait: PMID (where PMID was not available it is listed as 0)

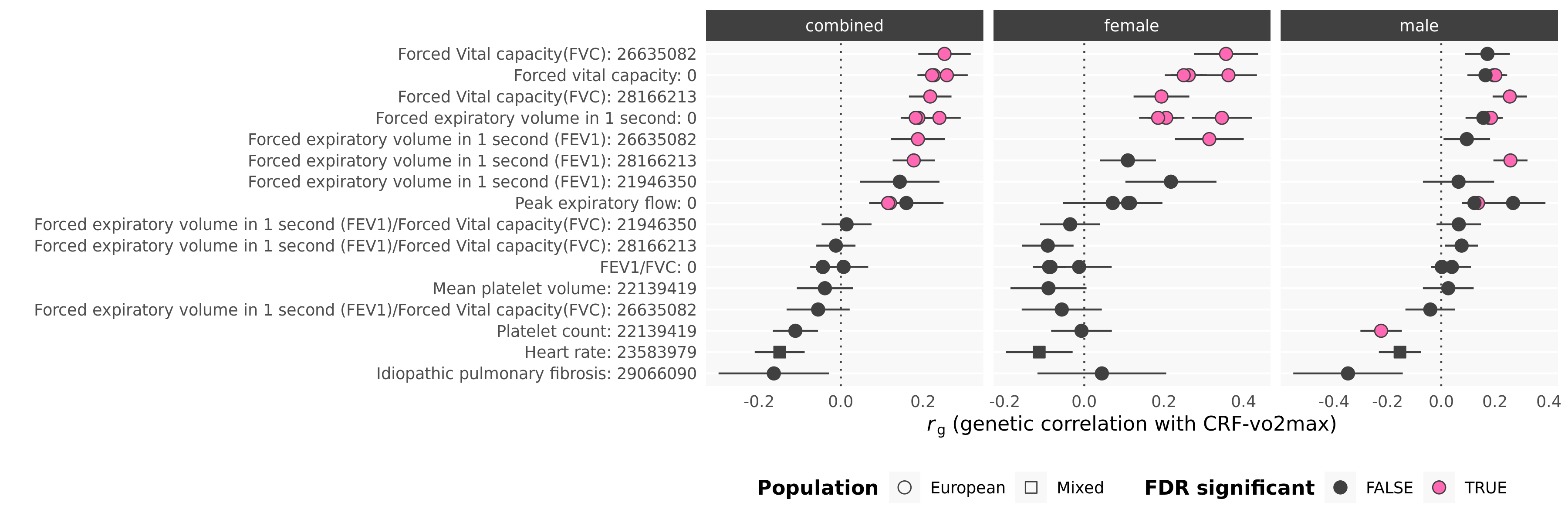

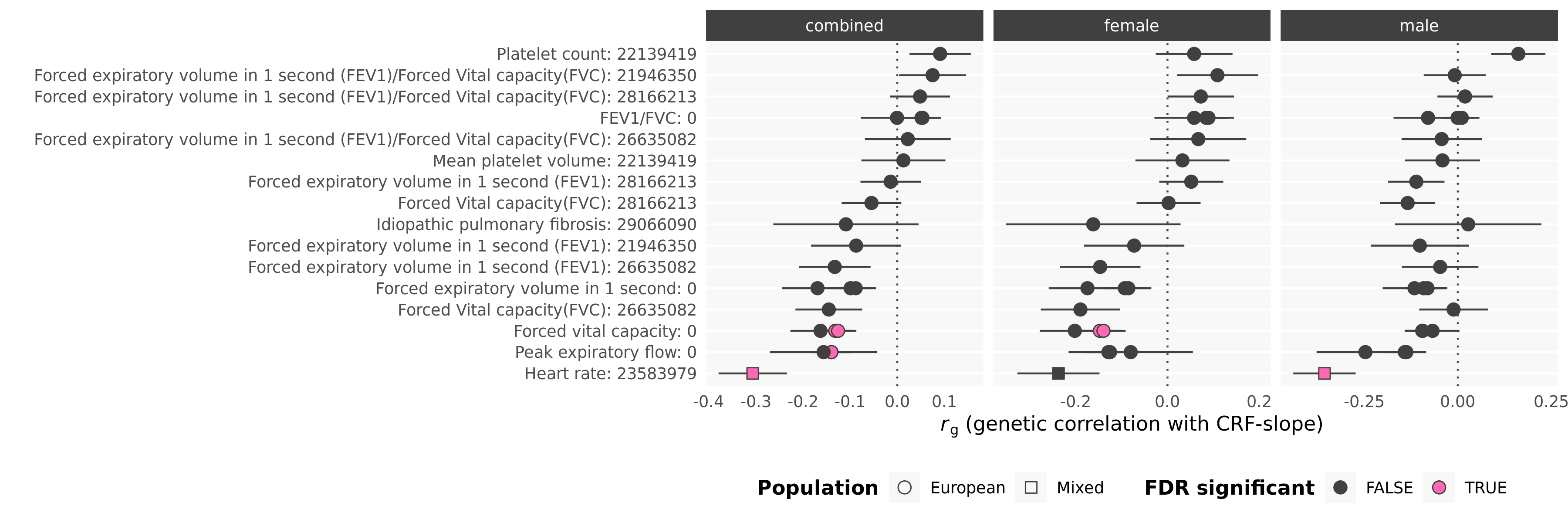

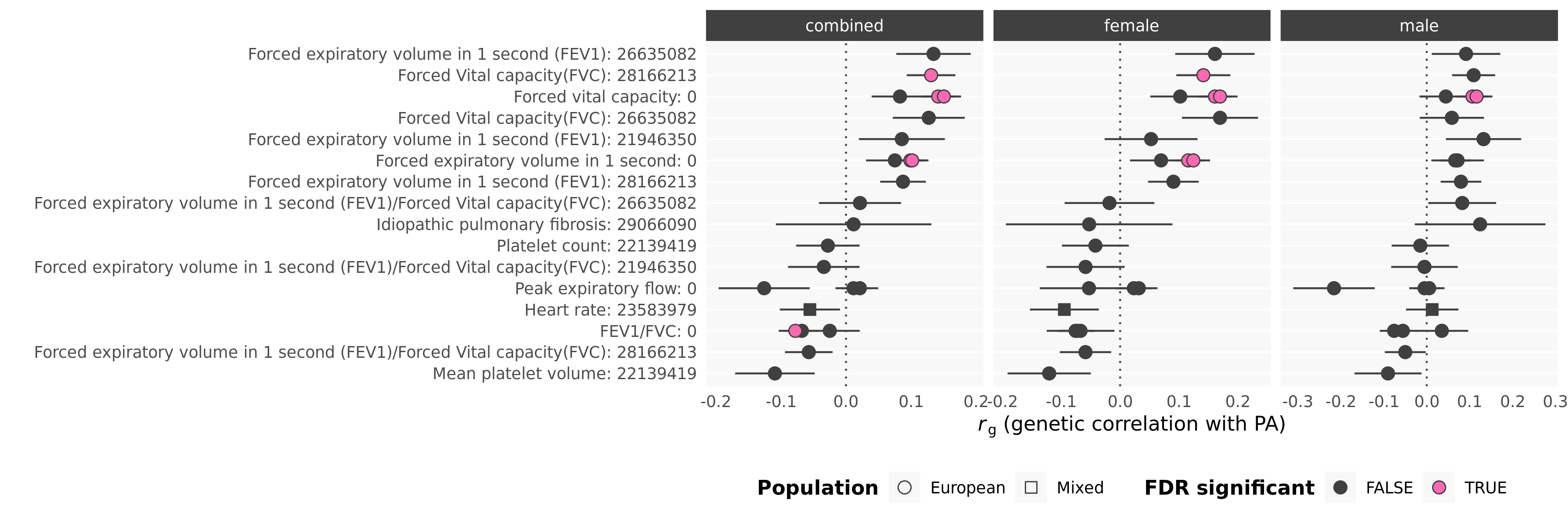

**Figure S49. Genetic correlations with lung function.** GWASs listed as GWAS Trait: PMID (where PMID was not available it is listed as 0)

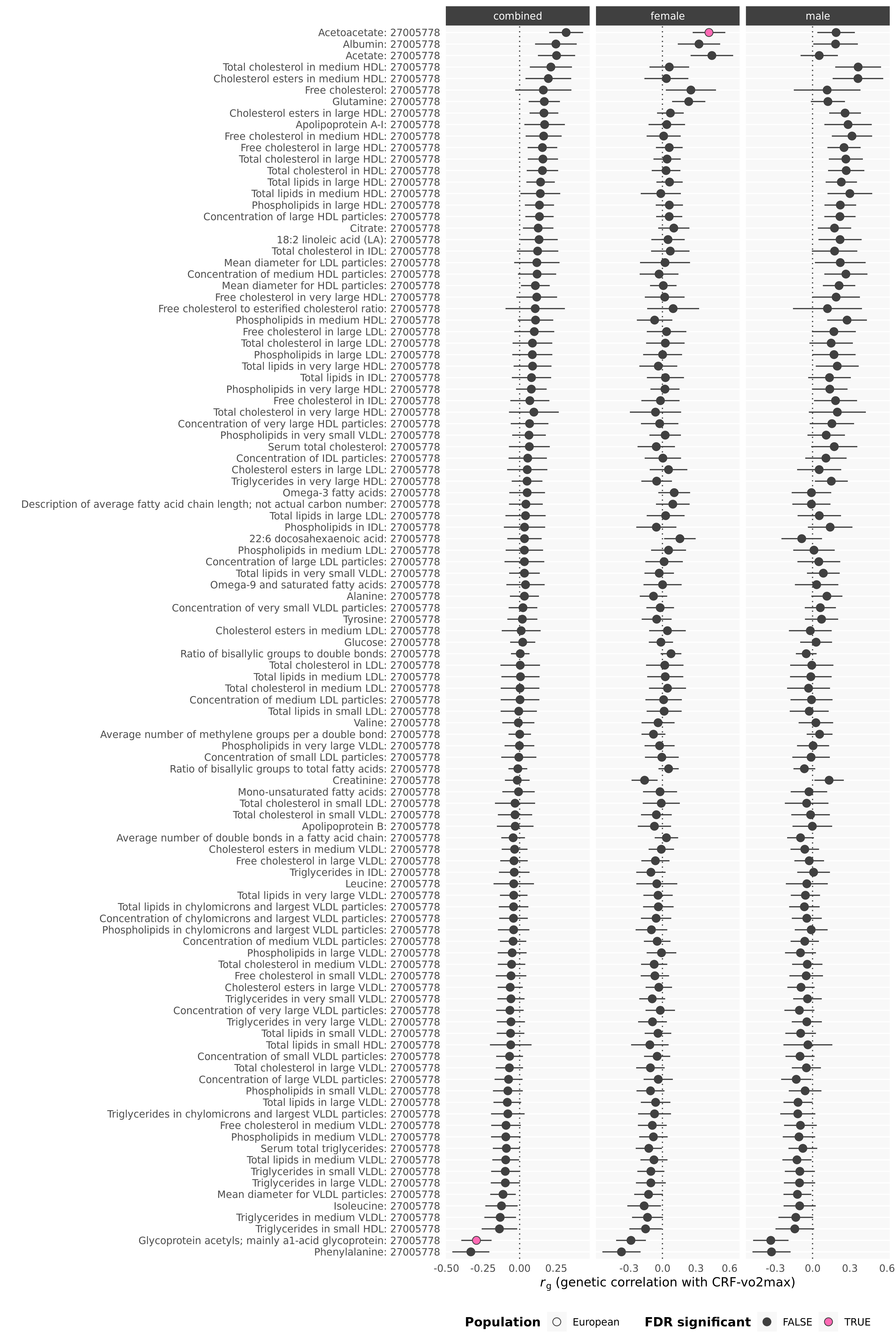

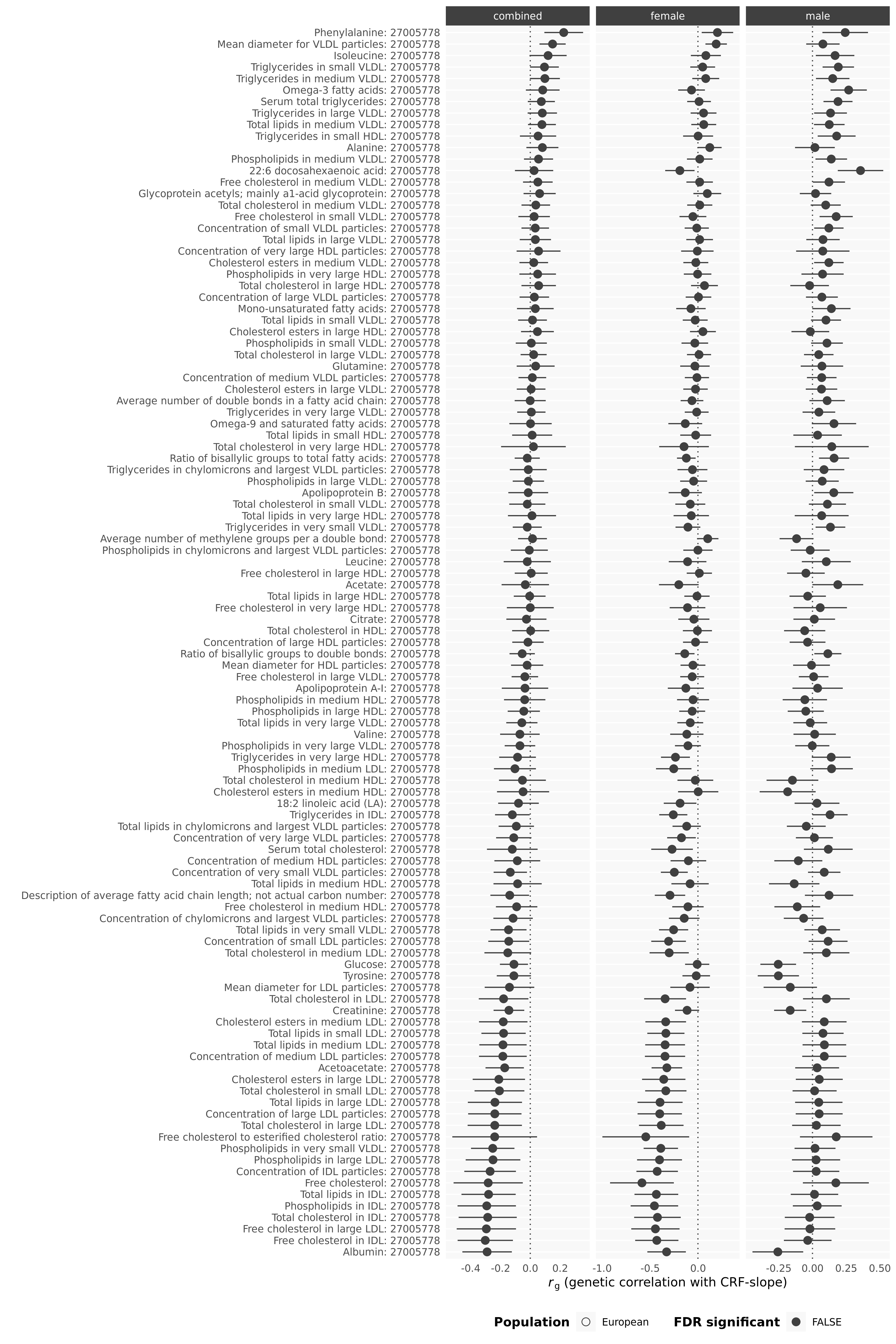

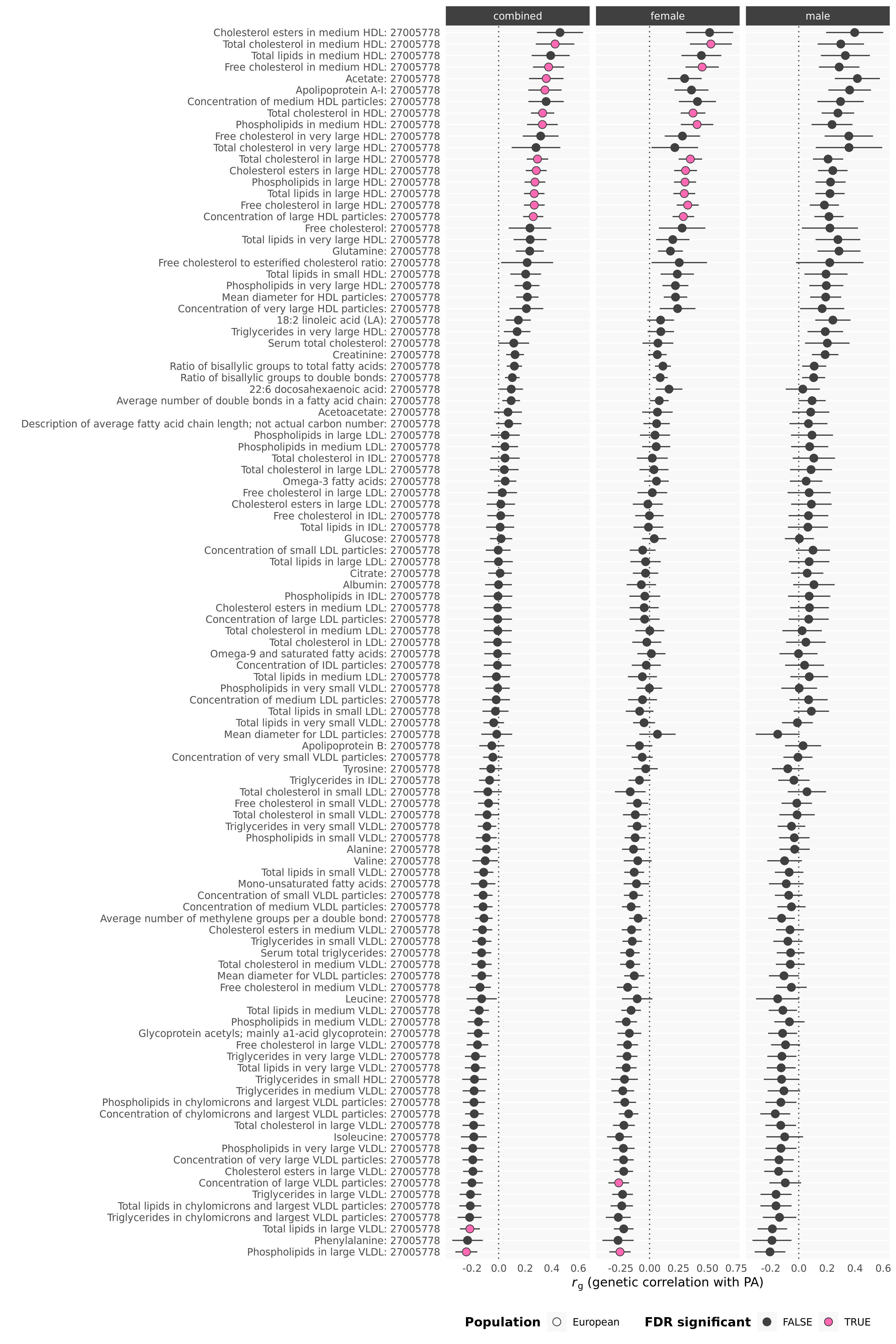

**Figure S50. Genetic correlation with blood metabolites**

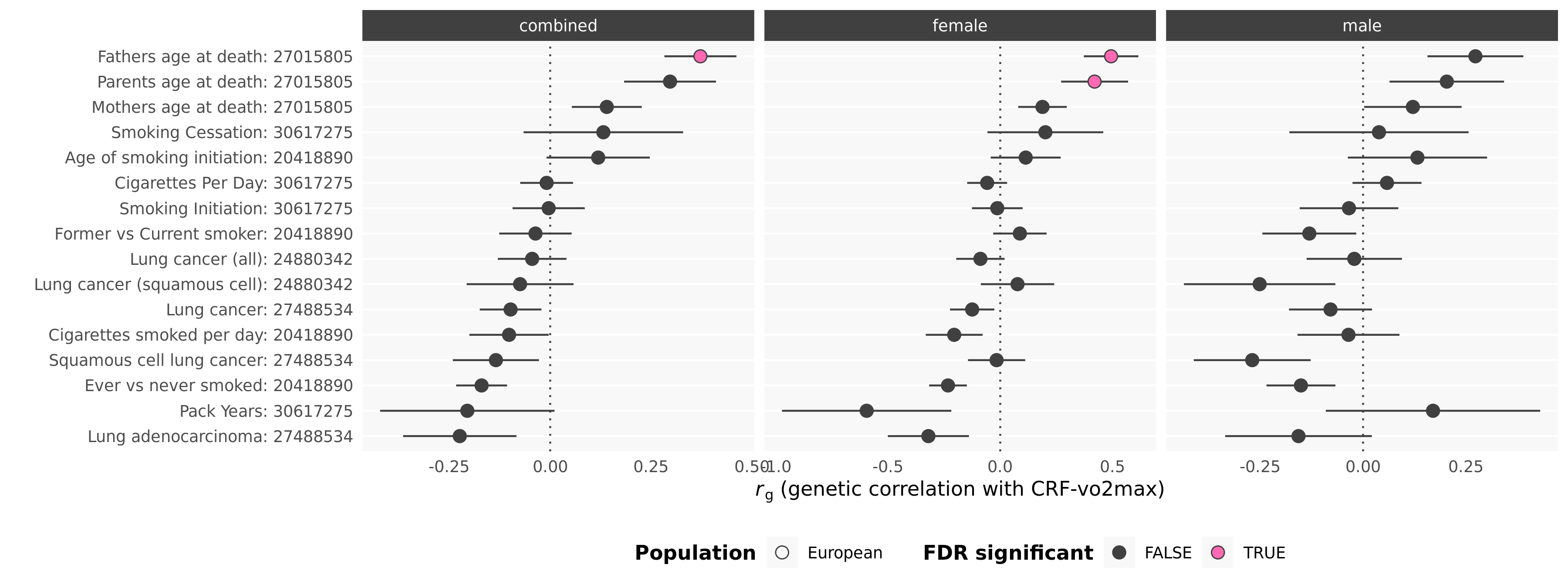

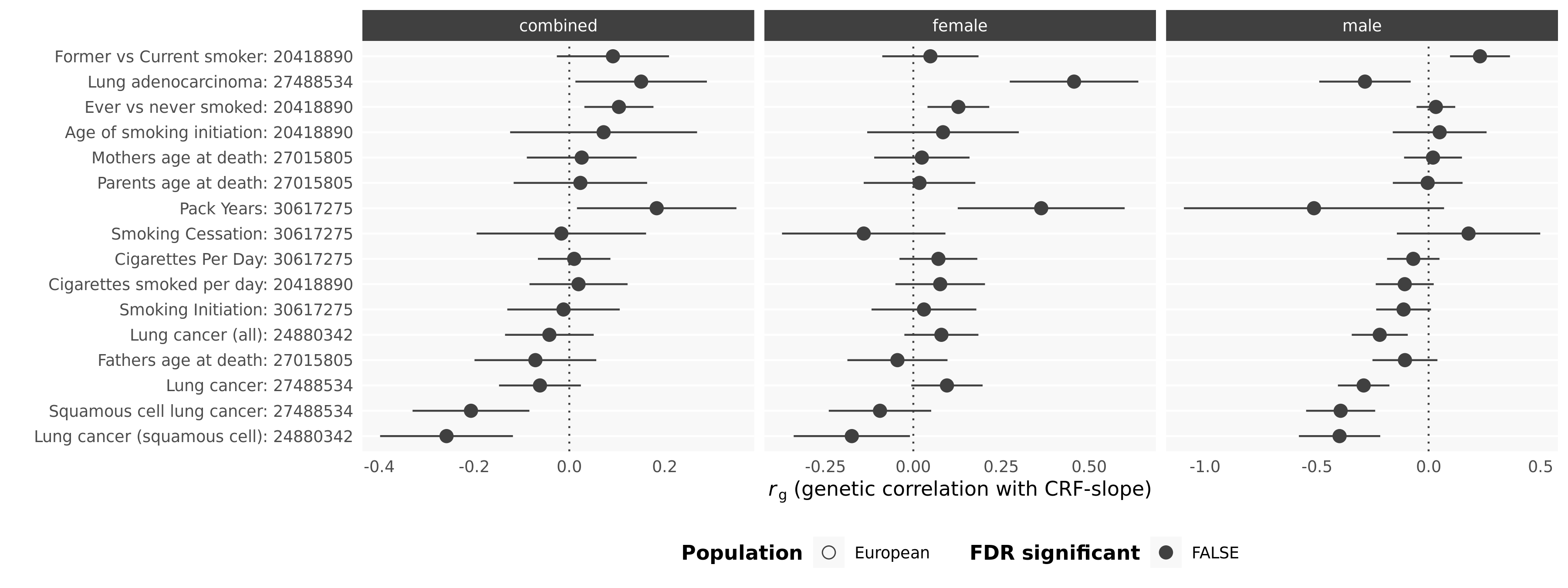

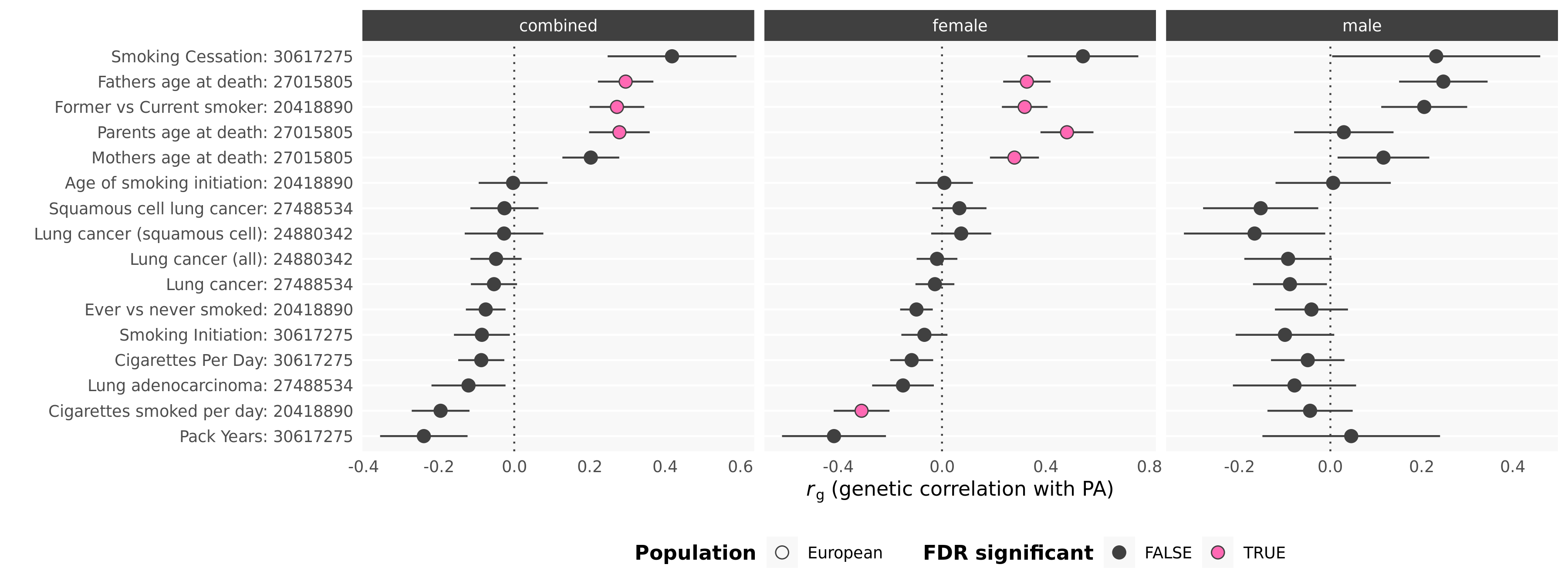

**Figure S51. Genetic correlations with smoking behaviour, cancer and longevity**

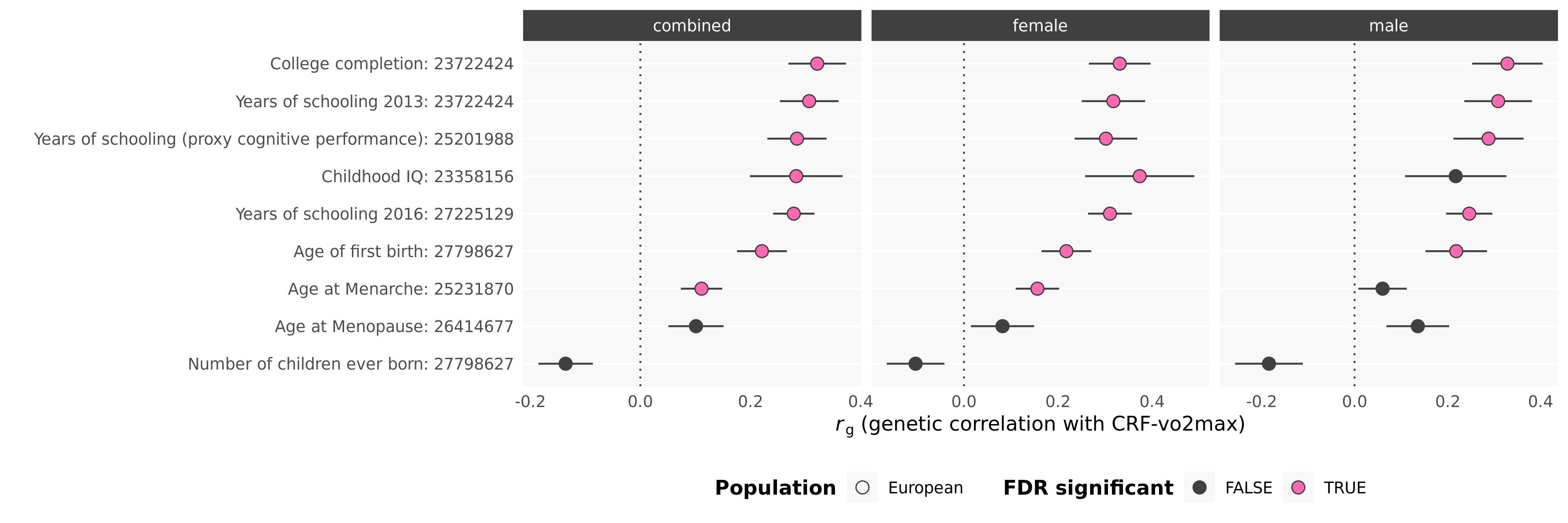

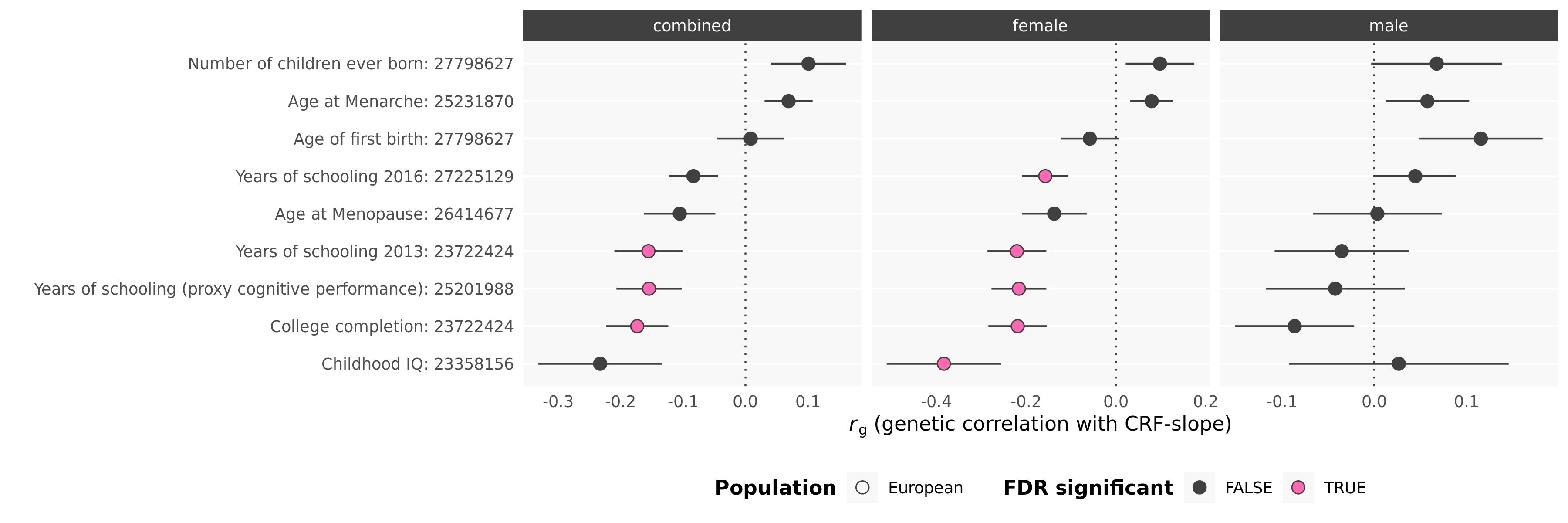

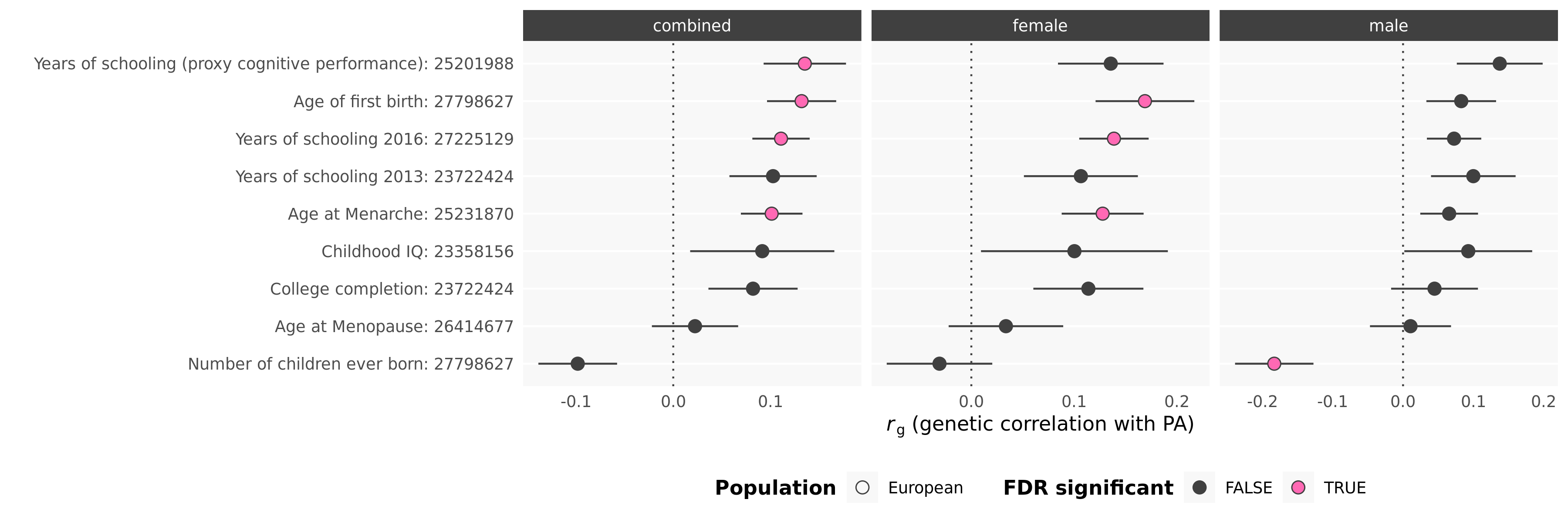

**Figure S52. Genetic correlations with education and reproductive measures**
